# The projected prevalence of comorbidities and multimorbidity in people with HIV in the United States through the year 2030

**DOI:** 10.1101/2022.11.04.22281891

**Authors:** Keri N Althoff, Cameron Stewart, Elizabeth Humes, Lucas Gerace, Cynthia Boyd, Kelly Gebo, Amy C Justice, Emily Hyle, Sally Coburn, Raynell Lang, Michael J Silverberg, Michael Horberg, Viviane D Lima, M John Gill, Maile Karris, Peter F Rebeiro, Jennifer Thorne, Ashleigh J Rich, Heidi Crane, Mari Kitahata, Anna Rubtsova, Cherise Wong, Sean Leng, Vincent C Marconi, Gypsyamber D’Souza, Hyang Nina Kim, Sonia Napravnik, Kathleen McGinnis, Gregory D Kirk, Timothy R Sterling, Richard D Moore, Parastu Kasaie

**Affiliations:** Department of Epidemiology, Johns Hopkins Bloomberg School of Public Health, Baltimore, MD, USA; Division of Geriatric Medicine and Gerontology, Department of Medicine, Johns Hopkins School of Medicine, Baltimore, MD, USA; Department of Health Policy and Management, Johns Hopkins Bloomberg School of Public Health, Baltimore, MD, USA; Division of Infectious Diseases, Department of Medicine, Johns Hopkins School of Medicine, Baltimore, MD, USA; Yale Schools of Medicine and Public Health, New Haven, Connecticut, USA; Veterans Affairs Connecticut Healthcare System, West Haven, CT, USA; Harvard Medical School and the Division of Infectious Diseases, Massachusetts General Hospital, Boston, MA, USA; Harvard University Center for AIDS Research, Boston, MA, USA; Department of Medicine, University of Calgary, Calgary, Alberta, Canada; Division of Research, Kaiser Permanente Northern California, Oakland, CA, USA and Department of Health Systems Science, Kaiser Permanente Bernard J. Tyson School of Medicine, Pasadena, CA, USA; Department of Epidemiology and Biostatistics, University of California San Francisco, San Francisco, CA, USA; Mid-Atlantic Permanente Research Institute, Kaiser Permanente Mid-Atlantic Permanente Medical Group, Rockville, MD USA; Epidemiology and Population Health Program, British Columbia Centre for Excellence in HIV/AIDS, Vancouver, British Columbia, Canada; Department of Medicine, University of California San Diego, San Diego, CA, USA; Departments of Medicine and Biostatistics, Vanderbilt University School of Medicine, Nashville, TN, USA; Department of Ophthalmology, Wilmer Eye Institute, Johns Hopkins University School of Medicine, Baltimore, MD, USA; Department of Social Medicine, University of North Carolina, Chapel Hill, NC, USA; Division of Allergy and Infectious Diseases, Departments of Medicine and Epidemiology, University of Washington, Seattle, Washington, USA; Department of Behavioral, Social, and Health Education Sciences, Emory University Rollins School of Public Health, Atlanta, GA, USA; Division of Worldwide Research and Development, Pfizer Inc., New York, NY, USA; Division of Infectious Disease, Emory School of Medicine, Atlanta, GA, USA; Atlanta Veterans Affairs Health Care System, Decatur, GA, USA; Department of Medicine, University of North Carolina at Chapel Hill, Chapel Hill, NC, USA; Vanderbilt Tuberculosis Center, Vanderbilt University School of Medicine, Nashville, Tennessee, USA; Division of Infectious Diseases, Department of Medicine, Vanderbilt University School of Medicine, Nashville, Tennessee, USA; Division of General Internal Medicine, Department of Medicine, Johns Hopkins School of Medicine, Baltimore, MD, USA

**Keywords:** HIV, aging, comorbidities, multimorbidity, agent-based simulation model

## Abstract

**Importance:** Estimating the medical complexity of people aging with HIV can inform clinical programs and policy to meet future healthcare needs.

**Objective:** To project the prevalence of comorbidities and multimorbidity among people with HIV (PWH) using antiretroviral therapy (ART) in the US through 2030.

**Design:** Agent-based simulation model

**Setting:** HIV clinics in the United States in the recent past (2020) and near future (2030)

**Participants:** In 2020, 674,531 PWH were using ART; 9% were men and 4% women with history of injection drug use; 60% were men who have sex with men (MSM); 8% were heterosexual men and 19% heterosexual women; 44% were non-Hispanic Black/African American (Black); 32% were non-Hispanic White (White); and 23% were Hispanic.

**Exposure(s):** Demographic and HIV acquisition risk subgroups

**Main Outcomes and Measures:** Projected prevalence of anxiety, depression, stage ≥3 chronic kidney disease (CKD), dyslipidemia, diabetes, hypertension, cancer, end-stage liver disease (ESLD), myocardial infarction (MI), and multimorbidity (≥2 mental or physical comorbidities, other than HIV).

**Results:** We projected 914,738 PWH using ART in the US in 2030. Multimorbidity increased from 58% in 2020 to 63% in 2030. The prevalence of depression and/or anxiety was high and increased from 60% in 2020 to 64% in 2030. Hypertension and dyslipidemia decreased, diabetes and CKD increased, MI increased steeply, but there was little change in cancer and ESLD. Among Black women with history of injection drug use (oldest demographic subgroup in 2030), CKD, anxiety, hypertension, and depression were most prevalent and 93% were multimorbid. Among Black MSM (youngest demographic subgroup in 2030), depression was highly prevalent, followed by hypertension and 48% were multimorbid. Comparatively, 67% of White MSM were multimorbid in 2030 (median age in 2030=59 years) and anxiety, depression, dyslipidemia, CKD, and hypertension were highly prevalent.

**Conclusion and relevance:** The distribution of multimorbidity will continue to differ by race/ethnicity, gender, and HIV acquisition risk subgroups, and be influenced by age and risk factor distributions that reflect the impact of social disparities of the health on women, people of color, and people who use drugs. HIV clinical care models and funding are urgently required to meet the healthcare needs of people with HIV in the next decade.

**KEY POINTS:** 

**Question:** How will the prevalence of multimorbidity change among people with HIV (PWH) using antiretroviral therapy in the US from 2020 to 2030?

**Findings:** In this agent-based simulation study using data from the NA-ACCORD and the CDC, multimorbidity (≥2 mental/physical comorbidities other than HIV) will increase from 58% in 2020 to 63% in 2030. The composition of comorbidities among multimorbid PWH vary by race/ethnicity, gender, and HIV acquisition risk group.

**Meaning:** HIV clinical programs and policy makers must act now to identify resources and care models to meet the increasingly complex medical needs of PWH over time, particularly mental healthcare needs.

## INTRODUCTION

People with HIV (PWH) survive to older ages with effective antiretroviral treatment but have fewer comorbidity-free life years compared to people without HIV.^1^ Projecting the magnitude of mental and physical comorbidity, and multimorbidity, is critical for quantifying the medical complexity of PWH and allocating sufficient resources to systems and programs caring for people aging with HIV.

Many risk factors for comorbidities in the general population have a higher prevalence among people with HIV, including tobacco and substance use, higher body mass index (BMI), and hepatitis C virus (HCV) co-infection.^2–5^ The increased risk profile results in a higher prevalence of major depressive disorder (depression), generalized anxiety disorder (anxiety), hypertension, dyslipidemia, chronic kidney disease (CKD), diabetes, liver disease, cancer, and cardiovascular disease (CVD) in people with (vs. without) HIV.^6–15^ Other factors contributing to the increased burden of comorbidities in people with HIV include: 1) HIV-induced chronic immune activation and inflammation,^16,17^ 2) specific antiretroviral drugs and regimens,^18,19^ and 3) social determinants of health (SDoH).^20,21^ The disproportionate burdens of comorbidities and subsequent multimorbidity (i.e., ≥2 comorbidities not including HIV) pose persistent challenges in ensuring adequate healthcare for people with HIV.^22,23^ Multimorbidity is a function of the comorbidities included in the definition and estimates among PWH in the US have a wide range.^24–26^

Disparities persist in the comorbidity and multimorbidity among PWH. People of color with HIV have greater comorbidity burden, which is accentuated in women of color.^27,28^ Age-stratified incidence rates and risk of hypertension, diabetes, CKD, myocardial infarction (MI), and certain cancers are particularly high among non-Hispanic Black/African American (Black/AA) PWH, as are risk factors common to many comorbidities, including smoking, obesity, and SDoH.^11,29–31^ People with injection drug use as their HIV acquisition risk factor have the greatest burden of comorbidity and multimorbidity among PWH.^32,33^ To better assess future healthcare needs and address health inequities among PWH, clinical and policy decision-makers need projections of future multimorbidity burden in PWH subgroups.^34,35^ The objective of this study is to project the prevalence of future comorbidities and multimorbidity among PWH using ART in the US through the year 2030, overall and within 15 demographic subgroups.

## METHODS

The <u>P</u>roj<u>E</u>cting <u>A</u>ge, multimo<u>R</u>bidity, and po<u>L</u>ypharmacy (PEARL) model is an agent-based computer simulation model of PWH who have initiated ART in the US, including those who disengage from HIV care (**Figure 1**). The simulated population is comprised of fifteen subgroups: (1 & 2) men and women with history of injection drug use as an HIV acquisition risk factor, including those who have injection drug use and any additional HIV acquisition risk category specified (MWID and WWID respectively); (3) men who have sex with men (MSM); and (4 & 5) heterosexual men and women. These five groups were further stratified into non-Hispanic White, non-Hispanic Black/AA, and Hispanic. Race and ethnicity defined subgroups that compose the PEARL simulation model due to the disproportionate burden of HIV by race and ethnicity in the US. Asian and American Indian/Alaskan Native participants are not included in this analysis due to limited input parameters and functions within the 5 HIV acquisition risk groups.

**Figure 1:**
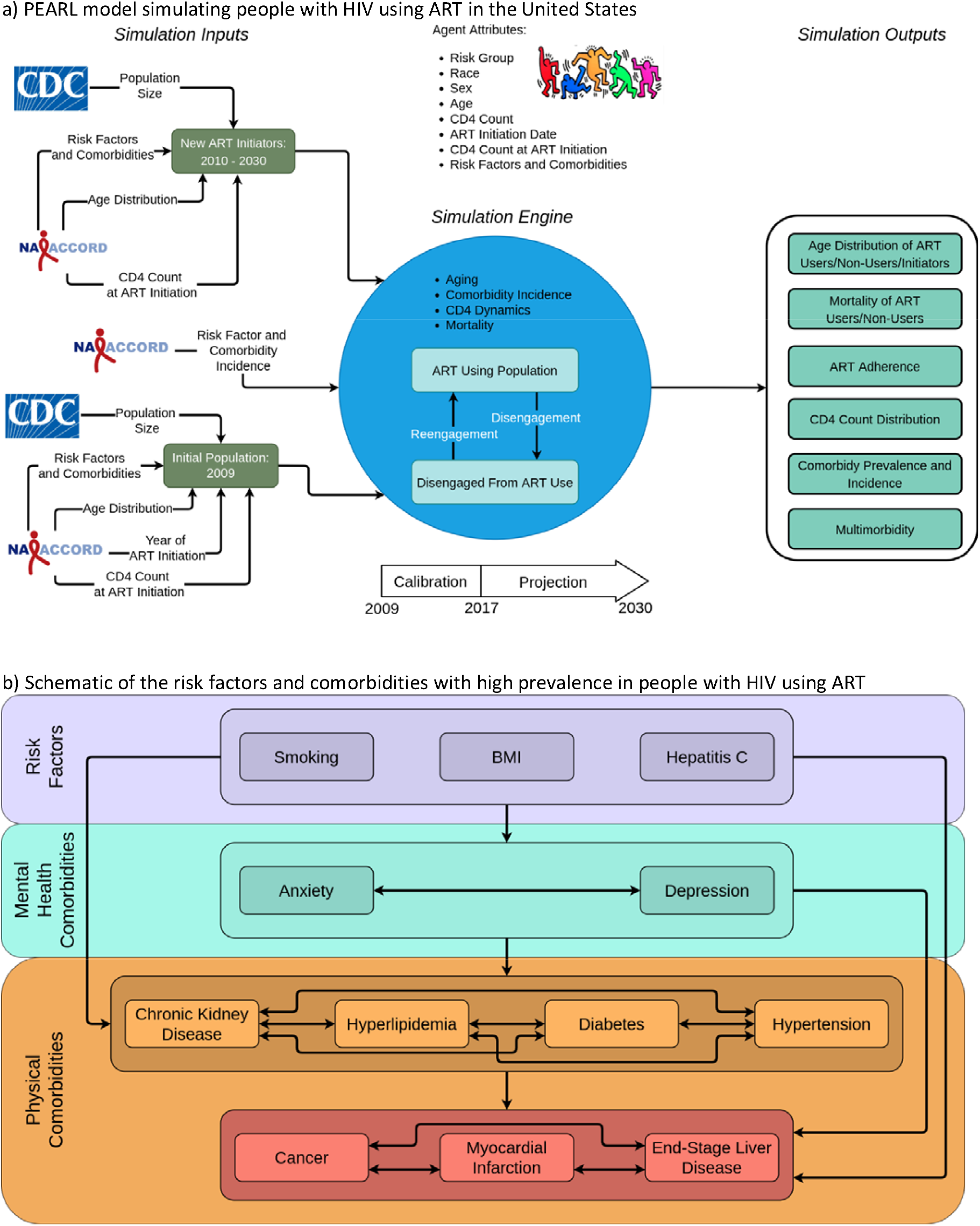
Schematic representation of a) the PEARL model and b) the risk factors and comorbidities with high prevalence in people with HIV using ART Footnotes: ART=antiretroviral therapy (HIV treatment) CD4=CD4 T-lymphocyte cell count Details on the mathematical functions represented by the arrows between the risk factors and mental and physical comorbidities can be found at PEARLHIVmodel.org.

Briefly, agents enter the model at ART initiation, and experience aging, disengagement and re-engagement in care, changing CD4 counts, and mortality via mathematical functions and parameters that are informed by observed data from the North American AIDS Cohort Collaboration on Research and Design (NA-ACCORD) and Centers for Disease Control and Prevention’s (CDC) HIV surveillance reports.^36,37^ All mathematical functions and parameters in PEARL are estimated separately for each of the 15 subgroups. Further methodological details regarding the model structure, parameterizations, standards for collapsing subgroups to ensure adequate sample size, and estimated functions and values are available at PEARLHIVMODEL.org/method_details.html.^36,38^

### Comorbidities parameterization

First, we characterized the prevalence of clinical risk factors that are highly prevalent in PWH, linked to numerous comorbidities, and measured in the NA-ACCORD (namely smoking status, obesity, and HCV co-infection status) for the simulated population in 2009 and those starting ART over time. Smoking and HCV co-infection status were determined at ART initiation (i.e., the simulated person’s entry to the model) and were time-fixed. Obesity (BMI ≥30 kg/m^2^) status was determined at ART initiation and at 24 months after ART initiation. Next, we estimated the prevalence (at ART initiation) and incidence (in the years after ART initiation) of highly prevalent comorbidities in PWH for simulated persons within the 15 subgroups via mathematical functions derived from observed NA-ACCORD data from 2009-2017; NA-ACCORD definitions for risk factors and comorbidities using electronic health record data are available in **Supplement Table S1**. The prevalence of having depression, anxiety, stage ≥3 CKD, dyslipidemia, type 2 diabetes, hypertension, cancer (all types), myocardial infarction (MI), and ESLD (at or prior to ART initiation) was estimated at the time the simulated person initiated ART (**Figure 1**).^24,30,39–41^ Incidence of each comorbidity was estimated as a function of age, CD4 at ART initiation (cells/μL), time since ART initiation, disengagement from care, change in BMI in the first 2 years after ART initiation, BMI 2 years after ART initiation, smoking, HCV co-infection, and the presence of the other comorbidities for simulated persons in the years after ART initiation (**Supplement Table S2**). Finally, mortality was estimated for simulated persons as a function of individual-level attributes, existing risk factors, and present comorbidities, and was estimated separately for those (1) engaged and (2) disengaged from care (≥2 years without CD4 or HIV RNA measurement) and in each of the 15 sub-groups using observed NA-ACCORD data from 2009-2017 (**Supplement Table S3**).

### Primary outcomes

While prior multimorbidity studies in PWH were restricted to physical comorbidities, we chose the following non-mutually exclusive definitions to produce findings comparable to prior studies and to expand the scope of multimorbidity to include the presence of mental health conditions:

a. physical multimorbidity (≥2 physical comorbidities)
b. depression and/or anxiety diagnosis (mental comorbidities)
c. mental *or* physical multimorbidity (≥2 mental *or* physical comorbidities), and
d. mental comorbidity *and* physical multimorbidity (≥1 mental health comorbidity *and* ≥2 physical comorbidities).

Given the association of age with comorbidities of interest, and heterogeneities in projected age distributions by subgroup, we report the projected median age in 2020 and 2030 within the 15 subgroups, as well as, the projected prevalence of each mental and physical comorbidity, overall and within subgroups. To navigate temporal trends, we also report the absolute percentage point change [ppc] in the prevalence of each mental and physical comorbidity from 2020 to 2030 within the 15 subgroups.

Simulations were replicated for 200 runs and projected outcomes are presented as median values and 95% uncertainty ranges (95%UR).

### Validation

To ensure the model’s ability to project realistic comorbidity patterns, we compared the observed annual incidence and prevalence of each comorbidity from observed NA-ACCORD data with respective projections from PEARL between 2009-2017 (calibration period) within each subgroup. Within each subgroup, we flagged the comorbidities with <75% of projections within the larger of two ranges surrounding the observed mean incidence and prevalence: (a) +/-5% of the observed mean incidence and prevalence or (b) the observed 95% confidence interval (**Supplement Figures S1-S2**).

### Robustness of projected comorbidity incidence

In the PEARL model, projected multimorbidity is a function of the comorbidities that arise from estimated probabilities for the incidence of each comorbidity within each subgroup. To assess the influence of the estimated probabilities for the incidence of each comorbidity on projected physical multimorbidity, we increased and decreased the probabilities for the incidence of each comorbidity by 25% for each simulated person in the model (i.e., the increase and decrease scenarios). We estimated the relative difference in the projected physical multimorbidity in 2030 in each scenario compared to the baseline scenario (i.e., no change in the estimated probabilities for the incidence of each comorbidity). We chose physical multimorbidity as the outcome for the robustness checks to allow for comparability to other estimates of physical multimorbidity in PWH in the US.^24–26^ The robustness of the prevalence and mortality estimates was similar, and results can be found at <u>PEARLHIVMODEL.org/method_details.html</u>.

## RESULTS

Using the PEARL model, we simulated a median population of 674,531 [95%UR 667,269–680,808] PWH using ART in 2020 in the US, of whom 52% were ≥50 years, 11% were age ≥65 years, 32% White, 44% Black/AA, 23% Hispanic, 60% MSM, 19% heterosexual women, 9% MWID, 8% heterosexual men, and 4% WWID, (**Table 1, Supplement Table S4**).

**Table 1:** Characteristics of the PEARL-simulated agents using ART in 2010, 2020 and 2030

| Characteristics | 2010 |  | 2020 (Projected) |  | 2030 (Projected) |  |
| --- | --- | --- | --- | --- | --- | --- |
|  | PEARL<sup>a</sup> |  | PEARL<sup>a</sup> |  | PEARL<sup>a</sup> |  |
|  | N= | 395,050 | N = | 674,531 | N = | 914,738 |
|  | n | %<sup>b</sup> | n | %<sup>b</sup> | n | %<sup>b</sup> |
| **Age (in years)** |  |  |  |  |  |  |
| <20 | 695 | 0% | 699 | 0% | 1,210 | 0% |
| 20-24 | 8,434 | 2% | 11,452 | 2% | 12,238 | 1% |
| 25-29 | 20,049 | 5% | 40,556 | 6% | 40,794 | 4% |
| 30-34 | 30,854 | 8% | 66,908 | 10% | 78,542 | 9% |
| 35-39 | 47,940 | 12% | 63,128 | 9% | 101,300 | 11% |
| 40-44 | 67,573 | 17% | 61,290 | 9% | 102,383 | 11% |
| 45-49 | 78,964 | 20% | 80,046 | 12% | 85,847 | 9% |
| 50-54 | 66,239 | 17% | 98,716 | 15% | 81,764 | 9% |
| 55-59 | 42,424 | 11% | 101,289 | 15% | 95,226 | 10% |
| 60-64 | 20,650 | 5% | 76,974 | 11% | 103,896 | 11% |
| 65-69 | 7,820 | 2% | 44,785 | 7% | 95,728 | 10% |
| 70-74 | 2,472 | 1% | 19,800 | 3% | 65,606 | 7% |
| ≥75 | 916 | 0% | 8,969 | 1% | 48,778 | 5% |
| **Male sex at birth** | 289,993 | 73% | 518,620 | 77% | 714,764 | 78% |
| **Race** |  |  |  |  |  |  |
| White | 145,688 | 37% | 218,290 | 32% | 261,230 | 29% |
| Black/AA | 167,442 | 42% | 298,157 | 44% | 395,170 | 43% |
| Hispanic | 81,908 | 21% | 157,923 | 23% | 256,343 | 28% |
| **Sub-groups, n % (median age) [IQR]** |  |  |  |  |  |  |
| MSM | 210,646 | 53% | 406,590 | 60% | 587,148 | 64% |
|  | (45) | [37, 51] | (48) | [35, 57] | (47) | [37, 61] |
| White MSM | 102,916 | 26% | 156,942 | 23% | 180,206 | 20% |
|  | (47) | [41, 53] | (54) | [44, 60] | (59) | [46, 67] |
| Black/AA MSM | 63,098 | 16% | 144,577 | 21% | 218,374 | 24% |
|  | (42) | [33, 49] | (41) | [32, 53] | (42) | [35, 56] |
| Hispanic MSM | 44,620 | 11% | 104,670 | 16% | 188,080 | 21% |
|  | (41) | [35, 47] | (44) | [34, 52] | (43) | [36, 56] |
| Men who injected drugs (MWID)<sup>c</sup> | 49,860 | 13% | 59,818 | 9% | 65,672 | 7% |
|  | (52) | [47, 57] | (58) | [49, 64] | (58) | [39, 69] |
| White MWID | 17,539 | 4% | 23,652 | 4% | 29,460 | 3% |
|  | (50) | [44, 55] | (56) | [47, 62] | (57) | [40, 67] |
| Black/AA MWID | 20,536 | 5% | 21,163 | 3% | 17,748 | 2% |
|  | (54) | [50, 58] | (61) | [55, 66] | (63) | [34, 72] |
| Hispanic MWID | 11,783 | 3% | 15,000 | 2% | 18,506 | 2% |
|  | (52) | [45, 57] | (57) | [47, 64] | (56) | [41, 69] |
| Women who injected drugs (WWID) | 25,817 | 7% | 27,804 | 4% | 31,149 | 3% |
|  | (49) | [43, 54] | (57) | [50, 63] | (62) | [51, 70] |
| White WWID | 7,463 | 2% | 9,592 | 1% | 13,401 | 1% |
|  | (46) | [39, 51] | (53) | [44, 59] | (56) | [47, 65] |
| Black/AA WWID | 14,570 | 4% | 14,222 | 2% | 12,936 | 1% |
|  | (50) | [45, 54] | (59) | [53, 64] | (66) | [58, 72] |
| Hispanic WWID | 3,786 | 1% | 3,994 | 1% | 4,820 | 1% |
|  | (49) | [44, 54] | (58) | [51, 63] | (63) | [53, 70] |
| Heterosexual men | 29,505 | 7% | 52,364 | 8% | 62,334 | 7% |
|  | (47) | [40, 53] | (53) | [44, 60] | (58) | [47, 67] |
| White heterosexual men | 3,486 | 1% | 6,840 | 1% | 9,369 | 1% |
|  | (49) | [43, 55] | (55) | [46, 62] | (61) | [48, 69] |
| Black/AA heterosexual men | 19,166 | 5% | 34,418 | 5% | 39,848 | 4% |
|  | (47) | [41, 53] | (53) | [45, 60] | (58) | [46, 66] |
| Hispanic heterosexual men | 6,852 | 2% | 11,108 | 2% | 12,998 | 1% |
|  | (44) | [37, 52] | (52) | [43, 60] | (58) | [49, 67] |
| Heterosexual women | 79,226 | 20% | 128,017 | 19% | 167,442 | 18% |
|  | (44) | [36, 51] | (51) | [42, 59] | (57) | [47, 65] |
| White heterosexual women | 14,288 | 4% | 21,160 | 3% | 28,882 | 3% |
|  | (45) | [38, 52] | (52) | [44, 59] | (59) | [50, 67] |
| Black/AA heterosexual women | 50,071 | 13% | 83,713 | 12% | 106,270 | 12% |
|  | (43) | [36, 51] | (50) | [42, 58] | (56) | [47, 65] |
| Hispanic heterosexual women | 14,864 | 4% | 23,094 | 3% | 31,992 | 3% |
|  | (43) | [36, 51] | (50) | [41, 59] | (55) | [42, 66] |
| **Mental Comorbidities** |  |  |  |  |  |  |
| Anxiety | 95,047 | 24% | 246,192 | 36% | 428,545 | 47% |
| Depression | 157,590 | 40% | 317,676 | 47% | 445,518 | 49% |
| Anxiety and/or depression | 209,578 | 53% | 405,332 | 60% | 589,178 | 64% |
| **Physical Comorbidities** |  |  |  |  |  |  |
| Stage ≥3 Chronic Kidney Disease | 39,678 | 10% | 111,940 | 17% | 227,423 | 25% |
| Dyslipidemia | 119,246 | 30% | 184,860 | 27% | 234,910 | 26% |
| Diabetes | 45,510 | 12% | 105,239 | 16% | 219,064 | 24% |
| Hypertension | 145,030 | 37% | 239,688 | 36% | 287,116 | 31% |
| Cancer | 37,688 | 10% | 75,944 | 11% | 104,010 | 11% |
| End-Stage Liver Disease | 4,704 | 1% | 9,062 | 1% | 13,334 | 1% |
| Myocardial Infarction | 6,276 | 2% | 21,934 | 3% | 72,294 | 8% |
| **Physical Multimorbidity** |  |  |  |  |  |  |
| No physical comorbidities | 126,168 | 32% | 237,779 | 35% | 314,013 | 34% |
| 1 physical comorbidity | 164,476 | 42% | 226,742 | 34% | 275,250 | 30% |
| ≥2 physical comorbidities | 104,420 | 26% | 209,865 | 31% | 325,266 | 36% |
| **Mental and physical multimorbidity** |  |  |  |  |  |  |
| ≥2 mental or physical comorbidities | 205,456 | 52% | 393,150 | 58% | 579,564 | 63% |
| ≥1 mental and ≥2 physical comorbidities | 56,352 | 14% | 139,335 | 21% | 232,310 | 25% |
| **ART status** |  |  |  |  |  |  |
| PWH using ART | 395,050 |  | 674,531 |  | 914,738 |  |
| ART initiators | 25,104 |  | 32,963 |  | 33,324 |  |
| Disengaged from ART use<sup>d</sup> | 42,290 |  | 41,884 |  | 33,208 |  |

AA=African American
ART=antiretroviral therapy for HIV treatment
PEARL=Projecting Age, Multimorbidity, and Polypharmacy in Adults with HIV
PWH=people with HIV
MSM=men who have sex with men
<sup>a</sup>Values represent the median for each simulated outcome across 200 random simulation replications (See **Supplement Table S4** for 95% interquartile ranges).
<sup>b</sup>Percentages in this table are calculated using the median numerator (n) and median denominator (N) from 200 replications of the PEARL model; for each characteristic, percentages will sum to 100%.
<sup>c</sup>MSM who also have MWID as their HIV acquisition risk factor were included in the MWID HIV acquisition risk group.
<sup>d</sup>PEARL projections of PWH using ART do not include 41,884 and 33,208 people who initiated ART but were disengaged from care and not using ART in 2020 and 2030 (respectively) and people of race and ethnicities other than non-Hispanic White, non-Hispanic Black/AA, and Hispanic.

In 2020, the prevalence of anxiety and depression were 36% and 47%, respectively, with 23% of simulated agents having both diagnoses. Applying our non-mutually exclusive definitions of multimorbidity among ART users, 31% were physically multimorbid, 58% had mental *or* physical multimorbidity, and 21% had mental comorbidity *and* physical multimorbid. Of the physical conditions included in the model, hypertension was the most prevalent (36%), followed by dyslipidemia (27%), CKD (17%), diabetes (16%) and cancer (11%); MI and ESLD had a prevalence of <5%.

In 2030, the projected population of PWH using ART increased by almost a quarter of a million people (36% increase from 2020), and the proportion ≥65 years more than doubled to 23%; race/ethnicity and HIV acquisition risk group distributions changed by <5 percentage points from 2020 to 2030. The prevalence of hypertension and dyslipidemia decreased slightly by 5 and 1 percentage points (respectively) 2020 to 2030, the prevalence of CKD and diabetes increased by 8 percentage points, the prevalence of MI increased by 5 percentage points, and the prevalence of ESLD remained constant.

Validity of the mental and physical comorbidity projections was confirmed by comparing the observed comorbidity incidence and prevalence of PWH using ART from 2010–2017 in NA-ACCORD with the simulated outcomes, suggesting no significant pattern of bias in projections from the model among 15 subgroups over time (**Supplement Figures S1, S2**).

### Projections of multimorbidity

Overall, the prevalence of physical multimorbidity increased from 2020 to 2030 (**Figure 2a**), and in each subgroup (**Figure 2b**). The number of Black/AA MWID and WWID using ART did not decreased from 2020 to 2030, but the physical multimorbidity prevalence was high and increased throughout the decade (**Table 1, Supplement Table S5**). Black/AA MSM and heterosexual women had patterns of increasing number of ART users and physical comorbidity prevalence that were similar to the White and Hispanic subgroups.

**Figure 2:**
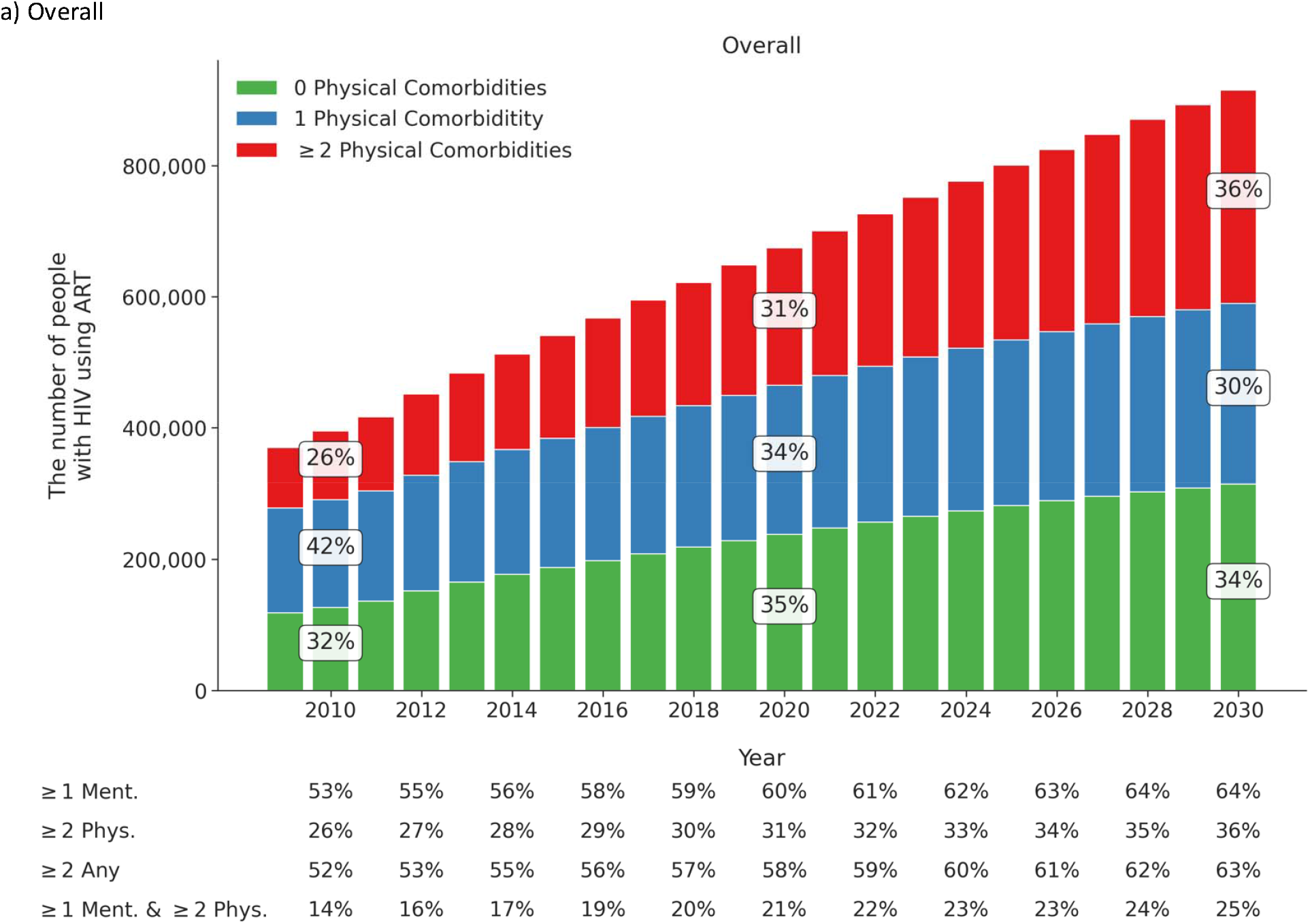
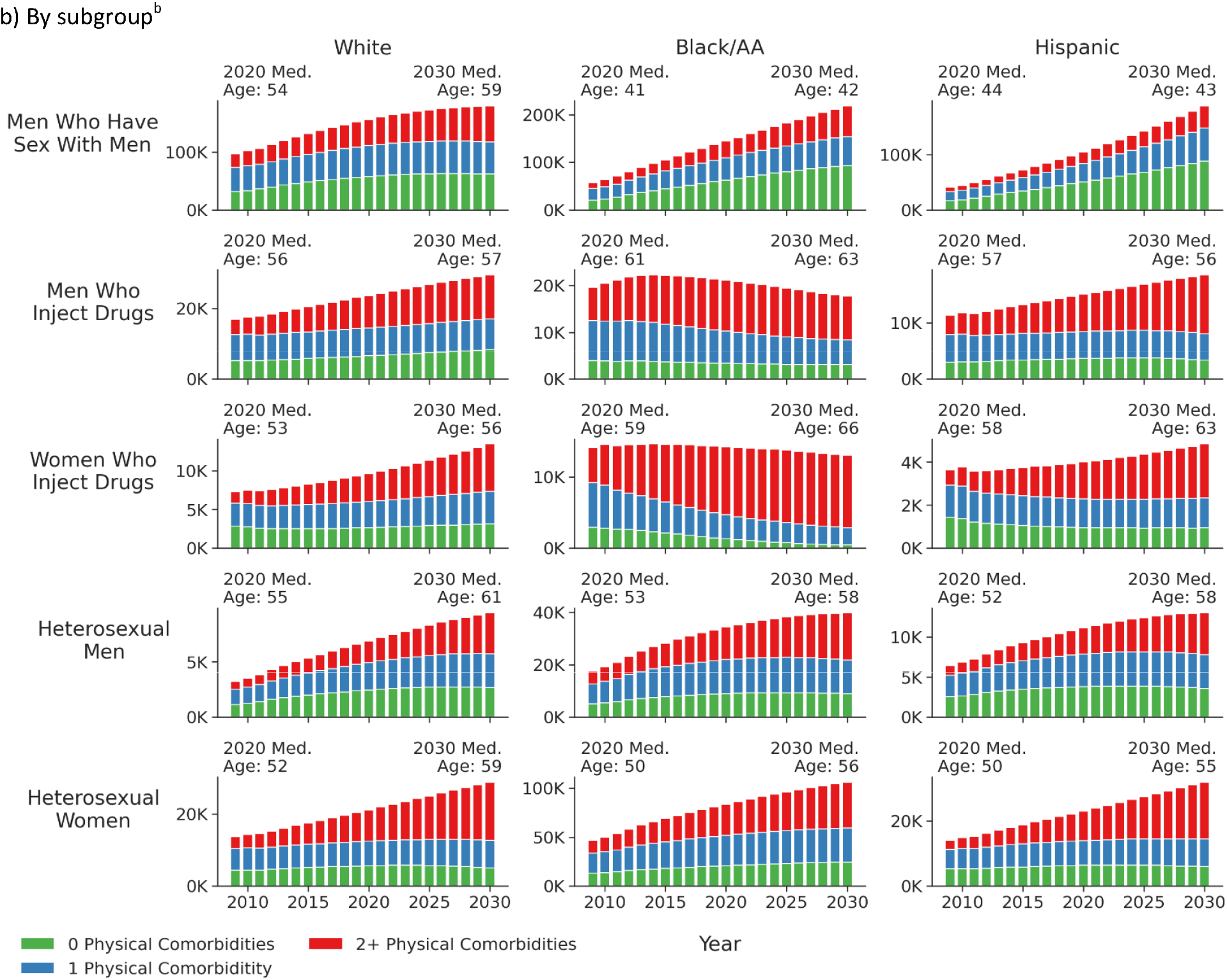
Projected^a^ number of PWH using ART in the US and projected prevalence of mental and physical comorbidities and multimorbidity among PWH using ART in the US a) overall and b) by subgroup^b^ Footnotes: ≥1 Ment. = anxiety and/or depression (i.e., ≥1 of the mental comorbidities included) ≥2 Phys. = physical multimorbidity, defined as ≥2 physical comorbidities ≥2 Any = mental *or* physical multimorbidity, defined as ≥2 physical or mental comorbidities ≥1 Ment. & 2 Phys. = mental comorbidity *and* physical multimorbidity, defined as ≥1 mental comorbidity and ≥2 physical comorbidities ^a^Although these estimates are all PEARL projections, 2010 was during the calibration period (where observed NA-ACCORD data were available to inform the estimates) and 2020 and 2030 were projection periods (without observed NA-ACCORD data). ^b^Note that the y axes are different across the subgroups to allow visualization of the number of comorbidities within each year

Overall, the prevalence of depression and/or anxiety was higher than any physical comorbidity in 2020 (60%) and in 2030 (65%, **Figure 2a**). The prevalence with depression and/or anxiety in 2030 was greatest in Hispanic MWID (**Supplement Table S5**). The increase in physical multimorbidity burden from 2020 to 2030 was greatest in Hispanic heterosexual women (**Supplement Table S5**). The proportion of PWH using ART with physical *or* mental multimorbidity was 58% in 2020 and 63% in 2030 (**Figure 2a**). The prevalence with mental comorbidities *and* physical multimorbidity increased from 21% in 2020 to 25% in 2030 (**Figure 2a**).

### Projections of each comorbidity

Among all PWH using ART from 2020 to 2030 projected by the PEARL model, depression had the highest prevalence over the 10-year period (49% [95%UR 48-49%] in 2030), and anxiety increased from 37% [95%UR 36-37%] in 2020 to 47% [95%UR 46-48%] in 2030 (**Figure 3a, Supplement Table S6**). Hypertension and dyslipidemia prevalence declined slightly (<4 ppc), and diabetes and CKD increased (>7 ppc), over this decade; prevalence for these four physical comorbidities ranged from 24% [95%UR 24-25%] for diabetes to 31% [95%UR 31-32%] for hypertension in 2030. Cancer and ESLD had little change in prevalence; however, cancer had a higher prevalence in 2030 (11% [95%UR 11-12%]) than ESLD (2% [95%UR 1-2%]). In comparison, MI was projected to increase steeply from 3% [95%UR 3-3%] in 2020 to 8% [95%UR 8-8%] in 2030.

**Figure 3:**
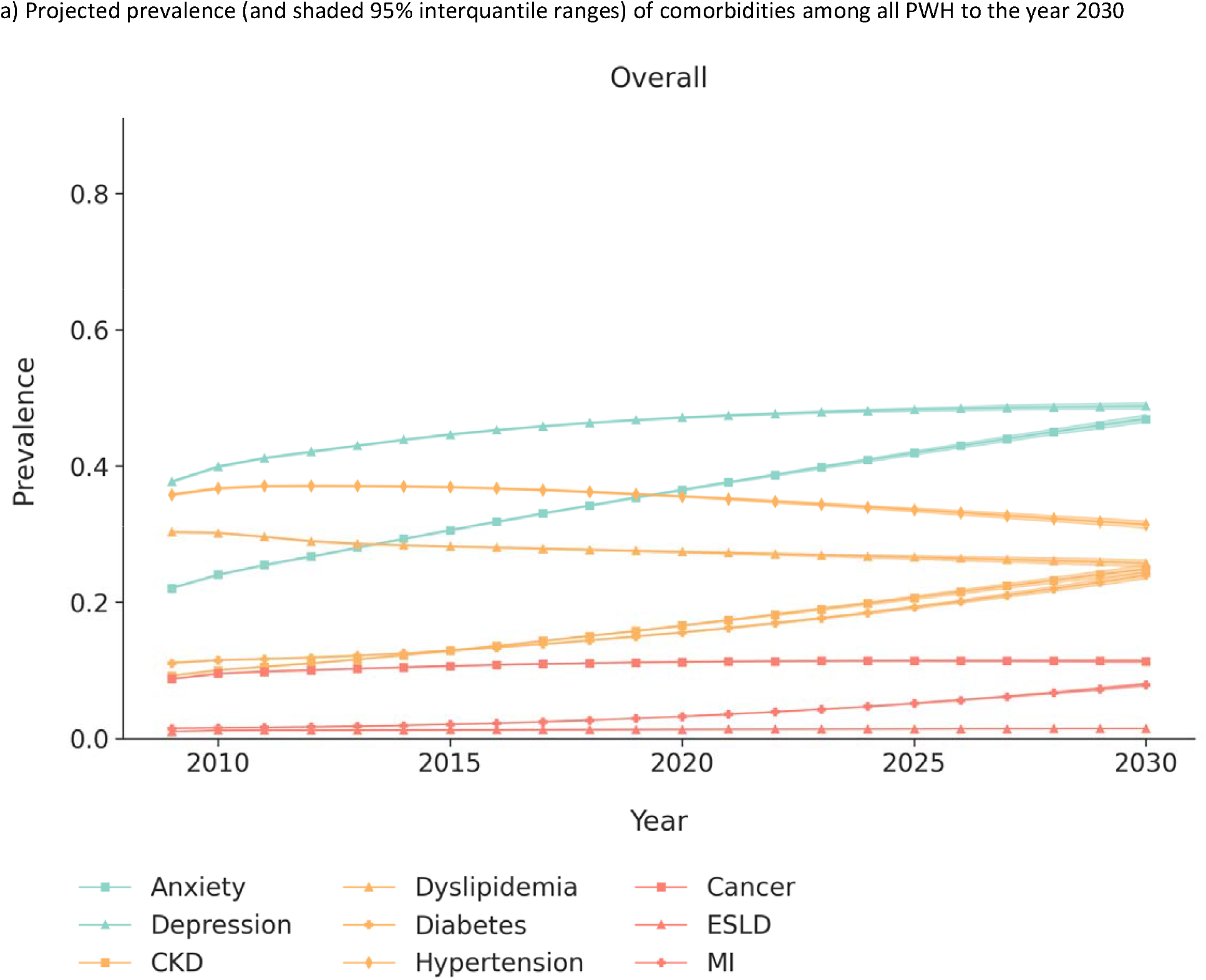
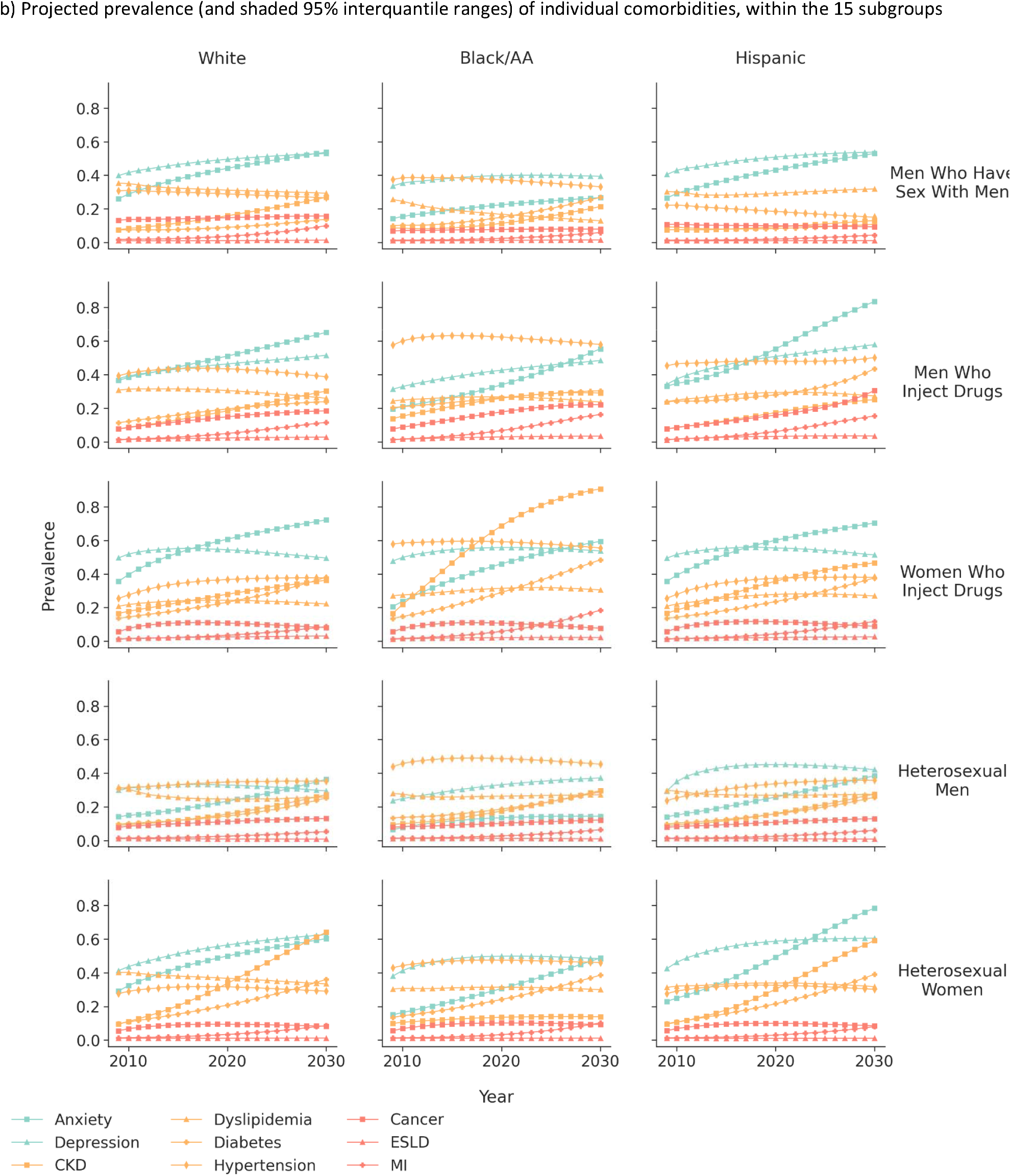
Projected prevalence (and shaded 95% interquantile ranges) of individual comorbidities among PWH using ART a) overall, b) among the 15 subgroups, c) among the subgroup with the oldest median age in 2030, and c) among the subgroup with the youngest median age in 2030 Footnotes: CKD=stage ≥3 chronic kidney disease ESLD=end-stage renal disease MI=myocardial infarction The 95% credibility interval is estimated as the 2.5% and 97.5% range of results from running the simulation 200 times.

There were differences in the change in comorbidity burdens from 2020 to 2030 across the 15 subgroups (**Figure 3b**). The subgroup with the oldest median age (66 [95%UR 65-68] years) in 2030 was Black/AA WWID, and Black MSM had the youngest median age in 2030 (42 [95%UR 41-43] years). Among Black/AA WWID, CKD, anxiety, hypertension, and depression were the most prevalent comorbidities in 2030, and diabetes and MI had lower but increasing prevalence from 2020-2030 (**Figure 3c**). Hyperlipidemia, cancer, and ESLD had little change in prevalence and declined slightly at the end of the study period. Among Black MSM, depression had the highest prevalence in 2030 followed by hypertension which decreased from 2020 to 2030 (**Figure 3d**). Anxiety, diabetes, CKD, and MI all increased during the two decades, whereas dyslipidemia prevalence decreased and cancer and ESLD did not change. Larger depictions of comorbidity prevalence estimates in each subgroup are available in **Supplement Figures S3a-o**.

The ppc from 2020 to 2030 in the prevalence of each physical and mental comorbidity is shown in **Figure 4**, stratified by the 15 subgroups of PWH using ART. The prevalence of anxiety and MI increased in all 15 subgroups and overall, by 10 ppc and 5 ppc (respectively). Diabetes and CKD prevalence increased by 8 ppc overall, and an increase was experienced in each subgroup for both comorbidities. Conversely, dyslipidemia and hypertension decreased in all but 4 and 5 subgroups and by 2 ppc and 4 ppc overall, respectively. There was <2 ppc change for depression, cancer, and ESLD.

**Figure 4:**
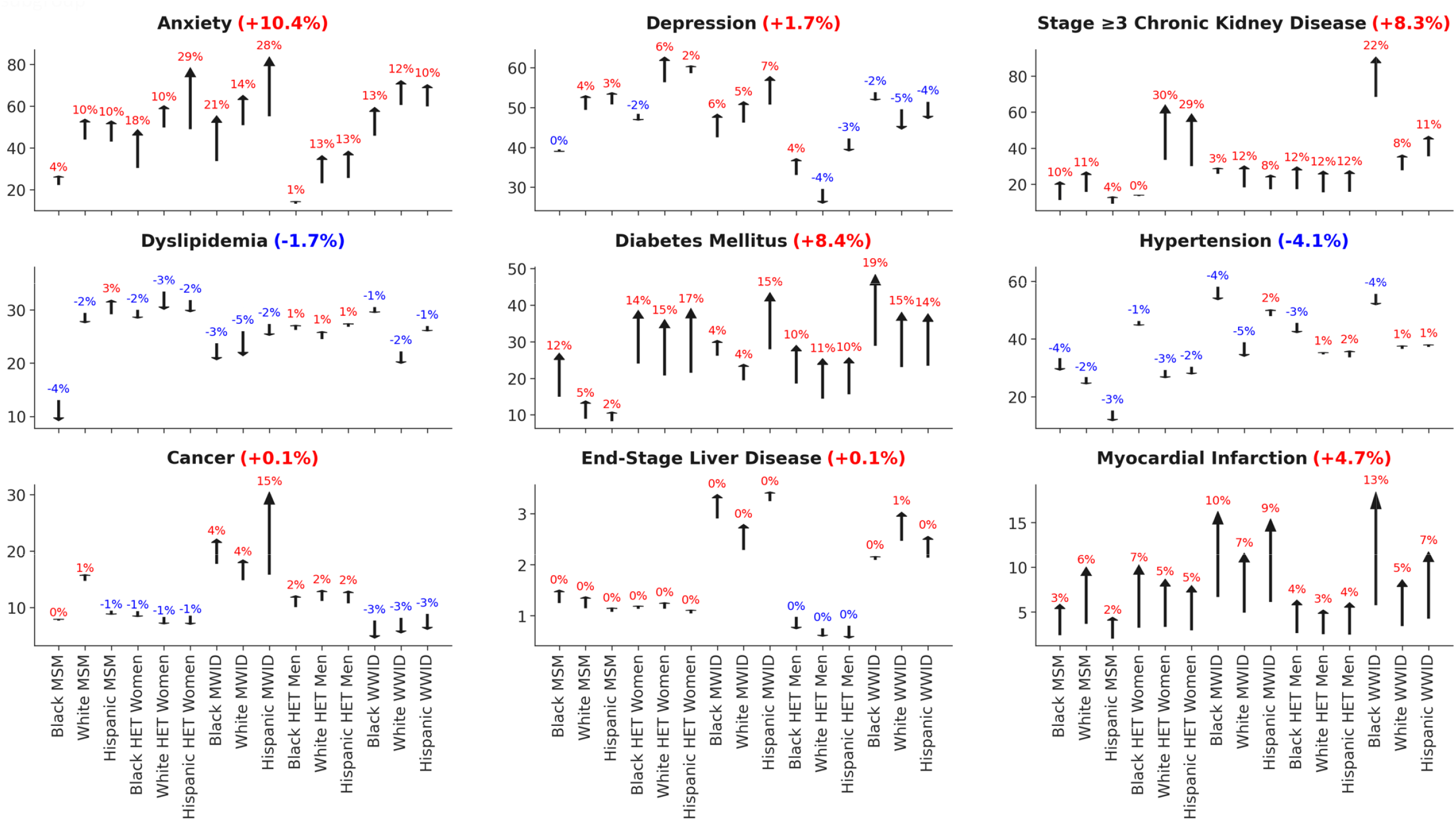
Projected absolute percentage point change (blue=decrease, red=increase) in the prevalence of individual comorbidities from 2020 to 2030, by subgroup Footnotes: ^b^The y axes are different across the subgroups to allow visualization of the number of comorbidities within each year.

**Figure 5:**
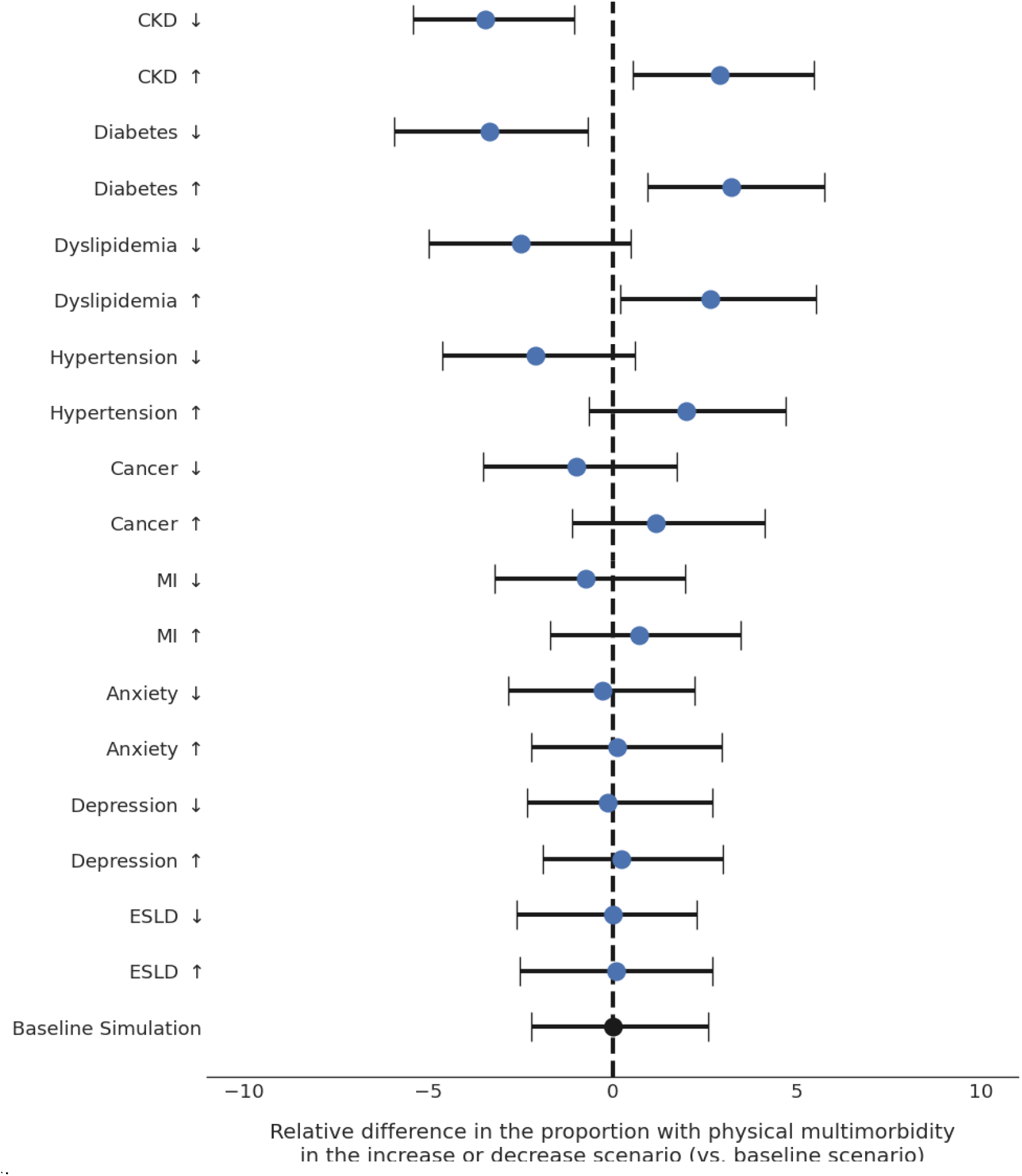
The relative difference of the proportion with physical multimorbidity in 2030 [outcome] comparing scenarios in which comorbidity incidence was decreased by 25% (down arrow scenario) and increased by 25% (up arrow scenario) to assess the influence of estimated probabilities on prevalence estimates. Relative difference when probability of the incidence of a comorbidity was decreased by 25% and increased by 25%, compared to the baseline scenario (no modification to the probability of the incidence of a comorbidity) Footnotes:

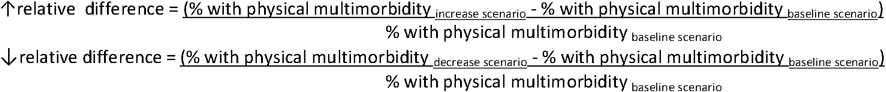

Physical multimorbidity = ≥2 physical comorbidities

### Robustness of each projected comorbidity incidence

The relative differences from analyzing the effect of decreasing or increase increasing comorbidity incidence probability (vs. baseline scenario) showed a <5% relative difference in the projected prevalence of physical multimorbidity, demonstrating the robustness of this estimate of projected multimorbidity to the variability in the estimated incidence of each comorbidity.

## DISCUSSION

We project an increasing prevalence of multimorbidity in PWH using ART in the US through the year 2030, with different compositions of contributing comorbidities by race/ethnicity, gender, and HIV acquisition risk. Among the comorbidities included in the PEARL model, the projections suggest the greatest contributors to multimorbidity are two common mental health diagnoses that occur throughout the lifespan: depression and anxiety. The prevalence of these mental comorbidities was projected to increase in nearly all the subgroups by the year 2030; anxiety had the steepest increase. The prevalence of mental *or* physical multimorbidity is projected to be 63% and 1 in 4 PWH using ART will have mental comorbidity *and* physical multimorbidity by 2030; these estimates are a lower limit due to the inclusion of only 2 mental and 7 physical comorbidities in the PEARL model. HIV clinical program and policy decision-makers must modify current care models and payor capabilities (e.g., the Ryan White HIV/AIDS Program funding) to meet the growing healthcare needs, in particular the mental healthcare needs, of PWH using ART. Furthermore, our findings show the most prevalent comorbidities in the next decade will differ by gender, HIV acquisition risk group, race, and ethnicity, suggesting the clinical population composition is important when adapting current HIV care models.

Our findings underscore the role of mental health comorbidities in PWH. For example, PWH are three times more likely to currently be experiencing a major depressive episode as compared to people without HIV.^42^ Not only has depression been linked to missed HIV clinical care visits, virologic failure, and all-cause mortality in PWH, but depression has also been linked to similar mechanisms of immune suppression and inflammation.^43,44^ Mental health services are allowable costs for PWH eligible for Ryan White HIV/AIDS Program support; funding the establishment of mental health services associated with HIV clinics and ensuring adequate staffing will be necessary to meet the projected burden of mental health comorbidity.

Our findings corroborate estimates of increasing multimorbidity among PWH (overall and in subgroups) in the US and provide the opportunity to project future multimorbidity and comorbidity. A study comparing multimorbidity prevalence in PWH ages 45-to-89-years-old attending one visit at a Ryan White HIV/AIDS Program clinic in 2016 vs 2006 observed an increase in multimorbidity prevalence among those of similar age and an increase in women (vs. men).^45^ A study of physical multimorbidity in the NA-ACCORD noted the common comorbidity combinations included hypercholesterolemia, hypertension, and CKD in 2009.^24^ Over the next decade, these metabolic and vascular diseases may drive the PEARL-projected increases in diabetes, CKD, and MIs. Importantly, the prevalence of hypertension and dyslipidemia are projected to stabilize, perhaps due to increases in younger, healthier adults entering HIV care through successful test-and-treat (without delay) initiatives and the “Treat All” clinical guidelines, the reduced use of protease inhibitors (which have been linked to metabolic disorders), or the greater likelihood of a plateau or decrease when prevalence is high.^18,46^ Although hypertension and dyslipidemia stabilize, the incidence of MIs are projected to increase, which is influenced by an increasing number of PWH at older ages in the next decade.^30,47^

Within each subgroup, the PEARL-estimated comorbidity combinations were influenced by the age distribution, physical and behavioral risk factors, and SDoH through the stratification by gender, HIV acquisition risk, race, and ethnicity. Subgroup stratification is essential due to the heterogeneity of PWH. For example, Black/AA WWID are projected to have the oldest age distribution in 2030, which is likely a function of gender differences in life expectancy, a survivor effect of these women who had injected drugs during the period they acquired HIV, and the national reduction in new HIV infections (which typically happen in young adults) due to injection drug use.^48,49^ Higher food insecurity, space for physical activity, and access to healthcare (influenced by structural racism persistent healthcare provider implicit bias/racism) can lead to higher rates of diabetes (estimated to be 28.9% in Black/AA WWID in 2020 in the PEARL output) and hypertension (estimated to be 59.1% prevalence in Black/AA WWID in 2020 in the PEARL output) which subsequently increases the risk of progression to renal failure, which is more prevalent in Black/AA (vs. White) individuals in the US.^50–52^ We projected CKD prevalence increased from 68.6% in 2020 to 90.7% in 2030 among Black/AA WWID. Through shared pathways of inflammatory mediators, reactive oxygen species (ROS), oxidative stress, and renin-angiotensin system [RAS] components, the 13 ppc increase in anxiety is also influencing the projected increase in CKD.^53^

These comorbidities also influence the 13 ppc increase in MI among Black/AA WWID in 2030. Implementing clinical program interventions that are tailored to Black/AA women and focused on prevention and management of diabetes, hypertension, and anxiety may prove beneficial in slowing future multimorbidity growth among Black/AA WWID.

Our study has limitations. PEARL includes nine highly prevalent comorbidities that necessitate clinical management, but it does not include arthritis, stroke, fractures, which were found to be among the top comorbidities noted in a recent UK analysis of 304 physical and mental health conditions in PWH; the PEARL-projected multimorbidity prevalence is likely an underestimate.^54^ PEARL models clinically-diagnosed conditions, which is beneficial for projecting the needed clinical care resources but does not include undiagnosed conditions. Given the higher risk of COVID in HIV-infected individuals and potential of long COVID to induce similar comorbidities, it may be that our estimates will be a lower bound to future comorbidity estimates. We did not compare the comorbidities and multimorbidity projections to similar people without HIV. Although this comparison would be useful, our goal was to provide future morbidity predictions to inform HIV clinical directors’ planning and direct HIV policy decision-making. Finally, as with all agent-based simulation models, the accuracy of the output is dependent upon the quality of the mathematical functions that compose the model. We utilized observed data from the NA-ACCORD to estimate the mathematical functions as it is the largest collaboration of PWH in the US and Canada and has similar demographics to all persons living with HIV In the US (according to the CDC’s HIV surveillance data).^55^

## CONCLUSIONS

Although the gap in life expectancy has narrowed in some subgroups of people with (compared to without) HIV, multimorbidity is common in PWH in the US and will increase in prevalence over the next decade.^48^ Robust, sustainable, multidisciplinary care models (with appropriate funding) are urgently needed to meet the medically-complex healthcare needs of PWH in the US, in particular, access to affordable mental healthcare should be a priority. Predominant comorbidities differ by subgroups of PWH, which must be considered when planning for necessary resources and adapting care models. HIV clinical program and policy decision-makers must act now to identify effective multidisciplinary care models and funding to prevent and manage comorbidities and multimorbidity among the growing population of PWH using ART in the US.

## Data Availability

All mathematical functions and input parameters used in the PEARL agent-based computer simulation model are available at www.PEARLHIVmodel.org.

https://pearlhivmodel.org/method_details.html

