## Supplemental materials for "The projected prevalence of comorbidities and multimorbidity in people with HIV in the United States through the year 2030"

**SUPPLEMENT**

**Table S1:** Definitions of highly prevalent risk factors and comorbidities measured in the NA-ACCORD and included in the PEARL model

| **Risk factor or comorbidity** | **Data source** | **Definition** |
| --- | --- | --- |
| Smoking | EHR and patient-reported surveys (when available) | Ever having EHR-based evidence or patient-reported smoking (collected via surveys) while under observation |
| Obesity | EHR | Body mass index ≥30 kg/m2 (height and weight measurements are exported from EHR) |
| Hepatitis C virus infection | EHR | Ever having EHR-based evidence of HCV infection while under observation, defined as:  1) a positive HCV antibody test, OR  2) a detectable HCV RNA, OR  3) the presence of an HCV genotype test |
| Depression^1^ | EHR | An absorbing state from the month/day/year an individual has a major depressive disorder diagnosis. |
| Anxiety^1^ | EHR | An absorbing state from the month/day/year an individual has a generalized anxiety disorder diagnosis |
| Dyslipidemia^2^ | EHR | An absorbing state from the month/day/year an individual first meets the following definition:  1) total cholesterol ≥240 mg/dL, OR  2) HDL ≤40 mg/dL for men or ≤50 for women, OR  3) LDL ≥130 mg/dL, OR  4) lipid-lowering medication prescription |
| Hypertension^3^ | EHR | An absorbing state from the month/day/year an individual first meets the following definition:  1) prescription of anti-hypertension medications, AND  2) a hypertension diagnosis |
| Stage ≥3 Chronic Kidney Disease^3^ | EHR | An absorbing state from the month/day/year an individual first meets the following definition: ever had eGFR <60 mL/min/1.73m2 consistently for at least 3 months  eGFR estimated using CKD-Epi equation that included an indicator for race. |
| Diabetes^3^ | EHR | An absorbing state from the month/day/year an individual first meets the following definition:  1) HgA1c >6.5%, OR  2) diabetes specific medication was prescribed OR  3) a Type 2 or unspecified diabetes diagnosis was recorded and diabetes-related medications were prescribed |
| Myocardial infarction^4^ | EHR with validation by cardiologists | An absorbing state from the month/day/year an individual who screens positive for a potential MI event has a validated MI event (regardless of MI type) as determined by an adjudication process.  Once an MI event is validated, no subsequent events are investigated. |
| Cancer (all types)^5^ | Cancer registries or EHR data that is validated for cancer diagnosis | An absorbing state from the month/day/year an individual first meets the following definition:  1) Validated cancer diagnosis (all types) confirmed in a cancer registry  2) Validated cancer diagnosis (all types) confirmed with review of the patient’s EHR following screening positive for a potential cancer diagnosis |
| End-stage Renal Disease (ESLD)^6^ | EHR data that is validated for cancer diagnosis | An absorbing state from the month/day/year an individual who screens positive for a potential ESLD event has a validated MI event as determined with review of the patient’s EHR |

Abbreviations:

EHR=electronic health record

**Table S2:** Prevalence and incidence functions applied to PEARL agents who have initiated ART

a1) Anxiety prevalence estimates (from the NA-ACCORD)
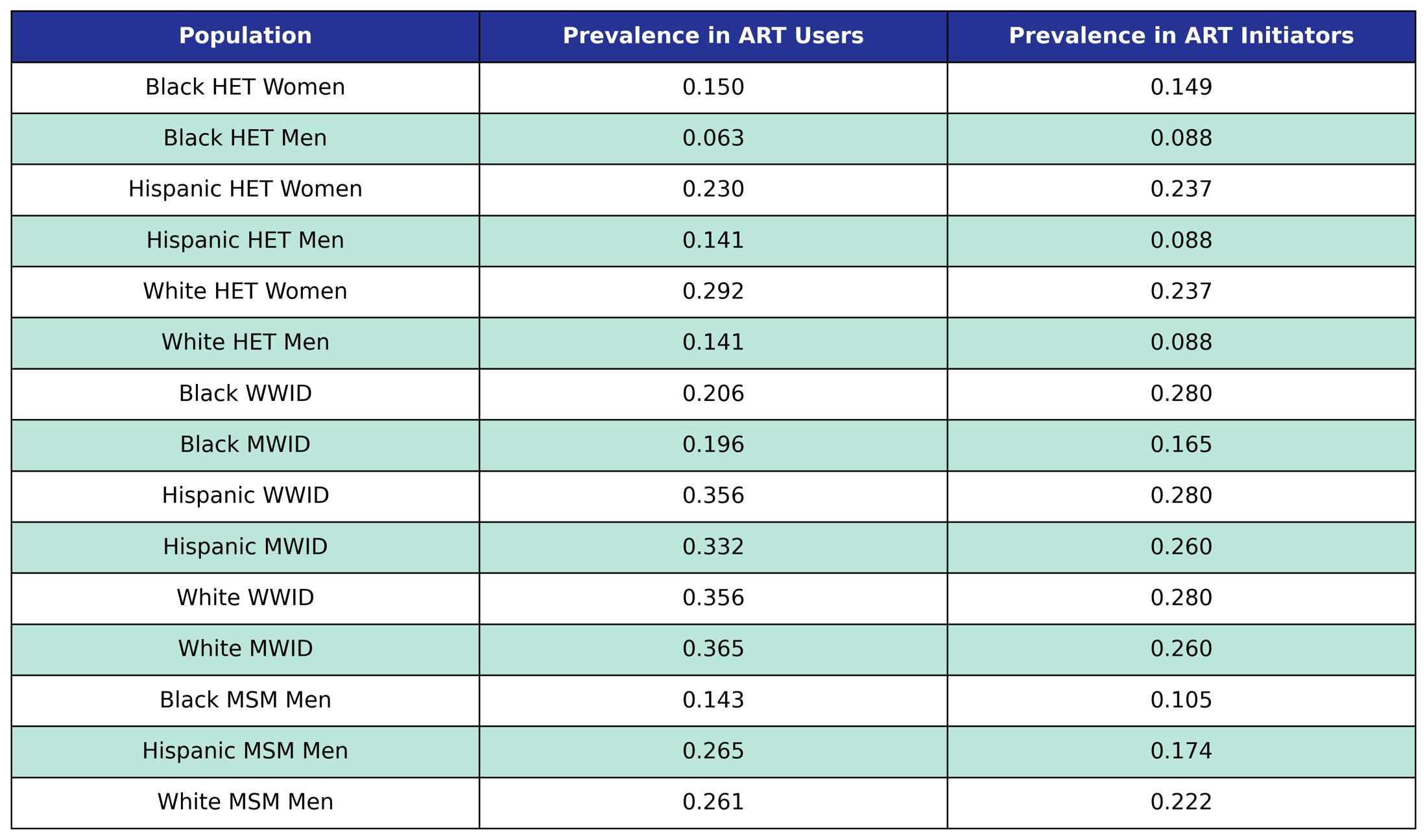

Prevalence in the 2009 ART user population is taken from the 2009 NA-ACCORD population, while prevalence in ART initiators was taken from the 2009 - 2017 NA-ACCORD ART initiator population.

a2) Coefficient estimates from anxiety incidence functions (from the NA-ACCORD)

 
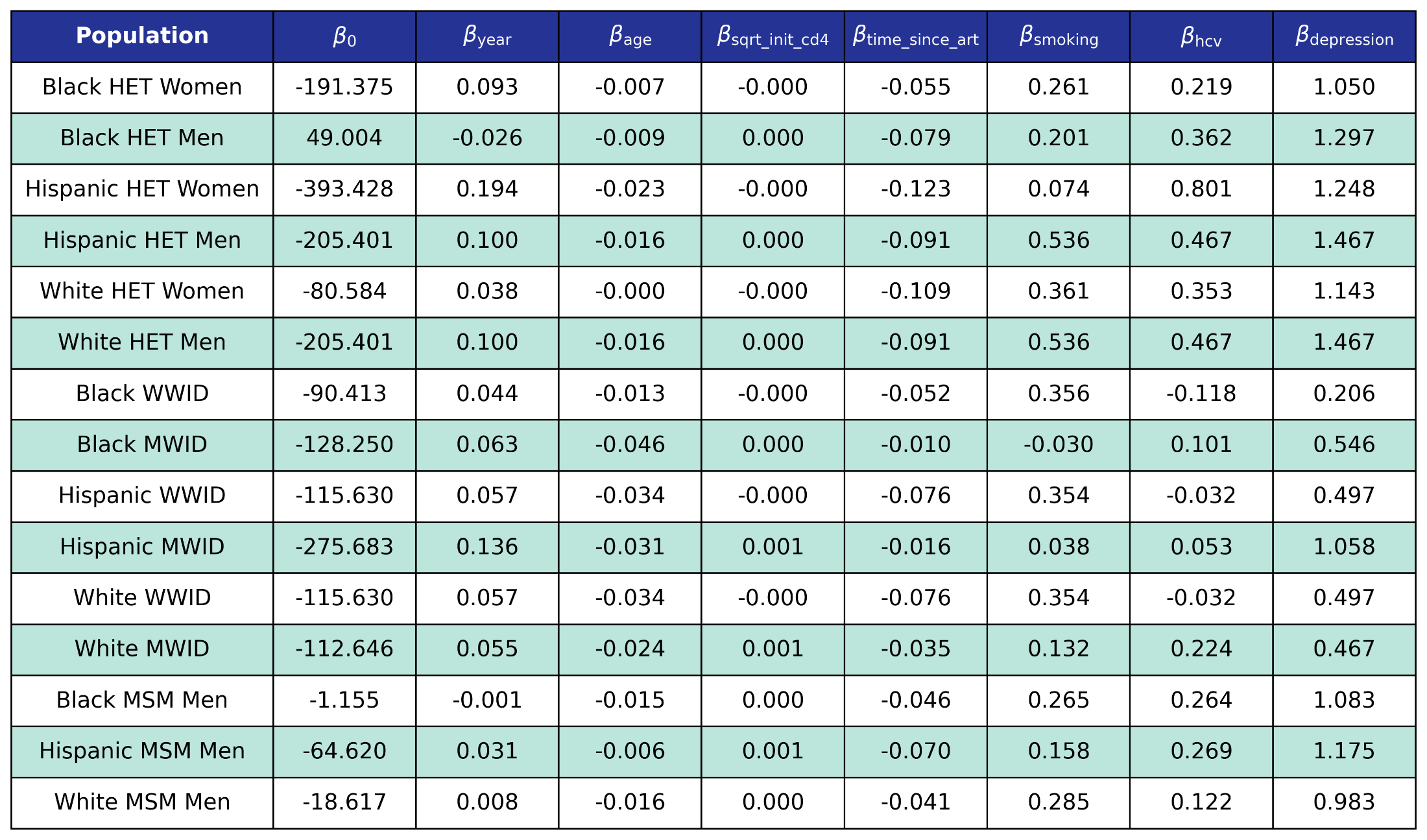

We use logistic regression to model the probability of incidence of anxiety as a function of calendar year (year), age (age), square root of CD4 count at ART initiation (sqrt_init_cd4), number of years since ART initiation (time_since_art), smoking status (smoking), HCV status (hcv) and depression status (depression). The status variables are encoded as Boolean variables.

b1) Depression prevalence estimates (from the NA-ACCORD)
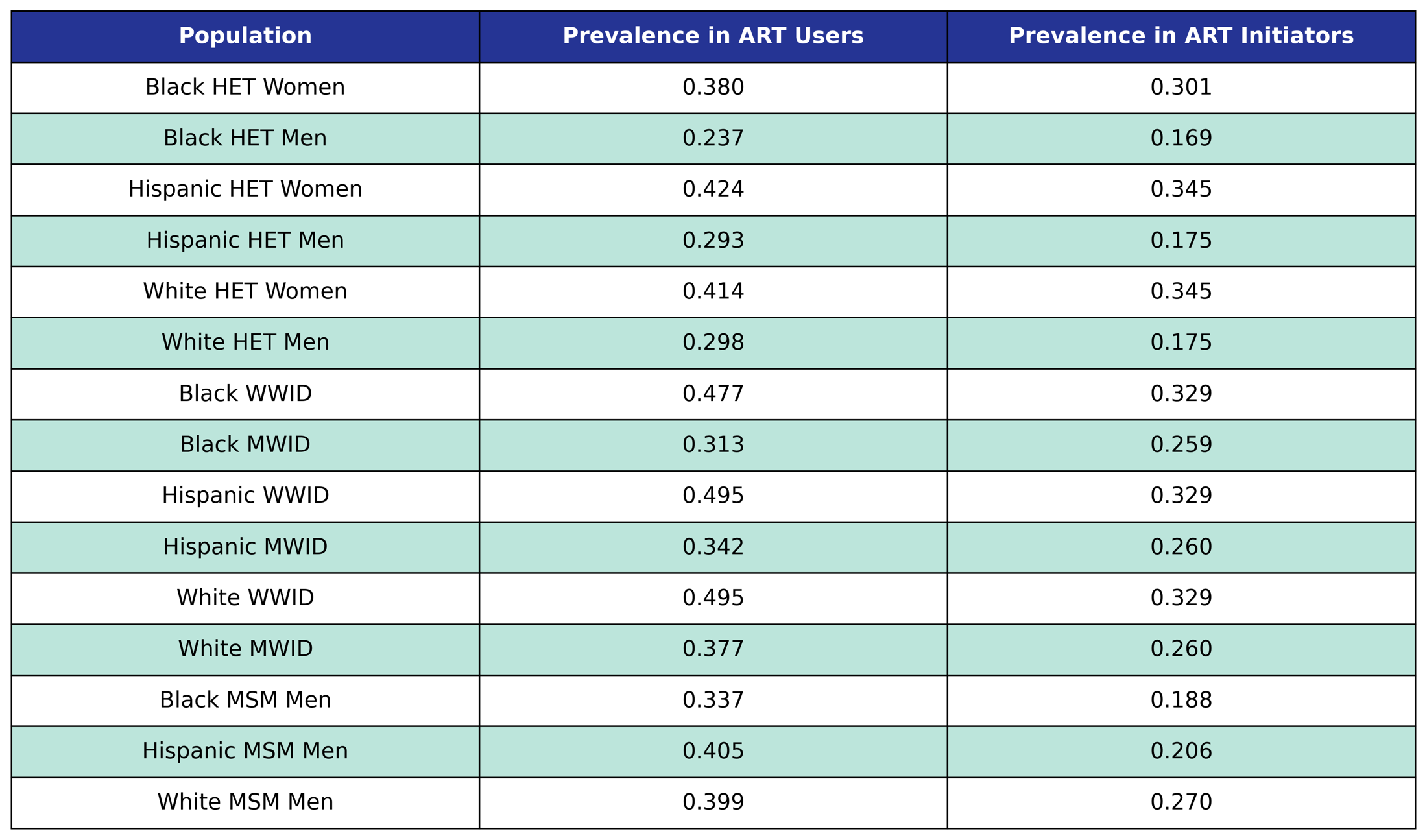

Prevalence in the 2009 ART user population is taken from the 2009 NA-ACCORD population, while prevalence in ART initiators was taken from the 2009 - 2017 NA-ACCORD ART initiator population.

b2) Coefficient estimates from depression incidence functions (from the NA-ACCORD)
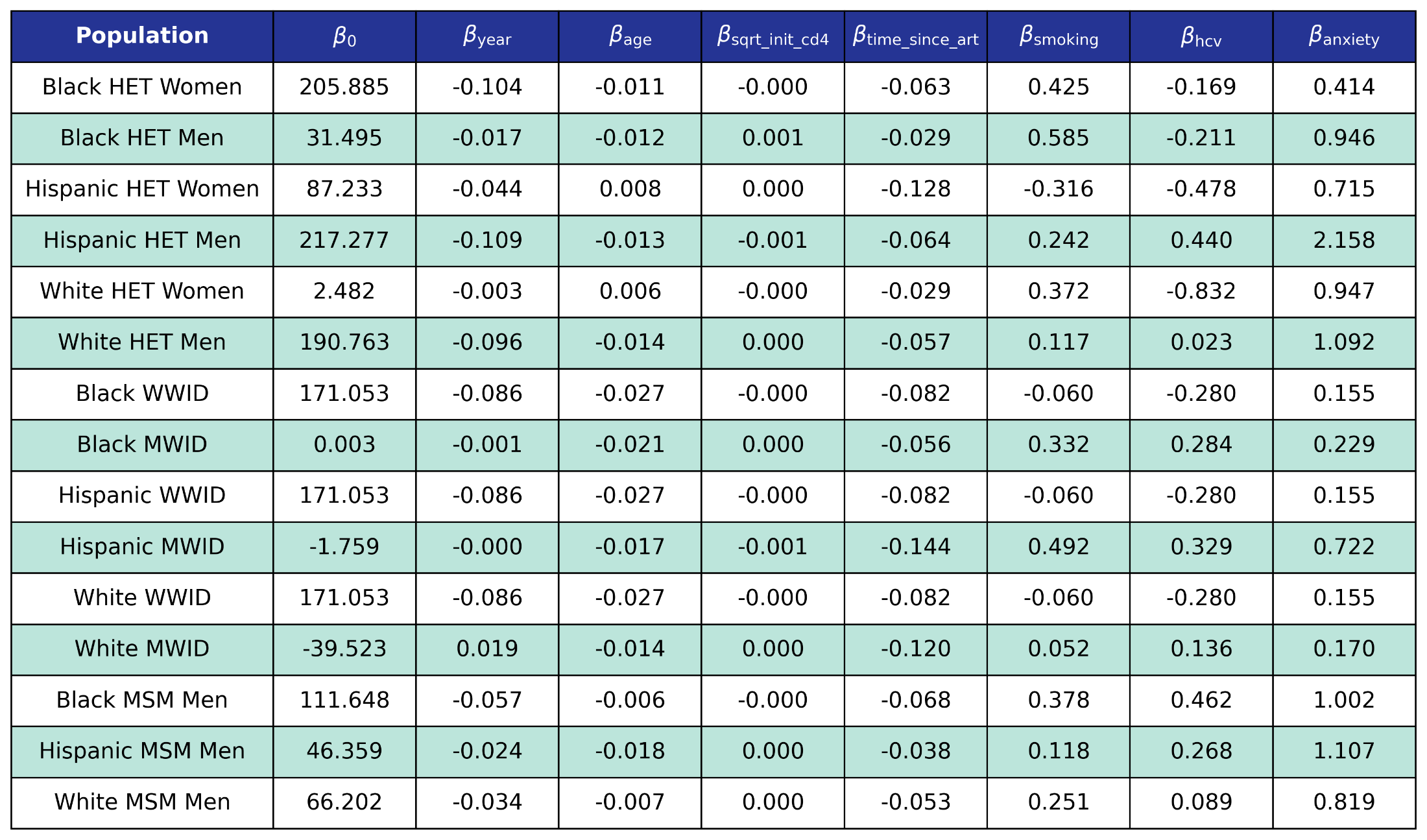

We use logistic regression to model the probability of incidence of depression as a function of calendar year (year), age (age), square root of CD4 count at ART initiation (sqrt_init_cd4), number of years since ART initiation (time_since_art), smoking status (smoking), HCV status (hcv) and anxiety status (anxiety). The status variables are encoded as Boolean variables.

c1) Stage ≥3 chronic kidney disease prevalence estimates (from the NA-ACCORD)
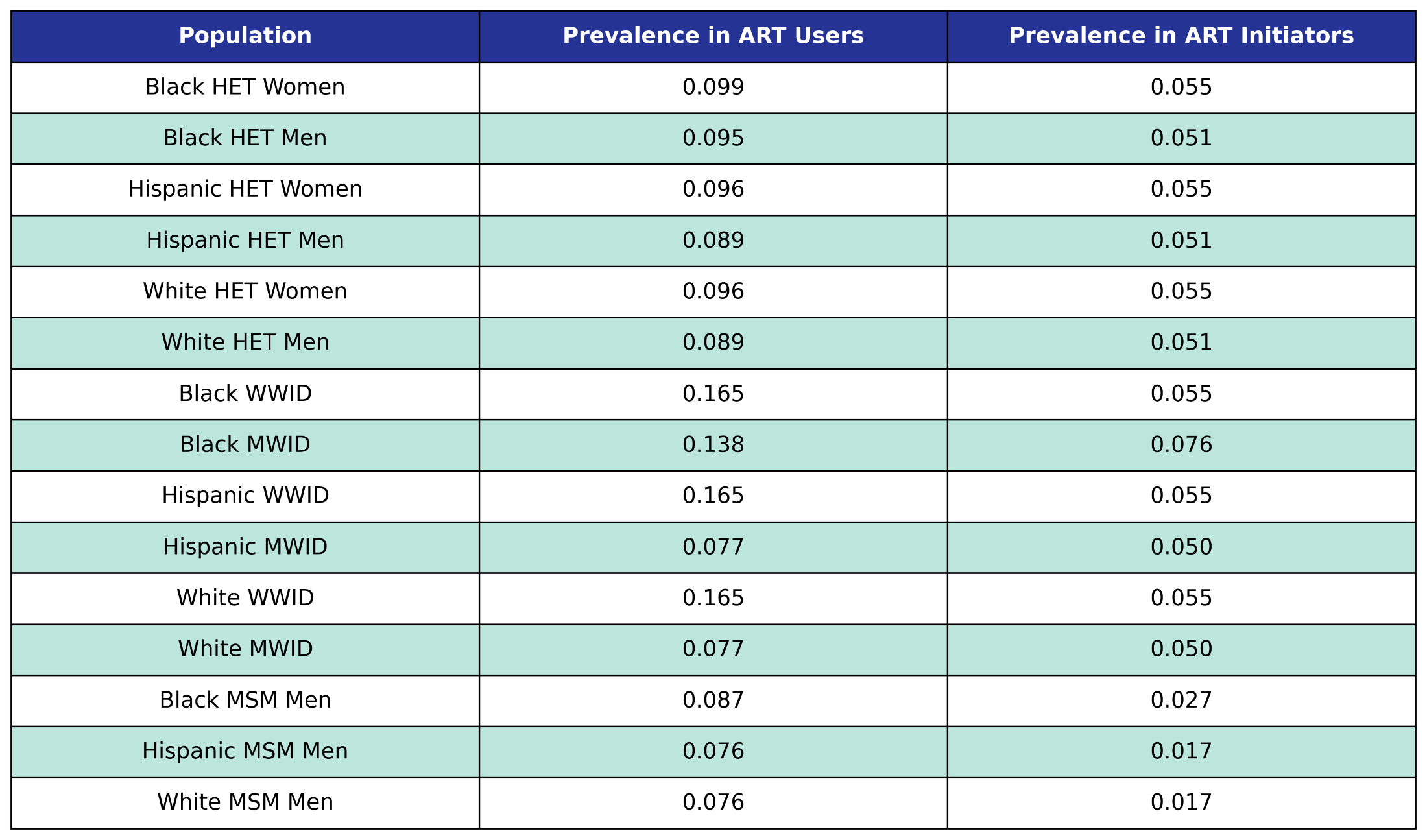

Prevalence in the 2009 ART user population is taken from the 2009 NA-ACCORD population, while prevalence in ART initiators was taken from the 2009 - 2017 NA-ACCORD ART initiator population.

c2) Coefficient estimates from stage ≥3 chronic kidney disease incidence functions (from the NA-ACCORD)
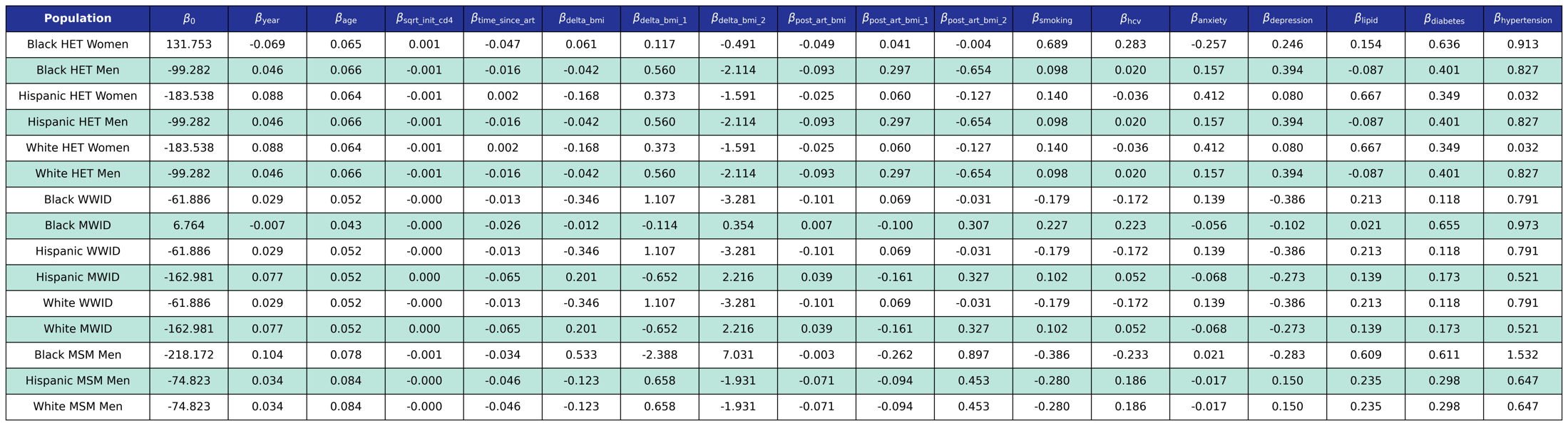

We use logistic regression to model the probability of incidence of this comorbidity as a linear function of calendar year (year), age (age), square root of CD4 count at ART initiation (sqrt_init_cd4), number of years since ART initiation (time_since_art), change in BMI after ART initiation (delta_bmi) and BMI after ART initiation (post_art_bmi) modeled as [restricted cubic splines](https://pearlhivmodel.org/method_details.html#restricted-cubic-spline) (see <https://pearlhivmodel.org/method_details.htm>l for knots), smoking status (smoking), hepatitis C virus (hcv), anxiety (anxiety), depression (depression), dyslipidemia (lipid), diabetes (diabetes), and hypertension (hypertension).

d1) Dyslipidemia prevalence estimates (from the NA-ACCORD)
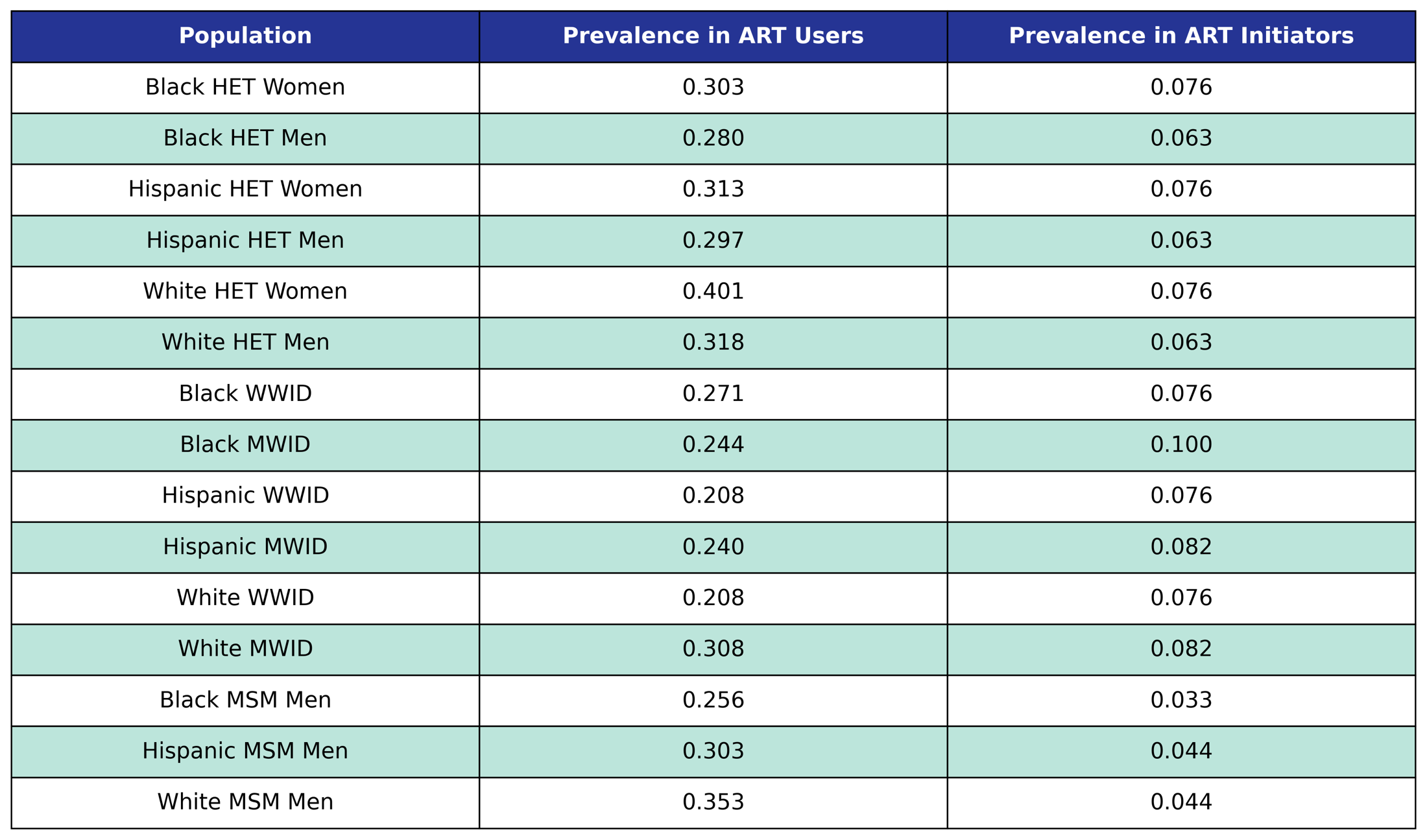

Prevalence in the 2009 ART user population is taken from the 2009 NA-ACCORD population, while prevalence in ART initiators was taken from the 2009 - 2017 NA-ACCORD ART initiator population.

d2) Coefficient estimates from dyslipidemia incidence functions (from the NA-ACCORD)
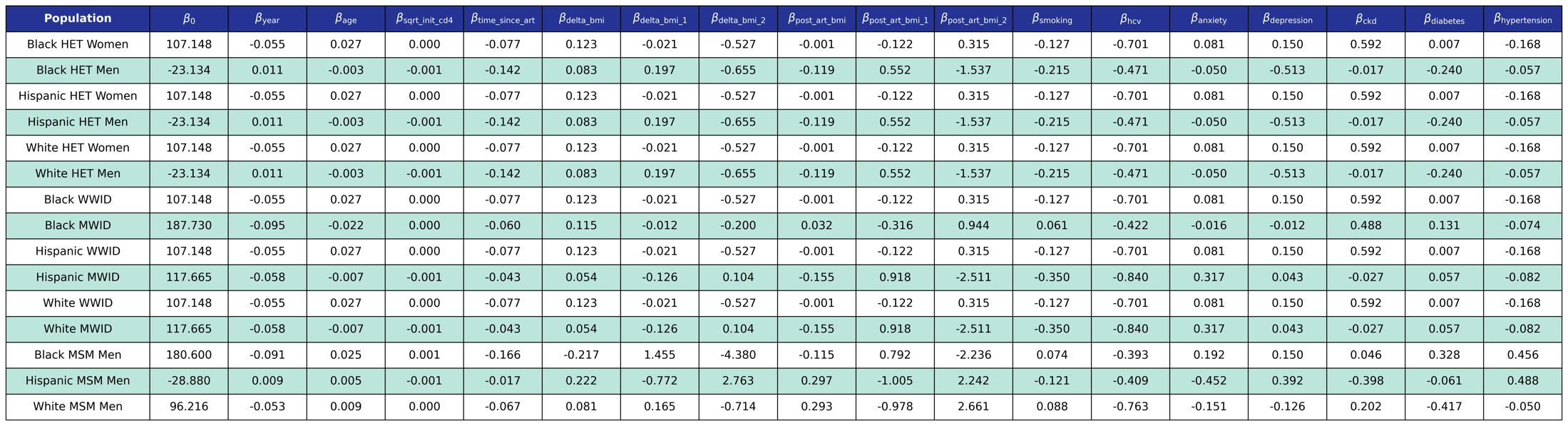

We use logistic regression to model the probability of incidence of this comorbidity as a linear function of calendar year (year), age (age), square root of CD4 count at ART initiation (sqrt_init_cd4), number of years since ART initiation (time_since_art), change in BMI after ART initiation (delta_bmi) and BMI after ART initiation (post_art_bmi) modeled as [restricted cubic splines](https://pearlhivmodel.org/method_details.html#restricted-cubic-spline) (see [https://pearlhivmodel.org/method_details.html](https://pearlhivmodel.org/method_details.html#depression) for knots), smoking status (smoking), hepatitis C virus (hcv), anxiety (anxiety), depression (depression), stage ≥3 chronic kidney disease (ckd), diabetes (diabetes), and hypertension (hypertension).

e1) Diabetes prevalence estimates (from the NA-ACCORD)

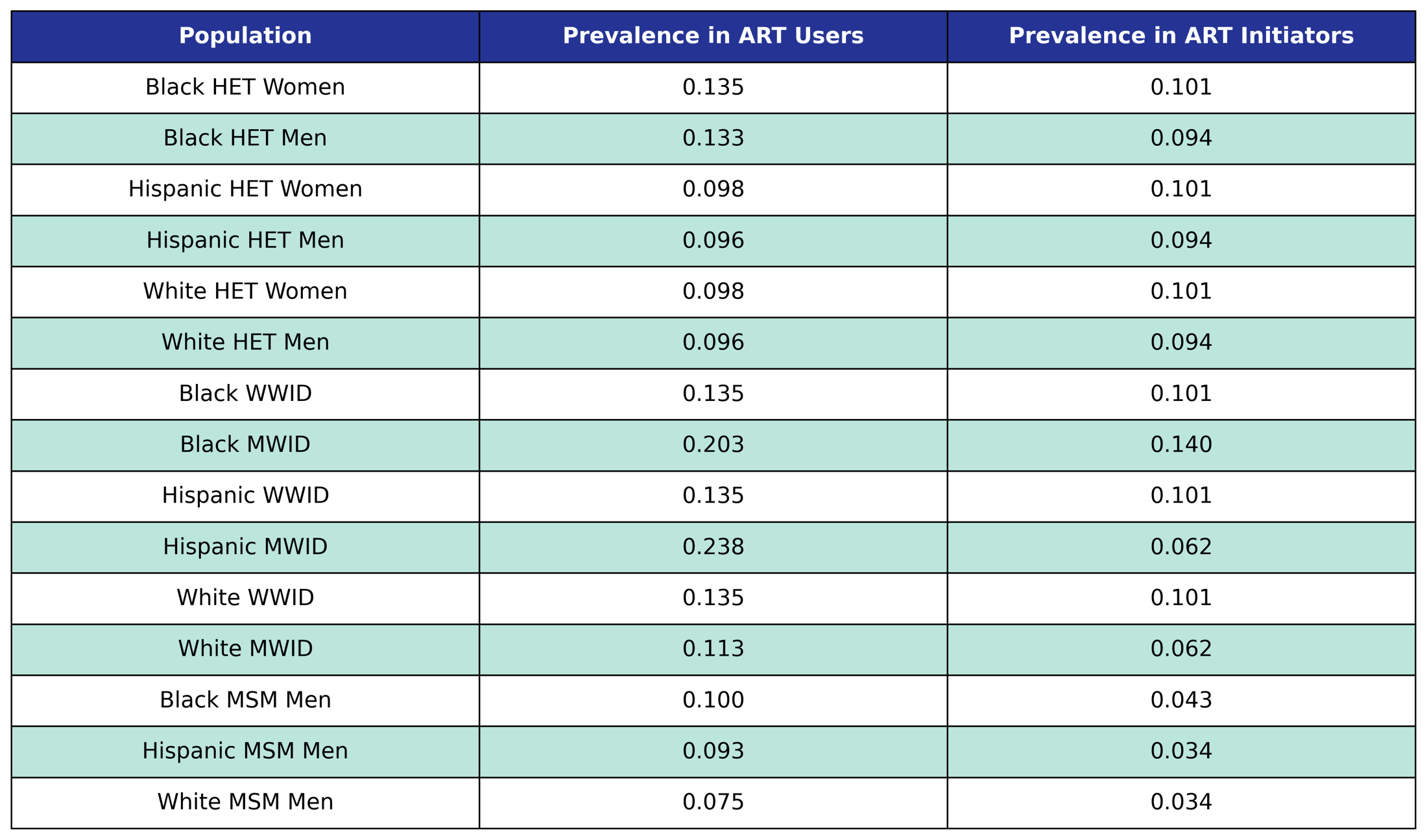

Prevalence in the 2009 ART user population is taken from the 2009 NA-ACCORD population, while prevalence in ART initiators was taken from the 2009 - 2017 NA-ACCORD ART initiator population.

e2) Coefficient estimates from diabetes incidence functions (from the NA-ACCORD)

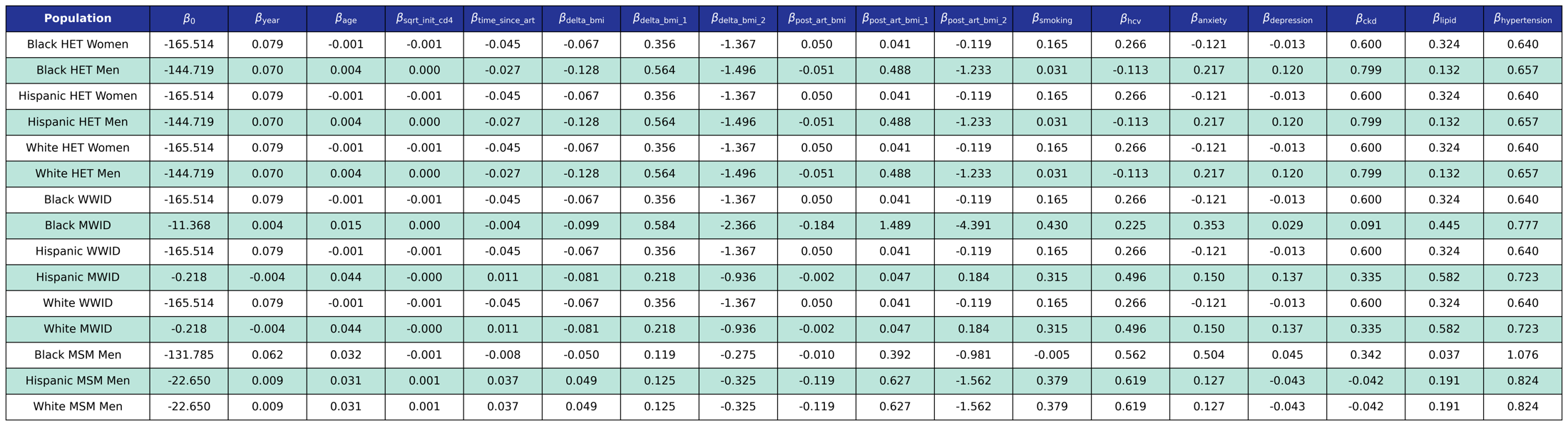

We use logistic regression to model the probability of incidence of this comorbidity as a linear function of calendar year (year), age (age), square root of CD4 count at ART initiation (sqrt_init_cd4), number of years since ART initiation (time_since_art), change in BMI after ART initiation (delta_bmi) and BMI after ART initiation (post_art_bmi) modeled as [restricted cubic splines](https://pearlhivmodel.org/method_details.html#restricted-cubic-spline) (see [https://pearlhivmodel.org/method_details.html](https://pearlhivmodel.org/method_details.html#depression) for knots), smoking status (smoking), hepatitis C virus (hcv), anxiety (anxiety), depression (depression), stage ≥3 chronic kidney disease (ckd), dyslipidemia (lipids), and hypertension (hypertension).

f1) Hypertension prevalence estimates (from the NA-ACCORD)

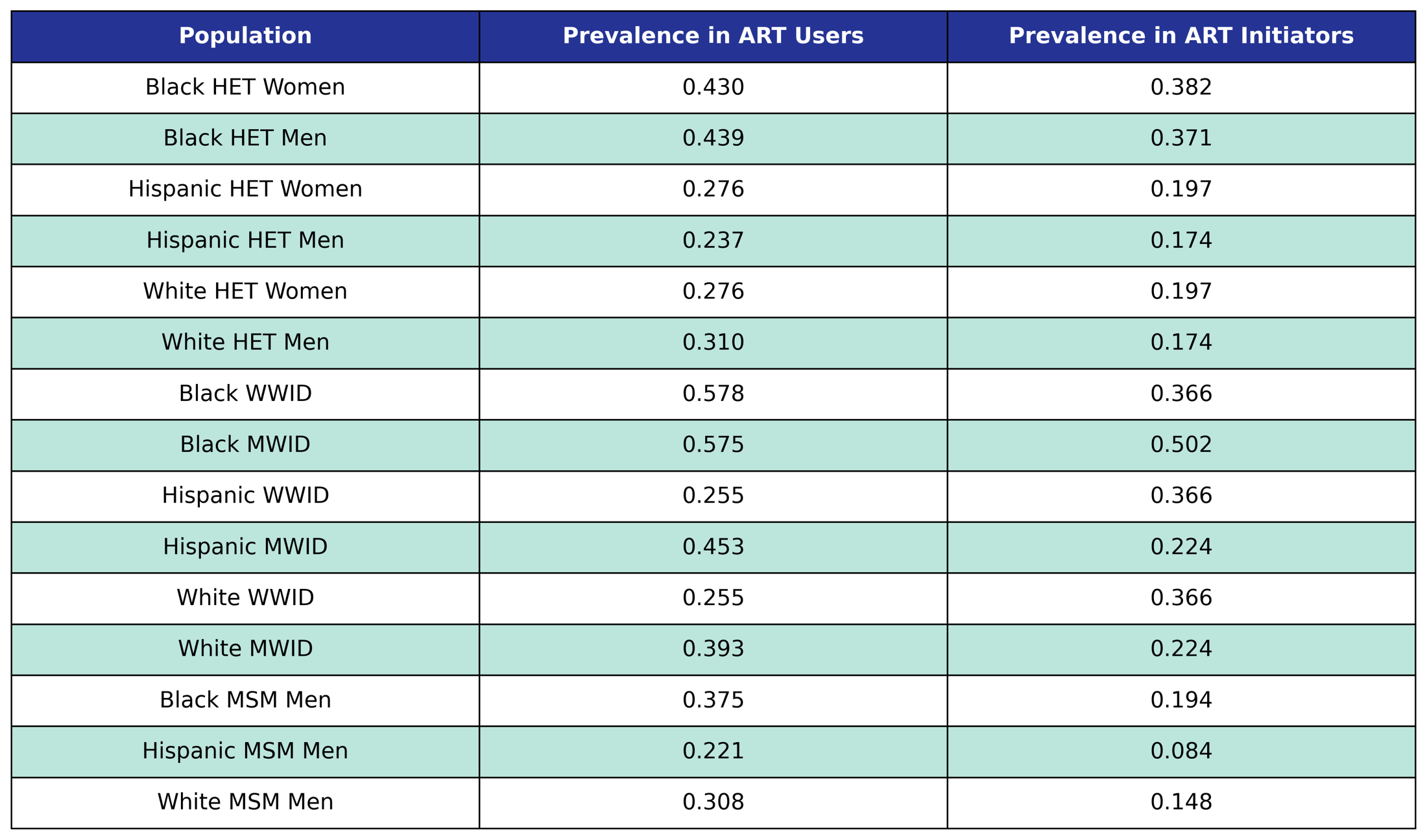

Prevalence in the 2009 ART user population is taken from the 2009 NA-ACCORD population, while prevalence in ART initiators was taken from the 2009 - 2017 NA-ACCORD ART initiator population.

f2) Coefficient estimates from hypertension incidence functions (from the NA-ACCORD)

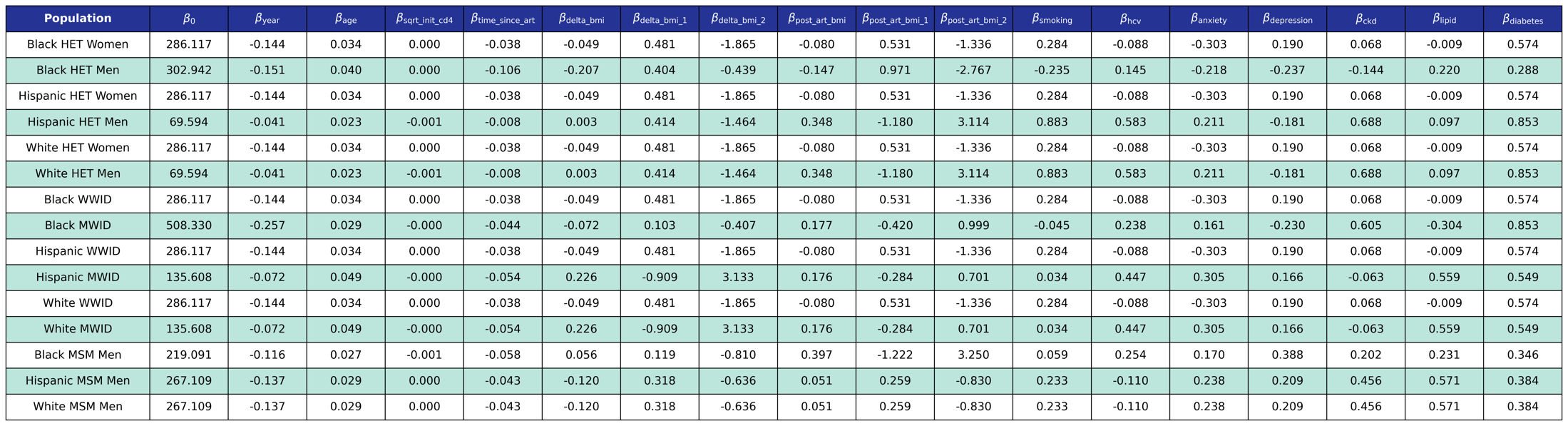

We use logistic regression to model the probability of incidence of this comorbidity as a linear function of calendar year (year), age (age), square root of CD4 count at ART initiation (sqrt_init_cd4), number of years since ART initiation (time_since_art), change in BMI after ART initiation (delta_bmi) and BMI after ART initiation (post_art_bmi) modeled as [restricted cubic splines](https://pearlhivmodel.org/method_details.html#restricted-cubic-spline) (see [https://pearlhivmodel.org/method_details.html](https://pearlhivmodel.org/method_details.html#depression) for knots), smoking status (smoking), hepatitis C virus (hcv), anxiety (anxiety), depression (depression), stage ≥3 chronic kidney disease (ckd), dyslipidemia (lipids), and diabetes (diabetes).

g1) Cancer prevalence estimates (from the NA-ACCORD)

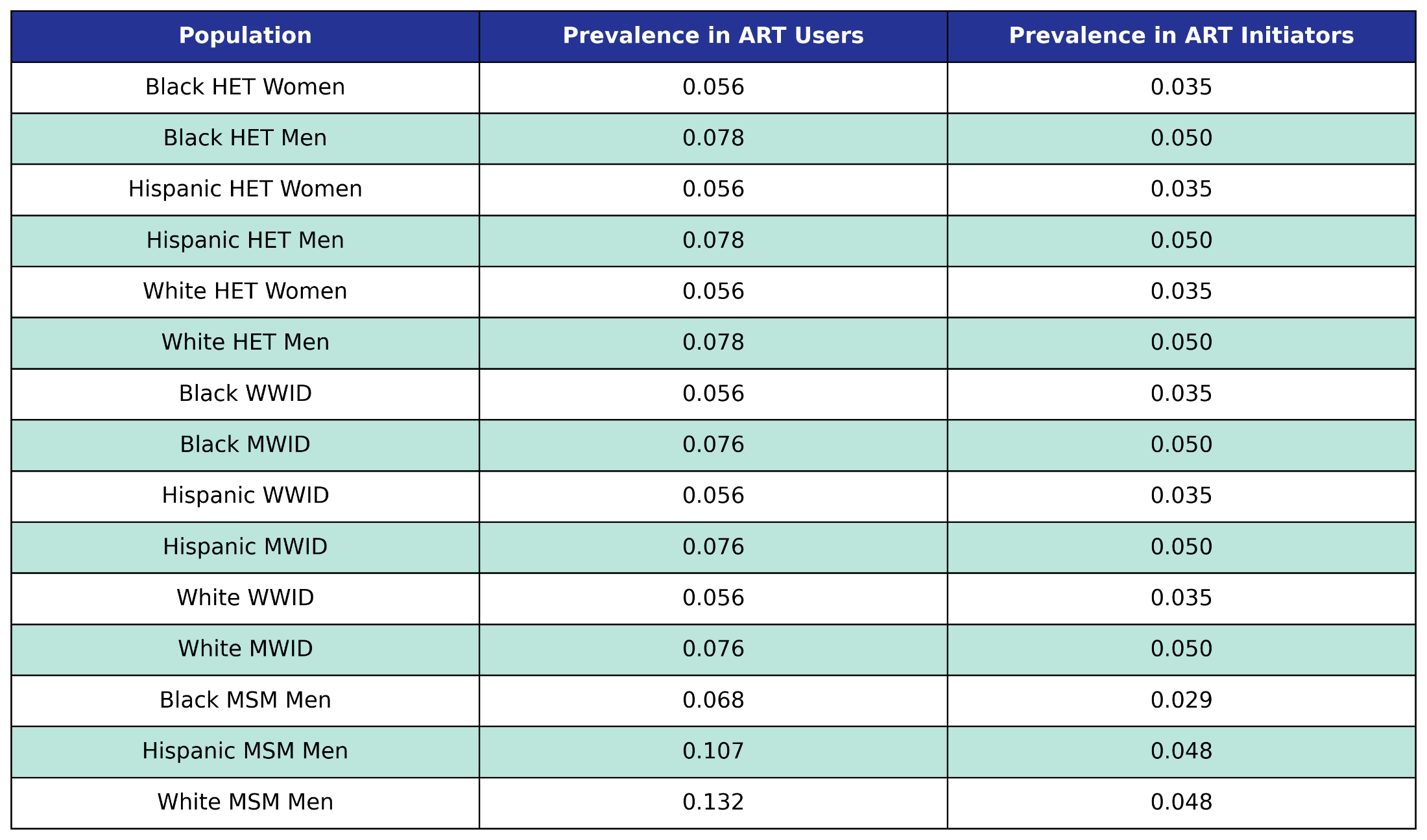

Prevalence in the 2009 ART user population is taken from the 2009 NA-ACCORD population, while prevalence in ART initiators was taken from the 2009 - 2017 NA-ACCORD ART initiator population.

g2) Coefficient estimates from cancer incidence functions (from the NA-ACCORD)

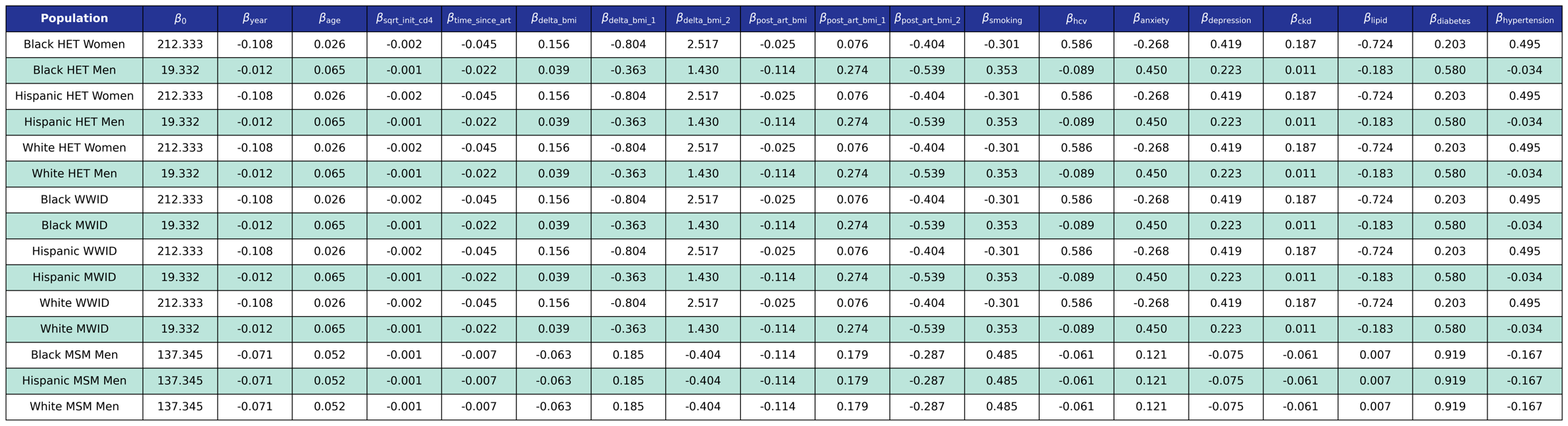

We use logistic regression to model the probability of incidence of this comorbidity as a linear function of calendar year (year), age (age), square root of CD4 count at ART initiation (sqrt_init_cd4), number of years since ART initiation (time_since_art), change in BMI after ART initiation (delta_bmi) and BMI after ART initiation (post_art_bmi) modeled as restricted cubic splines (see [https://pearlhivmodel.org/method_details.html](https://pearlhivmodel.org/method_details.html#depression) for knots), smoking status (smoking), hepatitis C virus (hcv), anxiety (anxiety), depression (depression), stage ≥3 chronic kidney disease (ckd), dyslipidemia (lipid), diabetes (diabetes), and hypertension (hypertension).

h1) End-stage liver disease prevalence estimates (from the NA-ACCORD)

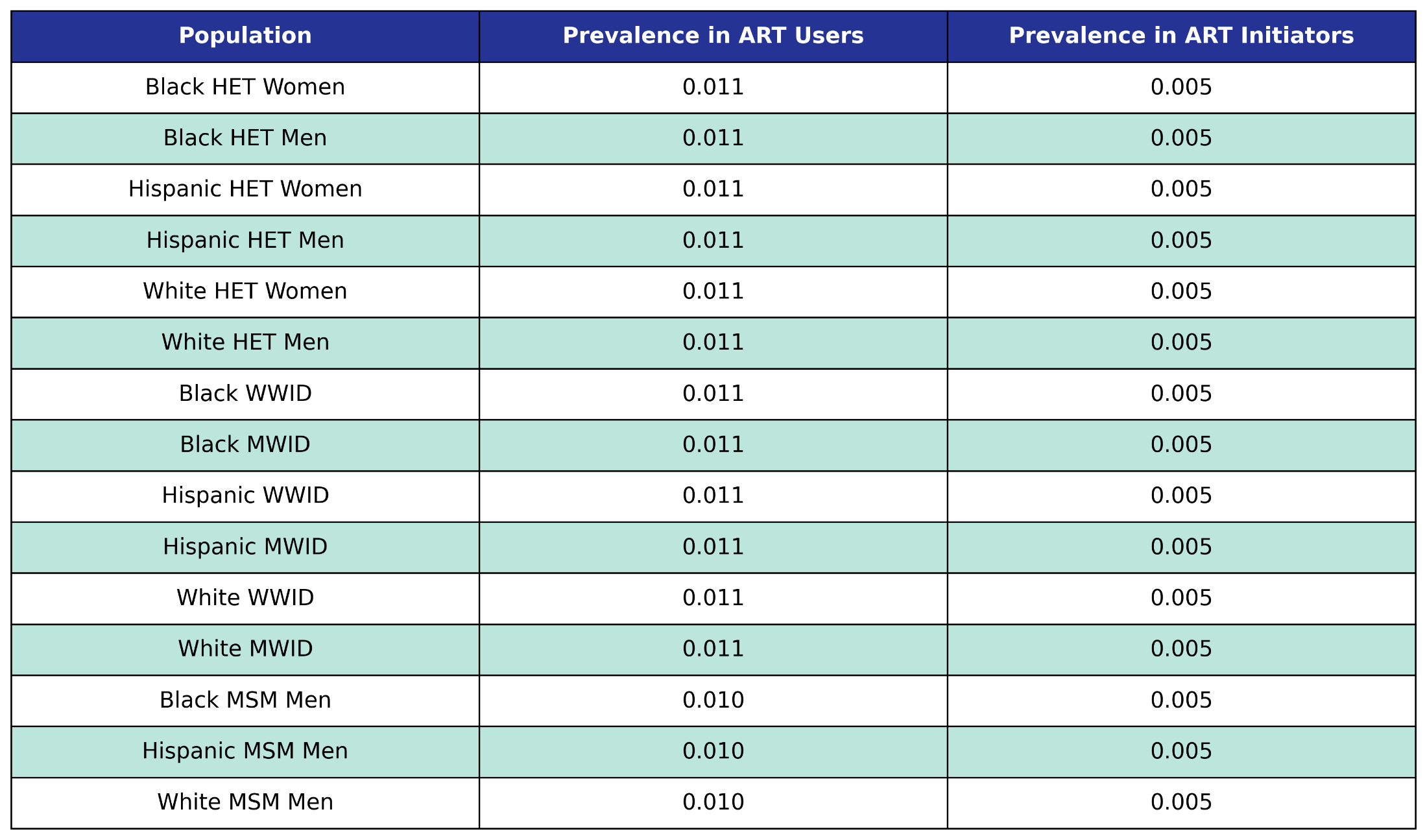

Prevalence in the 2009 ART user population is taken from the 2009 NA-ACCORD population, while prevalence in ART initiators was taken from the 2009 - 2017 NA-ACCORD ART initiator population.

h2) Coefficient estimates from end-stage liver disease incidence functions (from the NA-ACCORD)

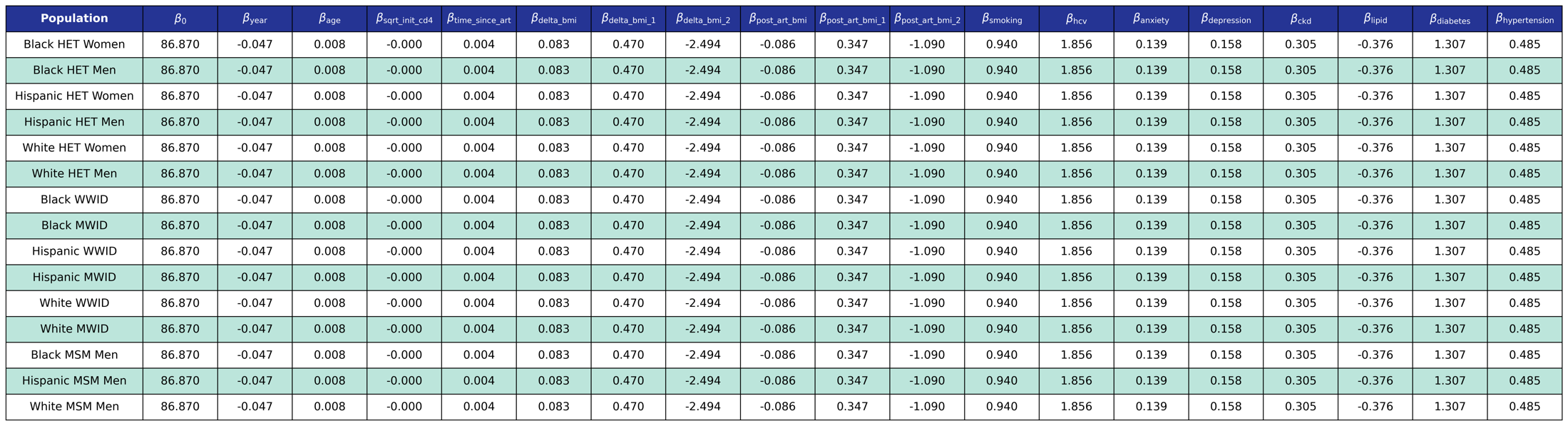

We use logistic regression to model the probability of incidence of this comorbidity as a linear function of calendar year (year), age (age), square root of CD4 count at ART initiation (sqrt_init_cd4), number of years since ART initiation (time_since_art), change in BMI after ART initiation (delta_bmi) and BMI after ART initiation (post_art_bmi) modeled as restricted cubic splines (see [https://pearlhivmodel.org/method_details.html](https://pearlhivmodel.org/method_details.html#depression) for knots), smoking status (smoking), hepatitis C virus (hcv), anxiety (anxiety), depression (depression), stage ≥3 chronic kidney disease (ckd), dyslipidemia (lipid), diabetes (diabetes), and hypertension (hypertension).

i1) Myocardial infarction prevalence estimates (from the NA-ACCORD)

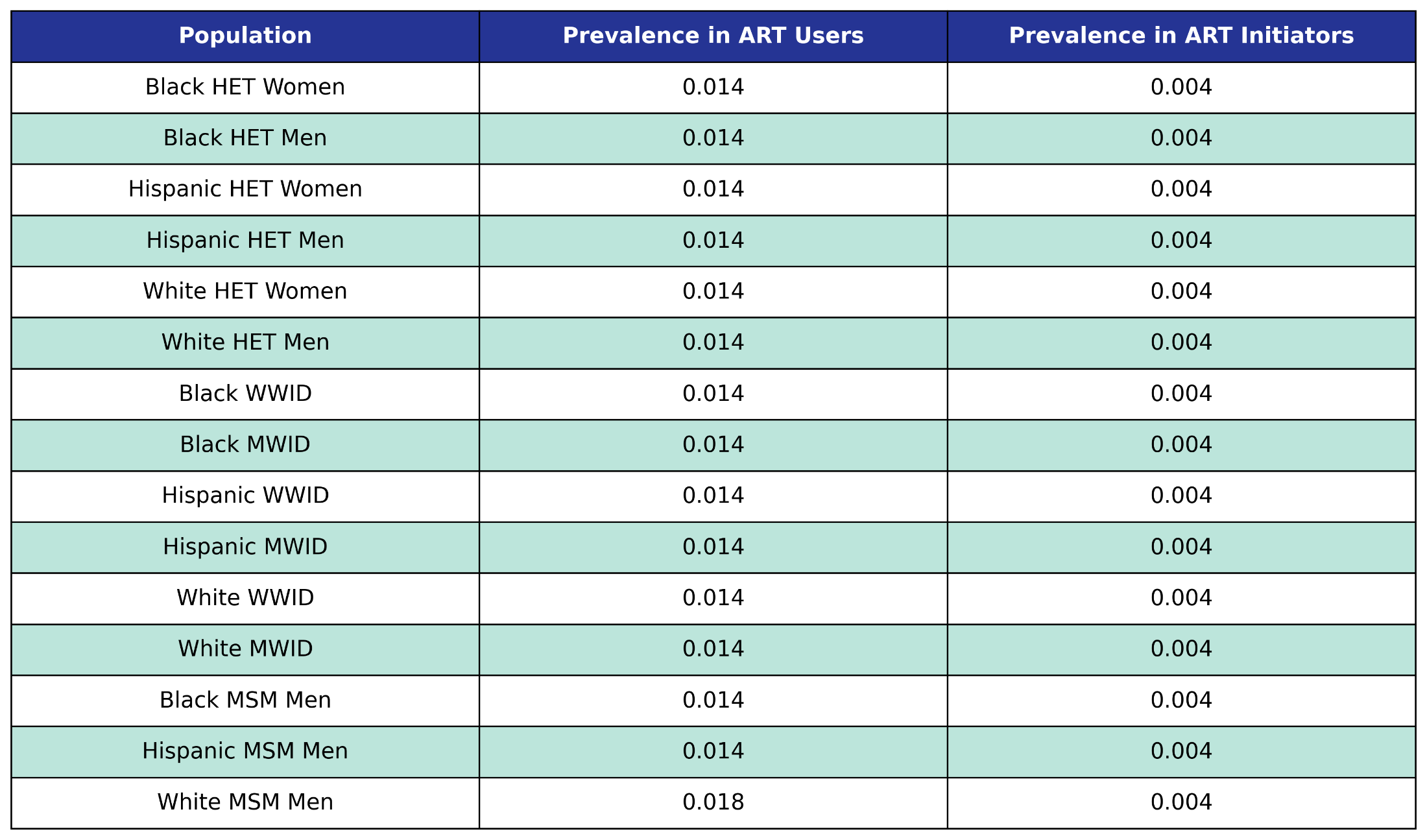
Prevalence in the 2009 ART user population is taken from the 2009 NA-ACCORD population, while prevalence in ART initiators was taken from the 2009 - 2017 NA-ACCORD ART initiator population.

i2) Coefficient estimates from myocardial infarction incidence functions (from the NA-ACCORD)

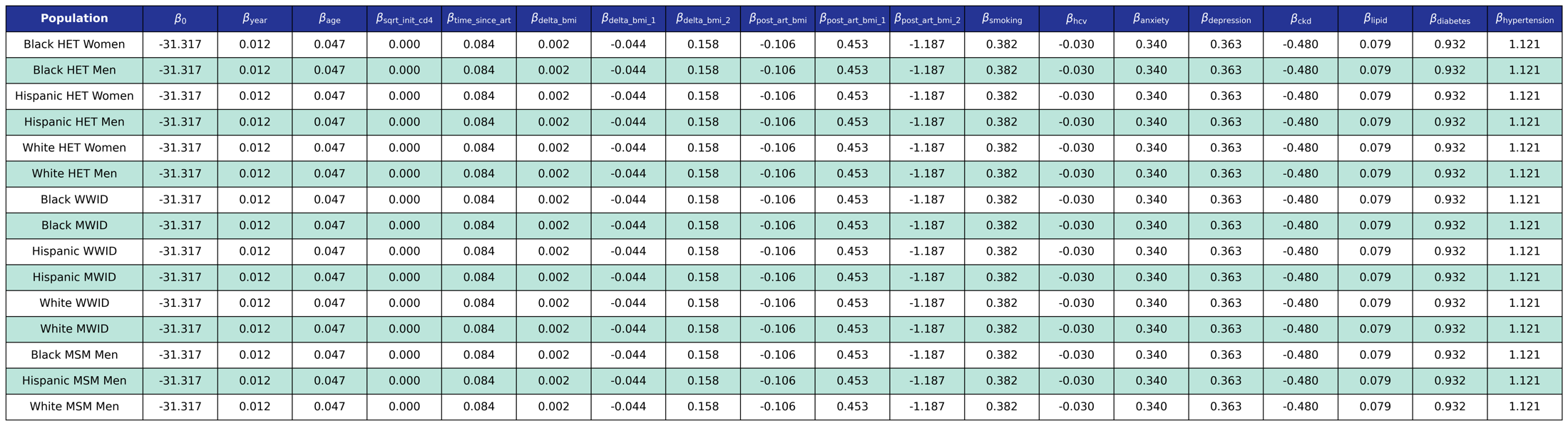

We use logistic regression to model the probability of incidence of this comorbidity as a linear function of calendar year (year), age (age), square root of CD4 count at ART initiation (sqrt_init_cd4), number of years since ART initiation (time_since_art), change in BMI after ART initiation (delta_bmi) and BMI after ART initiation (post_art_bmi) modeled as restricted cubic splines (see [https://pearlhivmodel.org/method_details.html](https://pearlhivmodel.org/method_details.html#depression) for knots), smoking status (smoking), hepatitis C virus (hcv), anxiety (anxiety), depression (depression), stage ≥3 chronic kidney disease (ckd), dyslipidemia (lipid), diabetes (diabetes), and hypertension (hypertension).

**Table S3:** PEARL a) in-care and b) dis-engaged from care mortality functions that include comorbidity presence

a) Coefficient estimates from mortality functions for those in-care

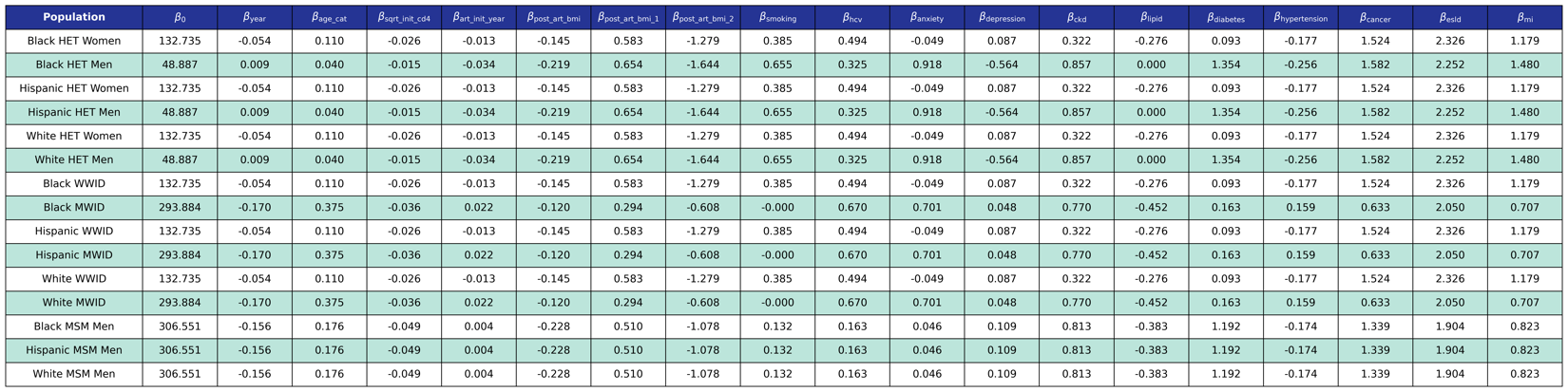

We use logistic regression to model the probability of dying in care as a function of calendar year (year), 10 year age category (age_cat), CD4 count at ART initiation (sqrt_init_cd4), year of ART initiation (art_init_year), BMI after ART initiation (post_art_bmi) modeled as a restricted cubic spline (see [https://pearlhivmodel.org/method_details.html](https://pearlhivmodel.org/method_details.html#depression) for knots), smoking status (smoking), hepatitis C virus (hcv), anxiety (anxiety), depression (depression), stage ≥3 chronic kidney disease (ckd), dyslipidemia (lipids), diabetes (diabetes), hypertension (hypertension), cancer (cancer), end-stage liver disease (esld), and myocardial infarction (mi). The coefficients were estimated using a Generalized Estimating Equation (GEE) with a logit link and an unstructured correlation structure using the geepack software package for R. The NA-ACCORD dataset was restricted to the years 2009-2015 and each patient is represented by a data point for each year they were alive and under observation in NA-ACCORD.

b) Coefficient estimates from mortality functions for those dis-engaged from care and ART

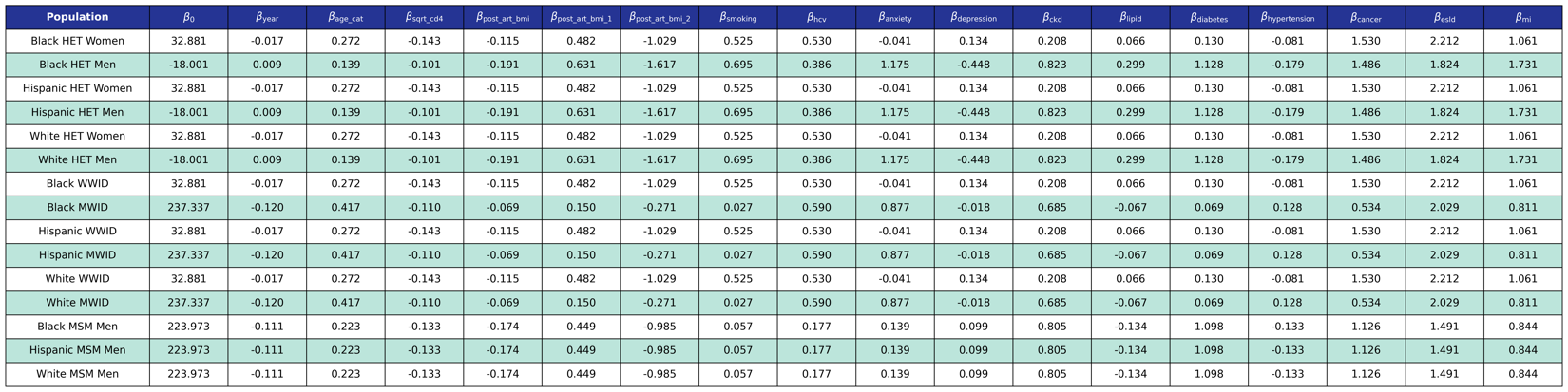

We use logistic regression to model the probability of dying out of care as a function of calendar year (year), 10 year age category (age_cat), CD4 count (sqrt_cd4), BMI after ART initiation (post_art_bmi) modeled as a restricted cubic spline (see [https://pearlhivmodel.org/method_details.html](https://pearlhivmodel.org/method_details.html#depression) for knots), smoking status (smoking), hepatitis C virus (hcv), anxiety (anxiety), depression (depression), stage ≥3 chronic kidney disease (ckd), dyslipidemia (lipids), diabetes (diabetes), hypertension (hypertension), cancer (cancer), end-stage liver disease (esld), and myocardial infarction (mi). The coefficients were estimated using a Generalized Estimating Equation (GEE) with a logit link and an unstructured correlation structure using the geepack software package for R. The NA-ACCORD dataset was restricted to the years 2009-2015.

**Figure S1: Comorbidity incidence validation plots, by subgroup.** Black line=observed annual incidence from the NA-ACCORD; gray shading = the 95% confidence intervals of the observed annual incidence from the NA-ACCORD data. Blue line=projected annual incidence from PEARL. Orange plot shading signals <75% PEARL estimates [total of 7 observed incidence (2009-2015) for MI and ESLD and 9 observed incidence (2009-2017) for all other comorbidities] are within +/-5% of the observed incidence or the 95% confidence interval of the observed incidence [whichever is larger].

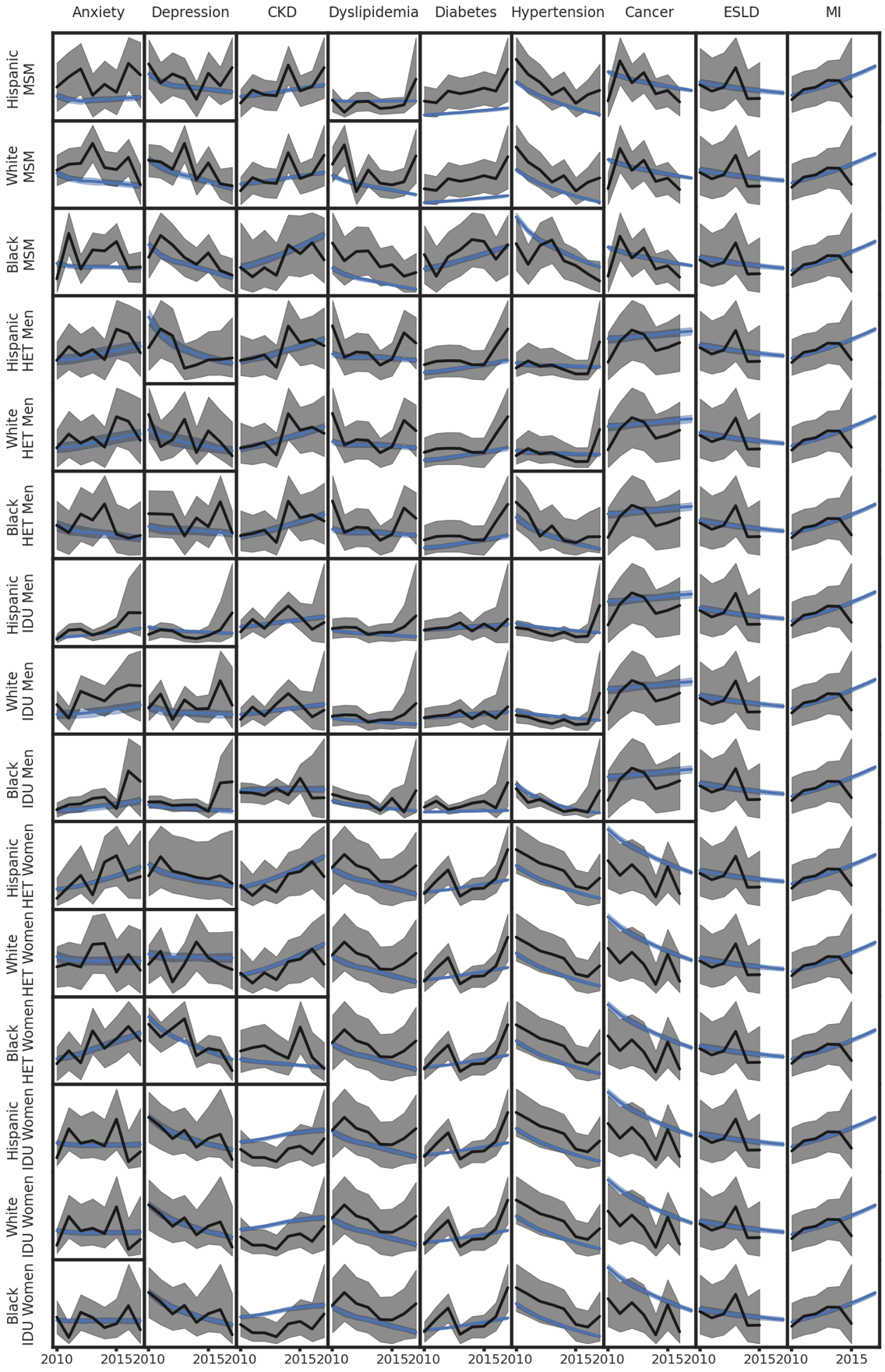

Footnotes:

The y axis is unlabeled and is not the same scale in each plot.

**Figure S2: Comorbidity prevalence validation plots, by subgroup.** Black line=observed annual prevalence from the NA-ACCORD; gray shading = the 95% confidence intervals of the observed annual prevalence from the NA-ACCORD data. Blue line=projected annual prevalence from PEARL. Orange plot = <75% PEARL estimates [total of 7 observed prevalence (2009-2015) for MI and ESLD and 9 observed prevalence (2009-2017) for all other comorbidities] are within +/-5% of the observed prevalence or the 95% confidence interval of the observed prevalence [whichever is larger]. Please note that the orange plots occur among Hispanic sub-groups, white heterosexual men, and Black/AA and White women who injected drugs; these groups have smaller observed sample sizes and were combined with other subgroups resulting in differences between the NA-ACCORD observed estimates and PEARL estimates.

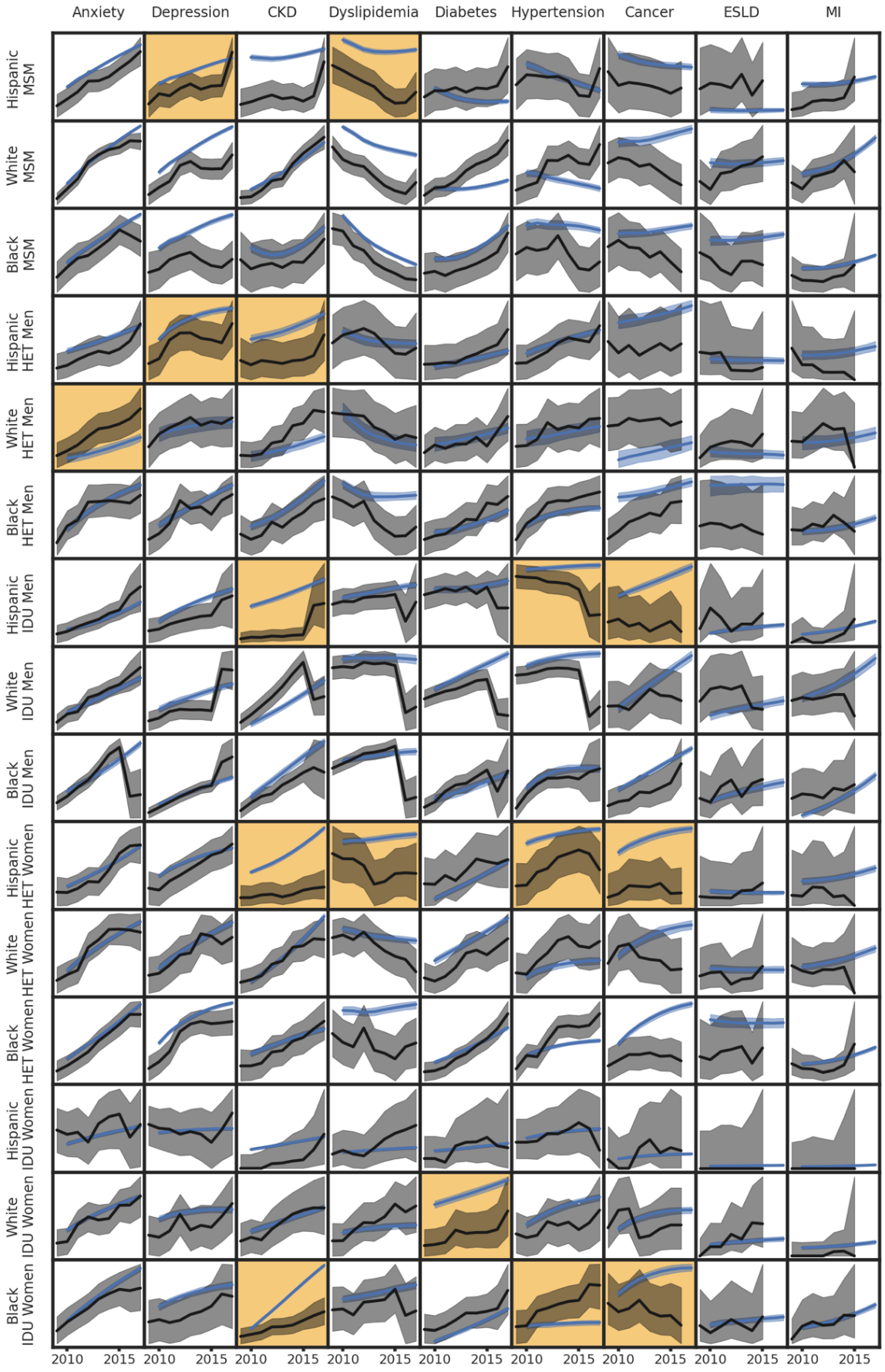

Footnotes:

The y axis is unlabeled and is not the same scale in each plot.

For subgroups with small observed sample size in the NA-ACCORD, the ordered collapsing strategy is: 1) by race/ethnicity, 2) by gender.

**Table S4:** Characteristics of the PEARL-simulated agents using ART, 2020 and 2030

|  | **2020 (Projected)** | | **2030 (Projected)** | |
| --- | --- | --- | --- | --- |
|  | **PEARL^a^** | | **PEARL^a^** | |
|  | **N =** | **674,531 [667,269, 680,808]** | **N =** | **914,738 [879,768, 946,129]** |
| **Characteristics** | **n [95% interquartile range]** | **% [95% interquartile range]** | **n [95% interquartile range]** | **% [95% interquartile range]** |
| **Age (in years)** |  |  |  |  |
| <20 | 699 [623, 777] | 0.1% [0.1%, 0.1%] | 1,210 [644, 1,914] | 0.1% [0.1%, 0.2%] |
| 20-24 | 11,452 [11,045, 11,923] | 1.7% [1.6%, 1.8%] | 12,238 [9,983, 15,393] | 1.3% [1.1%, 1.7%] |
| 25-29 | 40,556 [39,526, 41,461] | 6.0% [5.9%, 6.1%] | 40,794 [35,940, 46,208] | 4.5% [4.0%, 5.0%] |
| 30-34 | 66,908 [65,750, 68,228] | 9.9% [9.8%, 10.0%] | 78,542 [71,076, 86,728] | 8.6% [8.0%, 9.2%] |
| 35-39 | 63,128 [62,239, 63,976] | 9.4% [9.3%, 9.4%] | 101,300 [94,395, 108,110] | 11.1% [10.7%, 11.5%] |
| 40-44 | 61,290 [60,471, 62,008] | 9.1% [9.0%, 9.2%] | 102,383 [97,978, 106,340] | 11.2% [11.0%, 11.4%] |
| 45-49 | 80,046 [79,011, 81,014] | 11.9% [11.8%, 12.0%] | 85,847 [82,747, 89,462] | 9.4% [9.2%, 9.6%] |
| 50-54 | 98,716 [97,661, 99,655] | 14.6% [14.6%, 14.7%] | 81,764 [78,706, 85,619] | 8.9% [8.7%, 9.2%] |
| 55-59 | 101,289 [100,374, 102,209] | 15.0% [14.9%, 15.1%] | 95,226 [92,746, 98,155] | 10.4% [10.2%, 10.7%] |
| 60-64 | 76,974 [76,159, 77,640] | 11.4% [11.3%, 11.5%] | 103,896 [101,773, 106,004] | 11.4% [11.1%, 11.7%] |
| 65-69 | 44,785 [44,299, 45,235] | 6.6% [6.6%, 6.7%] | 95,728 [94,241, 97,315] | 10.5% [10.2%, 10.8%] |
| 70-74 | 19,800 [19,506, 20,051] | 2.9% [2.9%, 3.0%] | 65,606 [64,550, 66,473] | 7.2% [7.0%, 7.4%] |
| ≥75 | 8,969 [8,740, 9,146] | 1.3% [1.3%, 1.4%] | 48,778 [47,990, 49,430] | 5.3% [5.2%, 5.5%] |
| **Male sex** | 518,620 [512,548, 524,273] | 76.9% [76.5%, 77.2%] | 714,764 [682,892, 745,401] | 78.3% [77.1%, 79.4%] |
| **Race** |  |  |  |  |
| White | 218,290 [214,789, 221,569] | 32.4% [32.0%, 32.8%] | 261,230 [244,612, 277,367] | 28.7% [27.1%, 30.4%] |
| Black/AA | 298,157 [294,257, 302,535] | 44.2% [43.7%, 44.7%] | 395,170 [377,556, 417,044] | 43.2% [41.6%, 45.0%] |
| Hispanic | 157,923 [155,092, 161,338] | 23.4% [23.0%, 23.8%] | 256,343 [238,487, 277,014] | 28.1% [26.5%, 29.8%] |
| **Sub-groups** |  |  |  |  |
| MSM | 406,590 [400,695, 411,771] | 60.2% [59.8%, 60.6%] | 587,148 [557,766, 615,889] | 64.2% [62.7%, 65.7%] |
| White MSM | 156,942 [153,957, 160,091] | 23.3% [22.9%, 23.7%] | 180,206 [165,009, 195,275] | 19.7% [18.3%, 21.2%] |
| Black/AA MSM | 144,577 [141,991, 147,215] | 21.4% [21.0%, 21.8%] | 218,374 [205,294, 231,056] | 23.9% [22.3%, 25.3%] |
| Hispanic MSM | 104,670 [102,075, 107,565] | 15.5% [15.2%, 15.9%] | 188,080 [171,296, 206,202] | 20.6% [19.1%, 22.3%] |
| Men who injected drugs (MWID)**^b^** | 59,818 [58,188, 61,571] | 8.9% [8.6%, 9.1%] | 65,672 [58,566, 72,660] | 7.2% [6.4%, 8.0%] |
| White MWID | 23,652 [22,821, 24,596] | 3.5% [3.4%, 3.6%] | 29,460 [25,892, 32,940] | 3.2% [2.8%, 3.6%] |
| Black/AA MWID | 21,163 [20,455, 21,960] | 3.1% [3.0%, 3.3%] | 17,748 [14,339, 21,402] | 1.9% [1.6%, 2.3%] |
| Hispanic MWID | 15,000 [14,319, 15,856] | 2.2% [2.1%, 2.3%] | 18,506 [15,363, 22,119] | 2.0% [1.7%, 2.4%] |
| Women who injected drugs (WWID) | 27,804 [27,214, 28,423] | 4.1% [4.0%, 4.2%] | 31,149 [28,571, 33,665] | 3.4% [3.1%, 3.8%] |
| White WWID | 9,592 [9,385, 9,815] | 1.4% [1.4%, 1.5%] | 13,401 [12,641, 14,275] | 1.5% [1.4%, 1.6%] |
| Black/AA WWID | 14,222 [13,789, 14,647] | 2.1% [2.0%, 2.2%] | 12,936 [11,170, 14,740] | 1.4% [1.2%, 1.6%] |
| Hispanic WWID | 3,994 [3,744, 4,258] | 0.6% [0.6%, 0.6%] | 4,820 [3,716, 5,969] | 0.5% [0.4%, 0.7%] |
| Heterosexual men | 52,364 [50,896, 53,932] | 7.8% [7.5%, 8.0%] | 62,334 [55,279, 69,611] | 6.8% [6.0%, 7.6%] |
| White heterosexual men | 6,840 [6,552, 7,200] | 1.0% [1.0%, 1.1%] | 9,369 [8,129, 10,942] | 1.0% [0.9%, 1.2%] |
| Black/AA heterosexual men | 34,418 [33,156, 35,637] | 5.1% [4.9%, 5.3%] | 39,848 [33,775, 45,447] | 4.4% [3.7%, 5.0%] |
| Hispanic heterosexual men | 11,108 [10,645, 11,564] | 1.6% [1.6%, 1.7%] | 12,998 [10,805, 15,224] | 1.4% [1.2%, 1.7%] |
| Heterosexual women | 128,017 [125,848, 130,489] | 19.0% [18.7%, 19.3%] | 167,442 [158,018, 178,617] | 18.3% [17.2%, 19.5%] |
| White heterosexual women | 21,160 [20,568, 21,912] | 3.1% [3.0%, 3.2%] | 28,882 [26,130, 32,486] | 3.2% [2.8%, 3.6%] |
| Black/AA heterosexual women | 83,713 [82,236, 85,718] | 12.4% [12.2%, 12.7%] | 106,270 [99,595, 115,531] | 11.7% [10.8%, 12.6%] |
| Hispanic heterosexual women | 23,094 [22,053, 23,926] | 3.4% [3.3%, 3.6%] | 31,992 [27,464, 35,894] | 3.5% [3.0%, 4.0%] |
| **Mental Comorbidities** |  |  |  |  |
| Anxiety | 246,192 [243,773, 248,355] | 36.5% [36.3%, 36.7%] | 428,545 [415,882, 441,413] | 46.9% [46.4%, 47.6%] |
| Depression | 317,676 [314,736, 320,336] | 47.1% [47.0%, 47.3%] | 445,518 [433,042, 457,839] | 48.8% [48.3%, 49.3%] |
| ≥1 mental comorbidity | 405,332 [401,727, 408,753] | 60.1% [59.9%, 60.3%] | 589,178 [570,913, 607,285] | 64.5% [64.0%, 65.1%] |
| **Physical Comorbidities** |  |  |  |  |
| Chronic Kidney Disease | 111,940 [110,943, 112,766] | 16.6% [16.5%, 16.7%] | 227,423 [222,269, 231,579] | 24.9% [24.3%, 25.5%] |
| Dyslipidemia | 184,860 [183,476, 186,227] | 27.4% [27.2%, 27.5%] | 234,910 [230,381, 239,349] | 25.7% [25.2%, 26.3%] |
| Diabetes | 105,239 [104,253, 106,039] | 15.6% [15.5%, 15.7%] | 219,064 [213,758, 224,026] | 24.0% [23.5%, 24.6%] |
| Hypertension | 239,688 [237,624, 241,812] | 35.5% [35.4%, 35.7%] | 287,116 [279,586, 295,007] | 31.4% [30.9%, 32.0%] |
| Cancer | 75,944 [75,271, 76,556] | 11.3% [11.2%, 11.3%] | 104,010 [101,267, 106,318] | 11.4% [11.2%, 11.6%] |
| End-Stage Liver Disease | 9,062 [8,903, 9,251] | 1.3% [1.3%, 1.4%] | 13,334 [13,036, 13,726] | 1.5% [1.4%, 1.5%] |
| Myocardial Infarction | 21,934 [21,629, 22,205] | 3.2% [3.2%, 3.3%] | 72,294 [71,454, 72,971] | 7.9% [7.7%, 8.2%] |
| **Physical Multimorbidity** |  |  |  |  |
| No physical comorbidities | 237,779 [234,498, 241,111] | 35.3% [35.0%, 35.4%] | 314,013 [294,246, 332,028] | 34.3% [33.3%, 35.2%] |
| 1 physical comorbidity | 226,742 [224,319, 229,149] | 33.6% [33.5%, 33.7%] | 275,250 [263,964, 285,241] | 30.1% [29.9%, 30.3%] |
| ≥2 physical comorbidities | 209,865 [208,443, 211,142] | 31.1% [31.0%, 31.3%] | 325,266 [319,409, 330,676] | 35.5% [34.7%, 36.5%] |
| **Mental and physical multimorbidity** |  |  |  |  |
| ≥2 mental or physical comorbidities | 393,150 [389,986, 395,791] | 58.3% [58.1%, 58.5%] | 579,564 [564,560, 593,592] | 63.4% [62.5%, 64.5%] |
| ≥1 mental and ≥2 physical comorbidities | 139,335 [138,445, 140,283] | 20.7% [20.5%, 20.8%] | 232,310 [228,637, 235,724] | 25.4% [24.8%, 26.1%] |
| **ART status** |  |  |  |  |
| PWH using ART | 674,531 [667,269, 680,808] | n/a | 914,738 [879,768, 946,129] | n/a |
| ART initiators | 32,963 [31,720, 34,059] | n/a | 33,324 [28,488, 37,749] | n/a |
| Disengaged from ART use^d^ | 41,884 [41,375, 42,430] | n/a | 33,208 [31,780, 34,612] | n/a |

Footnotes:

AA=African American

ART=antiretroviral therapy

PEARL = ProjEcting Age, multimoRbidity, and poLypharmacy in Adults with HIV

PWH=people with HIV

MSM=men who have sex with men

^a^Values represent the median and uncertainty ranges for each simulated outcome across 200 random simulation replications.

^b^MSM who also have MWID as their HIV acquisition risk factor were include in the MWID HIV acquisition risk group.

^c^Percentages in this table are calculated using the median numerator and median denominator from 200 simulation runs of the PEARL model so that percentages will sum to 100%.

^d^PEARL projections of PWH using ART do not include 41,884 [41,375, 42,430] and 33,208 [31,780, 34,612] people who initiated ART but were not using ART treatments in 2020 and 2030 (respectively) and people of ‘other’ race and ethnicities.

**Table S5:** PEARL-projected multimorbidity prevalence, by year^a^ and within each subgroup of PWH using ART in the US

| Subgroup | **Multimorbidity measurement** | **2009** | **2010** | | **2011** | | **2012** | | **2013** | | **2014** | | **2015** | | **2016** | | **2017** | | **2018** | | **2019** | | **2020** | | **2021** | | **2022** | | **2023** | | **2024** | | **2025** | | **2026** | | **2027** | | **2028** | | **2029** | | **2030** |
| --- | --- | --- | --- | --- | --- | --- | --- | --- | --- | --- | --- | --- | --- | --- | --- | --- | --- | --- | --- | --- | --- | --- | --- | --- | --- | --- | --- | --- | --- | --- | --- | --- | --- | --- | --- | --- | --- | --- | --- | --- | --- | --- | --- |
| Overall | ≥1 Ment. | 51% | 53% | | 54% | | 55% | | 56% | | 56% | | 57% | | 58% | | 58% | | 59% | | 60% | | 60% | | 61% | | 61% | | 62% | | 62% | | 62% | | 63% | | 63% | | 64% | | 64% | | 64% |
| Overall | ≥2 Phys. | 25% | 26% | | 27% | | 27% | | 28% | | 28% | | 29% | | 29% | | 30% | | 30% | | 31% | | 31% | | 32% | | 32% | | 32% | | 33% | | 33% | | 34% | | 34% | | 35% | | 35% | | 36% |
| Overall | ≥2 Any | 50% | 52% | | 53% | | 53% | | 54% | | 55% | | 55% | | 56% | | 57% | | 57% | | 58% | | 58% | | 59% | | 59% | | 60% | | 60% | | 61% | | 61% | | 62% | | 62% | | 63% | | 63% |
| Overall | ≥1 Ment. & 2 Phys. | 13% | 14% | | 15% | | 16% | | 17% | | 17% | | 18% | | 19% | | 19% | | 20% | | 20% | | 21% | | 21% | | 22% | | 22% | | 23% | | 23% | | 23% | | 24% | | 24% | | 25% | | 25% |
| White MSM | ≥1 Ment. | 56% | 57% | | 58% | | 59% | | 60% | | 61% | | 62% | | 63% | | 64% | | 64% | | 65% | | 66% | | 66% | | 67% | | 67% | | 68% | | 68% | | 69% | | 69% | | 70% | | 70% | | 71% |
| White MSM | ≥2 Phys. | 25% | 26% | | 26% | | 26% | | 26% | | 27% | | 27% | | 27% | | 28% | | 28% | | 29% | | 29% | | 29% | | 30% | | 30% | | 31% | | 32% | | 32% | | 33% | | 33% | | 34% | | 35% |
| White MSM | ≥2 Any | 52% | 54% | | 54% | | 55% | | 55% | | 56% | | 57% | | 58% | | 58% | | 59% | | 60% | | 60% | | 61% | | 62% | | 62% | | 63% | | 64% | | 64% | | 65% | | 66% | | 66% | | 67% |
| White MSM | ≥1 Ment. & 2 Phys. | 14% | 15% | | 16% | | 16% | | 17% | | 17% | | 18% | | 18% | | 19% | | 19% | | 20% | | 20% | | 21% | | 21% | | 22% | | 23% | | 23% | | 24% | | 24% | | 25% | | 26% | | 26% |
| Black/AA MSM | ≥1 Ment. | 43% | 44% | | 45% | | 45% | | 46% | | 46% | | 47% | | 47% | | 48% | | 48% | | 48% | | 48% | | 49% | | 49% | | 49% | | 49% | | 49% | | 49% | | 49% | | 49% | | 49% | | 49% |
| Black/AA MSM | ≥2 Phys. | 22% | 23% | | 23% | | 22% | | 22% | | 22% | | 23% | | 23% | | 23% | | 23% | | 24% | | 24% | | 24% | | 25% | | 25% | | 26% | | 26% | | 27% | | 27% | | 28% | | 29% | | 29% |
| Black/AA MSM | ≥2 Any | 42% | 43% | | 43% | | 43% | | 43% | | 43% | | 43% | | 43% | | 44% | | 44% | | 44% | | 45% | | 45% | | 45% | | 45% | | 46% | | 46% | | 46% | | 47% | | 47% | | 48% | | 48% |
| Black/AA MSM | ≥1 Ment. & 2 Phys. | 9% | 11% | | 11% | | 11% | | 12% | | 12% | | 12% | | 13% | | 13% | | 13% | | 14% | | 14% | | 14% | | 15% | | 15% | | 15% | | 16% | | 16% | | 16% | | 17% | | 17% | | 17% |
| Hispanic MSM | ≥1 Ment. | 56% | 58% | 58% | | 58% | | 59% | | 59% | | 60% | | 61% | | 61% | | 62% | | 62% | | 63% | | 63% | | 64% | | 65% | | 65% | | 65% | | 66% | | 66% | | 67% | | 67% | | 67% | |
| Hispanic MSM | ≥2 Phys. | 19% | 20% | 19% | | 19% | | 19% | | 19% | | 19% | | 19% | | 19% | | 19% | | 20% | | 20% | | 20% | | 20% | | 20% | | 20% | | 20% | | 21% | | 21% | | 21% | | 21% | | 21% | |
| Hispanic MSM | ≥2 Any | 46% | 48% | 49% | | 48% | | 49% | | 49% | | 50% | | 50% | | 51% | | 52% | | 52% | | 53% | | 53% | | 54% | | 55% | | 55% | | 56% | | 56% | | 57% | | 57% | | 58% | | 58% | |
| Hispanic MSM | ≥1 Ment. & 2 Phys. | 11% | 12% | 12% | | 12% | | 13% | | 13% | | 13% | | 14% | | 14% | | 14% | | 14% | | 15% | | 15% | | 15% | | 15% | | 15% | | 16% | | 16% | | 16% | | 16% | | 16% | | 17% | |
| White MWID | ≥1 Ment. | 60% | 62% | 63% | | 64% | | 65% | | 66% | | 67% | | 68% | | 69% | | 69% | | 70% | | 71% | | 72% | | 73% | | 74% | | 74% | | 75% | | 76% | | 77% | | 78% | | 78% | | 79% | |
| White MWID | ≥2 Phys. | 25% | 28% | 30% | | 31% | | 32% | | 34% | | 35% | | 36% | | 37% | | 38% | | 38% | | 39% | | 40% | | 40% | | 41% | | 41% | | 41% | | 41% | | 42% | | 42% | | 42% | | 42% | |
| White MWID | ≥2 Any | 56% | 59% | 60% | | 61% | | 62% | | 63% | | 64% | | 65% | | 66% | | 67% | | 68% | | 69% | | 69% | | 70% | | 71% | | 71% | | 72% | | 72% | | 73% | | 74% | | 74% | | 75% | |
| White MWID | ≥1 Ment. & 2 Phys. | 15% | 18% | 19% | | 21% | | 22% | | 23% | | 24% | | 26% | | 27% | | 28% | | 28% | | 29% | | 30% | | 31% | | 31% | | 32% | | 33% | | 33% | | 33% | | 34% | | 34% | | 35% | |
| Black/AA MWID | ≥1 Ment. | 45% | 47% | 48% | | 49% | | 51% | | 52% | | 53% | | 55% | | 56% | | 57% | | 58% | | 60% | | 61% | | 62% | | 63% | | 65% | | 66% | | 67% | | 69% | | 70% | | 71% | | 73% | |
| Black/AA MWID | ≥2 Phys. | 36% | 39% | 41% | | 43% | | 44% | | 46% | | 47% | | 48% | | 49% | | 50% | | 51% | | 52% | | 52% | | 53% | | 53% | | 54% | | 54% | | 54% | | 54% | | 53% | | 53% | | 53% | |
| Black/AA MWID | ≥2 Any | 57% | 60% | 62% | | 64% | | 65% | | 67% | | 68% | | 69% | | 70% | | 71% | | 72% | | 73% | | 74% | | 75% | | 76% | | 76% | | 77% | | 78% | | 78% | | 79% | | 79% | | 80% | |
| Black/AA MWID | ≥1 Ment. & 2 Phys. | 16% | 19% | 20% | | 22% | | 23% | | 24% | | 26% | | 27% | | 28% | | 29% | | 30% | | 31% | | 32% | | 33% | | 34% | | 35% | | 36% | | 36% | | 37% | | 38% | | 38% | | 38% | |
| Hispanic MWID | ≥1 Ment. | 56% | 58% | 59% | | 60% | | 62% | | 63% | | 64% | | 66% | | 67% | | 68% | | 70% | | 71% | | 72% | | 74% | | 76% | | 77% | | 79% | | 80% | | 82% | | 84% | | 85% | | 87% | |
| Hispanic MWID | ≥2 Phys. | 30% | 33% | 34% | | 35% | | 36% | | 38% | | 39% | | 40% | | 41% | | 42% | | 43% | | 44% | | 45% | | 46% | | 47% | | 47% | | 49% | | 50% | | 51% | | 53% | | 55% | | 57% | |
| Hispanic MWID | ≥2 Any | 58% | 60% | 62% | | 63% | | 64% | | 65% | | 67% | | 68% | | 70% | | 71% | | 73% | | 74% | | 75% | | 77% | | 78% | | 79% | | 81% | | 82% | | 84% | | 85% | | 86% | | 88% | |
| Hispanic MWID | ≥1 Ment. & 2 Phys. | 17% | 19% | 21% | | 22% | | 23% | | 25% | | 26% | | 27% | | 29% | | 30% | | 31% | | 32% | | 34% | | 35% | | 36% | | 38% | | 39% | | 41% | | 43% | | 45% | | 47% | | 50% | |
| White WWID | ≥1 Ment. | 67% | 70% | 72% | | 73% | | 74% | | 75% | | 76% | | 77% | | 77% | | 78% | | 78% | | 79% | | 79% | | 79% | | 80% | | 80% | | 81% | | 81% | | 81% | | 82% | | 82% | | 83% | |
| White WWID | ≥2 Phys. | 20% | 23% | 25% | | 27% | | 29% | | 31% | | 32% | | 33% | | 34% | | 35% | | 36% | | 37% | | 38% | | 39% | | 40% | | 41% | | 42% | | 42% | | 43% | | 44% | | 45% | | 46% | |
| White WWID | ≥2 Any | 54% | 59% | 62% | | 65% | | 67% | | 69% | | 70% | | 71% | | 72% | | 73% | | 74% | | 74% | | 75% | | 75% | | 76% | | 76% | | 77% | | 77% | | 78% | | 78% | | 79% | | 79% | |
| White WWID | ≥1 Ment. & 2 Phys. | 13% | 16% | 19% | | 20% | | 22% | | 24% | | 25% | | 26% | | 27% | | 28% | | 29% | | 30% | | 31% | | 32% | | 33% | | 34% | | 34% | | 35% | | 36% | | 37% | | 38% | | 39% | |
| Black/AA WWID | ≥1 Ment. | 58% | 62% | 64% | | 66% | | 67% | | 69% | | 70% | | 71% | | 72% | | 73% | | 74% | | 75% | | 75% | | 76% | | 76% | | 77% | | 78% | | 78% | | 78% | | 79% | | 79% | | 79% | |
| Black/AA WWID | ≥2 Phys. | 35% | 40% | 43% | | 47% | | 50% | | 53% | | 56% | | 58% | | 61% | | 63% | | 65% | | 67% | | 69% | | 71% | | 72% | | 73% | | 74% | | 75% | | 76% | | 77% | | 78% | | 78% | |
| Black/AA WWID | ≥2 Any | 63% | 67% | 70% | | 73% | | 75% | | 77% | | 79% | | 81% | | 83% | | 84% | | 86% | | 87% | | 88% | | 89% | | 90% | | 91% | | 91% | | 92% | | 92% | | 93% | | 93% | | 93% | |
| Black/AA WWID | ≥1 Ment. & 2 Phys. | 20% | 24% | 28% | | 31% | | 34% | | 37% | | 39% | | 42% | | 44% | | 47% | | 49% | | 51% | | 52% | | 54% | | 55% | | 57% | | 58% | | 59% | | 60% | | 61% | | 62% | | 63% | |
| Hispanic WWID | ≥1 Ment. | 67% | 70% | 72% | | 73% | | 74% | | 75% | | 76% | | 77% | | 78% | | 78% | | 79% | | 79% | | 79% | | 80% | | 80% | | 81% | | 81% | | 81% | | 81% | | 82% | | 82% | | 82% | |
| Hispanic WWID | ≥2 Phys. | 20% | 24% | 26% | | 29% | | 31% | | 33% | | 35% | | 37% | | 38% | | 40% | | 41% | | 43% | | 44% | | 45% | | 46% | | 47% | | 48% | | 49% | | 50% | | 50% | | 51% | | 52% | |
| Hispanic WWID | ≥2 Any | 54% | 59% | 63% | | 65% | | 68% | | 70% | | 71% | | 73% | | 74% | | 75% | | 76% | | 77% | | 78% | | 79% | | 79% | | 80% | | 80% | | 80% | | 81% | | 81% | | 81% | | 82% | |
| Hispanic WWID | ≥1 Ment. & 2 Phys. | 13% | 17% | 19% | | 21% | | 23% | | 25% | | 27% | | 29% | | 30% | | 32% | | 33% | | 34% | | 35% | | 36% | | 38% | | 39% | | 39% | | 40% | | 41% | | 42% | | 43% | | 44% | |
| White Heterosexual Men | ≥1 Ment. | 40% | 41% | 41% | | 41% | | 41% | | 41% | | 42% | | 42% | | 42% | | 42% | | 43% | | 43% | | 43% | | 44% | | 44% | | 44% | | 45% | | 45% | | 46% | | 46% | | 47% | | 48% | |
| White Heterosexual Men | ≥2 Phys. | 22% | 23% | 23% | | 23% | | 24% | | 24% | | 25% | | 26% | | 26% | | 27% | | 28% | | 28% | | 29% | | 30% | | 31% | | 32% | | 33% | | 34% | | 35% | | 36% | | 37% | | 39% | |
| White Heterosexual Men | ≥2 Any | 41% | 42% | 42% | | 42% | | 42% | | 43% | | 44% | | 44% | | 45% | | 46% | | 47% | | 48% | | 49% | | 50% | | 51% | | 52% | | 53% | | 54% | | 55% | | 56% | | 57% | | 58% | |
| White Heterosexual Men | ≥1 Ment. & 2 Phys. | 9% | 10% | 10% | | 10% | | 11% | | 11% | | 12% | | 12% | | 12% | | 13% | | 13% | | 14% | | 14% | | 15% | | 15% | | 16% | | 16% | | 17% | | 18% | | 18% | | 19% | | 20% | |
| Black/AA Heterosexual Men | ≥1 Ment. | 29% | 30% | 32% | | 33% | | 34% | | 35% | | 36% | | 37% | | 37% | | 38% | | 39% | | 39% | | 40% | | 40% | | 41% | | 41% | | 41% | | 42% | | 42% | | 42% | | 42% | | 43% | |
| Black/AA Heterosexual Men | ≥2 Phys. | 28% | 29% | 30% | | 30% | | 31% | | 31% | | 32% | | 33% | | 34% | | 35% | | 35% | | 36% | | 37% | | 38% | | 39% | | 40% | | 40% | | 41% | | 42% | | 43% | | 44% | | 45% | |
| Black/AA Heterosexual Men | ≥2 Any | 41% | 43% | 44% | | 44% | | 46% | | 47% | | 48% | | 49% | | 50% | | 51% | | 52% | | 52% | | 53% | | 54% | | 55% | | 55% | | 56% | | 57% | | 58% | | 58% | | 59% | | 60% | |
| Black/AA Heterosexual Men | ≥1 Ment. & 2 Phys. | 8% | 9% | 10% | | 10% | | 11% | | 11% | | 12% | | 13% | | 13% | | 14% | | 15% | | 15% | | 16% | | 16% | | 17% | | 17% | | 18% | | 19% | | 19% | | 20% | | 20% | | 21% | |
| Hispanic Heterosexual Men | ≥1 Ment. | 39% | 42% | 44% | | 45% | | 46% | | 47% | | 48% | | 48% | | 49% | | 49% | | 49% | | 50% | | 50% | | 50% | | 51% | | 51% | | 51% | | 52% | | 52% | | 52% | | 53% | | 53% | |
| Hispanic Heterosexual Men | ≥2 Phys. | 19% | 20% | 21% | | 22% | | 22% | | 23% | | 24% | | 25% | | 26% | | 27% | | 28% | | 29% | | 30% | | 31% | | 32% | | 33% | | 34% | | 35% | | 37% | | 38% | | 39% | | 40% | |
| Hispanic Heterosexual Men | ≥2 Any | 37% | 40% | 42% | | 44% | | 45% | | 47% | | 48% | | 49% | | 50% | | 52% | | 53% | | 54% | | 55% | | 56% | | 56% | | 57% | | 58% | | 59% | | 60% | | 61% | | 62% | | 63% | |
| Hispanic Heterosexual Men | ≥1 Ment. & 2 Phys. | 7% | 9% | 10% | | 10% | | 11% | | 12% | | 13% | | 13% | | 14% | | 15% | | 15% | | 16% | | 17% | | 17% | | 18% | | 19% | | 19% | | 20% | | 21% | | 22% | | 22% | | 23% | |
| White Heterosexual Women | ≥1 Ment. | 59% | 60% | 62% | | 63% | | 64% | | 66% | | 67% | | 68% | | 69% | | 69% | | 70% | | 71% | | 72% | | 72% | | 73% | | 74% | | 74% | | 75% | | 75% | | 76% | | 76% | | 77% | |
| White Heterosexual Women | ≥2 Phys. | 23% | 26% | 28% | | 29% | | 31% | | 32% | | 34% | | 35% | | 37% | | 38% | | 40% | | 41% | | 43% | | 44% | | 46% | | 47% | | 49% | | 50% | | 51% | | 53% | | 54% | | 56% | |
| White Heterosexual Women | ≥2 Any | 53% | 57% | 59% | | 60% | | 62% | | 64% | | 65% | | 67% | | 68% | | 69% | | 70% | | 71% | | 72% | | 74% | | 75% | | 76% | | 77% | | 78% | | 78% | | 79% | | 81% | | 82% | |
| White Heterosexual Women | ≥1 Ment. & 2 Phys. | 14% | 16% | 18% | | 19% | | 21% | | 22% | | 24% | | 25% | | 27% | | 28% | | 30% | | 31% | | 32% | | 34% | | 35% | | 37% | | 38% | | 40% | | 41% | | 42% | | 44% | | 45% | |
| Black/AA Heterosexual Women | ≥1 Ment. | 47% | 50% | 52% | | 54% | | 55% | | 57% | | 58% | | 58% | | 59% | | 60% | | 60% | | 61% | | 61% | | 62% | | 62% | | 63% | | 64% | | 64% | | 65% | | 65% | | 66% | | 67% | |
| Black/AA Heterosexual Women | ≥2 Phys. | 28% | 30% | 31% | | 32% | | 33% | | 34% | | 35% | | 36% | | 36% | | 37% | | 38% | | 38% | | 39% | | 39% | | 40% | | 40% | | 41% | | 42% | | 42% | | 43% | | 43% | | 44% | |
| Black/AA Heterosexual Women | ≥2 Any | 50% | 53% | 55% | | 57% | | 58% | | 59% | | 60% | | 61% | | 62% | | 63% | | 64% | | 65% | | 65% | | 66% | | 67% | | 67% | | 68% | | 69% | | 69% | | 70% | | 71% | | 72% | |
| Black/AA Heterosexual Women | ≥1 Ment. & 2 Phys. | 13% | 15% | 17% | | 18% | | 19% | | 20% | | 21% | | 22% | | 23% | | 23% | | 24% | | 25% | | 25% | | 26% | | 26% | | 27% | | 27% | | 28% | | 29% | | 29% | | 30% | | 31% | |
| Hispanic Heterosexual Women | ≥1 Ment. | 56% | 59% | 61% | | 62% | | 64% | | 65% | | 66% | | 68% | | 69% | | 70% | | 71% | | 72% | | 73% | | 75% | | 76% | | 77% | | 79% | | 80% | | 81% | | 83% | | 84% | | 86% | |
| Hispanic Heterosexual Women | ≥2 Phys. | 20% | 23% | 25% | | 26% | | 28% | | 30% | | 31% | | 33% | | 34% | | 36% | | 38% | | 39% | | 41% | | 43% | | 44% | | 46% | | 47% | | 49% | | 50% | | 52% | | 53% | | 55% | |
| Hispanic Heterosexual Women | ≥2 Any | 48% | 52% | 54% | | 57% | | 59% | | 61% | | 63% | | 65% | | 67% | | 69% | | 70% | | 72% | | 74% | | 75% | | 77% | | 78% | | 80% | | 81% | | 83% | | 84% | | 85% | | 86% | |
| Hispanic Heterosexual Women | ≥1 Ment. & 2 Phys. | 11% | 13% | 15% | | 17% | | 18% | | 20% | | 22% | | 23% | | 25% | | 26% | | 28% | | 29% | | 31% | | 33% | | 34% | | 36% | | 38% | | 40% | | 41% | | 43% | | 45% | | 47% | |

Footnotes:

AA=African American

PEARL = ProjEcting Age, multimoRbidity, and poLypharmacy in Adults with HIV

PWH=people with HIV

MSM=men who have sex with men

MWID=men who had injection drug use as their HIV acquisition risk factor

WWID=men who had injection drug use as their HIV acquisition risk factor

≥1 Ment. = Anxiety or depression or both (i.e., ≥1 of the mental comorbidities included)

≥2 Phys. = physical multimorbidity, defined as ≥2 physical comorbidities

≥2 Any = physical or mental multimorbidity, defined as ≥2 physical or mental comorbidities

≥1 Ment. & 2 Phys. = mental comorbidity and physical multimorbidity, defined as ≥1 mental comorbidity and ≥2 physical comorbidities

^a^Although these estimates are all PEARL projections, 2010 was during the calibration period (where observed NA-ACCORD data were available to inform the estimates) and 2020 and 2030 were projection periods (without observed NA-ACCORD data).

**Table S6:** PEARL-projected comorbidity and multimorbidity prevalence [95% interquartile range], by subgroup, in 2010^a^, 2020, and 2030

| **Subgroup** | **Comorbidity /**  **Multimorbidity** | **2010** | **2020** | **2030** |
| --- | --- | --- | --- | --- |
| Overall | Anxiety | 24.1% [23.9%, 24.2%] | 36.5% [36.3%, 36.7%] | 46.9% [46.4%, 47.6%] |
| Overall | Depression | 39.9% [39.7%, 40.1%] | 47.1% [47.0%, 47.3%] | 48.8% [48.3%, 49.3%] |
| Overall | CKD | 10.0% [10.0%, 10.1%] | 16.6% [16.5%, 16.7%] | 24.9% [24.3%, 25.5%] |
| Overall | Dyslipidemia | 30.2% [30.1%, 30.3%] | 27.4% [27.2%, 27.5%] | 25.7% [25.2%, 26.3%] |
| Overall | Diabetes | 11.5% [11.4%, 11.6%] | 15.6% [15.5%, 15.7%] | 24.0% [23.5%, 24.6%] |
| Overall | Hypertension | 36.7% [36.6%, 36.9%] | 35.5% [35.4%, 35.7%] | 31.4% [30.9%, 32.0%] |
| Overall | Cancer | 9.5% [9.5%, 9.6%] | 11.3% [11.2%, 11.3%] | 11.4% [11.2%, 11.6%] |
| Overall | ESLD | 1.2% [1.2%, 1.2%] | 1.3% [1.3%, 1.4%] | 1.5% [1.4%, 1.5%] |
| Overall | MI | 1.6% [1.6%, 1.6%] | 3.2% [3.2%, 3.3%] | 7.9% [7.7%, 8.2%] |
| Overall | ≥1 Ment. | 53.1% [52.9%, 53.2%] | 60.1% [59.9%, 60.3%] | 64.5% [64.0%, 65.1%] |
| Overall | ≥2 Phys. | 26.4% [26.3%, 26.6%] | 31.1% [31.0%, 31.3%] | 35.5% [34.7%, 36.5%] |
| Overall | ≥2 Any | 52.0% [51.9%, 52.2%] | 58.3% [58.1%, 58.5%] | 63.4% [62.5%, 64.5%] |
| Overall | ≥1 Ment. & ≥2 Phys. | 14.3% [14.2%, 14.4%] | 20.7% [20.5%, 20.8%] | 25.4% [24.8%, 26.1%] |
| White MSM | Anxiety | 28.8% [28.5%, 29.1%] | 44.2% [43.9%, 44.4%] | 53.8% [52.5%, 55.2%] |
| White MSM | Depression | 41.7% [41.3%, 42.0%] | 49.5% [49.2%, 49.8%] | 53.0% [51.9%, 54.4%] |
| White MSM | CKD | 8.3% [8.2%, 8.5%] | 15.9% [15.7%, 16.1%] | 26.9% [25.6%, 28.4%] |
| White MSM | Dyslipidemia | 35.1% [34.8%, 35.3%] | 31.2% [30.8%, 31.5%] | 29.4% [27.9%, 31.1%] |
| White MSM | Diabetes | 7.6% [7.5%, 7.8%] | 9.0% [8.9%, 9.2%] | 13.9% [13.2%, 14.6%] |
| White MSM | Hypertension | 31.0% [30.7%, 31.3%] | 28.9% [28.6%, 29.2%] | 26.6% [25.8%, 27.6%] |
| White MSM | Cancer | 13.7% [13.5%, 13.9%] | 14.8% [14.6%, 15.0%] | 15.8% [15.1%, 16.6%] |
| White MSM | ESLD | 1.1% [1.0%, 1.1%] | 1.2% [1.1%, 1.2%] | 1.4% [1.3%, 1.4%] |
| White MSM | MI | 1.9% [1.8%, 2.0%] | 3.7% [3.6%, 3.8%] | 10.0% [9.4%, 10.8%] |
| White MSM | ≥1 Ment. | 57.3% [57.0%, 57.6%] | 65.7% [65.4%, 66.0%] | 70.7% [69.5%, 72.0%] |
| White MSM | ≥2 Phys. | 25.7% [25.4%, 25.9%] | 29.0% [28.6%, 29.4%] | 34.8% [32.8%, 36.9%] |
| White MSM | ≥2 Any | 53.8% [53.5%, 54.1%] | 60.3% [59.9%, 60.8%] | 67.1% [64.8%, 69.7%] |
| White MSM | ≥1 Ment. & ≥2 Phys. | 14.9% [14.7%, 15.1%] | 20.5% [20.2%, 20.7%] | 26.2% [24.6%, 28.0%] |
| Black/AA MSM | Anxiety | 15.5% [15.2%, 15.8%] | 22.5% [22.2%, 22.7%] | 26.6% [26.2%, 27.2%] |
| Black/AA MSM | Depression | 35.4% [35.0%, 35.7%] | 39.8% [39.6%, 40.1%] | 39.4% [38.8%, 40.1%] |
| Black/AA MSM | CKD | 8.5% [8.3%, 8.8%] | 11.4% [11.2%, 11.6%] | 21.4% [20.6%, 22.2%] |
| Black/AA MSM | Dyslipidemia | 24.6% [24.2%, 24.9%] | 16.8% [16.6%, 17.0%] | 13.0% [12.5%, 13.5%] |
| Black/AA MSM | Diabetes | 10.3% [10.0%, 10.5%] | 15.1% [14.8%, 15.2%] | 26.9% [26.1%, 27.6%] |
| Black/AA MSM | Hypertension | 38.6% [38.2%, 39.0%] | 37.3% [37.0%, 37.6%] | 33.2% [32.6%, 33.9%] |
| Black/AA MSM | Cancer | 7.2% [7.0%, 7.4%] | 7.8% [7.6%, 7.9%] | 8.0% [7.7%, 8.2%] |
| Black/AA MSM | ESLD | 1.1% [1.0%, 1.2%] | 1.3% [1.2%, 1.3%] | 1.5% [1.4%, 1.6%] |
| Black/AA MSM | MI | 1.4% [1.3%, 1.5%] | 2.4% [2.4%, 2.5%] | 5.9% [5.6%, 6.2%] |
| Black/AA MSM | ≥1 Ment. | 44.4% [44.1%, 44.8%] | 48.5% [48.2%, 48.8%] | 49.5% [48.9%, 50.2%] |
| Black/AA MSM | ≥2 Phys. | 22.7% [22.4%, 23.1%] | 24.1% [23.8%, 24.4%] | 29.3% [28.4%, 30.4%] |
| Black/AA MSM | ≥2 Any | 43.5% [43.2%, 43.9%] | 44.6% [44.2%, 44.9%] | 48.2% [47.0%, 49.4%] |
| Black/AA MSM | ≥1 Ment. & ≥2 Phys. | 10.6% [10.4%, 10.8%] | 14.0% [13.8%, 14.2%] | 17.4% [16.7%, 18.1%] |
| Hispanic MSM | Anxiety | 29.2% [28.8%, 29.6%] | 43.2% [42.8%, 43.6%] | 53.0% [51.8%, 54.2%] |
| Hispanic MSM | Depression | 43.0% [42.6%, 43.4%] | 50.9% [50.5%, 51.3%] | 53.8% [52.5%, 55.3%] |
| Hispanic MSM | CKD | 7.7% [7.5%, 8.0%] | 9.3% [9.1%, 9.5%] | 13.1% [12.3%, 13.8%] |
| Hispanic MSM | Dyslipidemia | 30.1% [29.7%, 30.5%] | 29.3% [28.9%, 29.7%] | 31.9% [30.5%, 33.3%] |
| Hispanic MSM | Diabetes | 9.1% [8.9%, 9.3%] | 8.4% [8.2%, 8.6%] | 10.8% [10.4%, 11.3%] |
| Hispanic MSM | Hypertension | 22.3% [21.8%, 22.7%] | 18.5% [18.3%, 18.8%] | 15.1% [14.6%, 15.6%] |
| Hispanic MSM | Cancer | 10.9% [10.6%, 11.1%] | 10.0% [9.8%, 10.2%] | 9.4% [9.1%, 9.7%] |
| Hispanic MSM | ESLD | 1.1% [1.0%, 1.2%] | 1.1% [1.0%, 1.2%] | 1.2% [1.1%, 1.2%] |
| Hispanic MSM | MI | 1.4% [1.3%, 1.5%] | 2.1% [2.0%, 2.1%] | 4.4% [4.1%, 4.8%] |
| Hispanic MSM | ≥1 Ment. | 57.5% [57.1%, 57.9%] | 62.9% [62.6%, 63.3%] | 67.5% [66.3%, 68.8%] |
| Hispanic MSM | ≥2 Phys. | 19.5% [19.2%, 19.9%] | 19.7% [19.3%, 20.0%] | 21.3% [20.0%, 22.5%] |
| Hispanic MSM | ≥2 Any | 48.4% [47.9%, 48.8%] | 52.9% [52.4%, 53.3%] | 58.3% [56.4%, 60.4%] |
| Hispanic MSM | ≥1 Ment. & ≥2 Phys. | 11.7% [11.4%, 12.0%] | 14.5% [14.2%, 14.8%] | 16.6% [15.6%, 17.7%] |
| White MWID | Anxiety | 38.4% [37.7%, 39.2%] | 51.1% [50.4%, 51.8%] | 65.2% [63.9%, 66.7%] |
| White MWID | Depression | 39.2% [38.6%, 40.0%] | 46.3% [45.6%, 47.0%] | 51.5% [50.7%, 52.5%] |
| White MWID | CKD | 8.7% [8.3%, 9.1%] | 18.4% [17.9%, 19.0%] | 30.2% [28.7%, 31.8%] |
| White MWID | Dyslipidemia | 31.3% [30.6%, 32.0%] | 30.5% [29.7%, 31.4%] | 25.9% [24.4%, 27.7%] |
| White MWID | Diabetes | 12.2% [11.8%, 12.6%] | 19.5% [19.0%, 20.1%] | 23.9% [22.5%, 25.7%] |
| White MWID | Hypertension | 40.9% [40.1%, 41.6%] | 43.5% [42.6%, 44.3%] | 38.7% [37.3%, 40.5%] |
| White MWID | Cancer | 8.5% [8.1%, 8.9%] | 14.9% [14.4%, 15.4%] | 18.5% [17.5%, 19.9%] |
| White MWID | ESLD | 1.4% [1.2%, 1.6%] | 2.3% [2.1%, 2.5%] | 2.8% [2.5%, 3.1%] |
| White MWID | MI | 1.6% [1.4%, 1.7%] | 5.0% [4.7%, 5.3%] | 11.6% [10.6%, 13.0%] |
| White MWID | ≥1 Ment. | 62.2% [61.6%, 63.0%] | 71.0% [70.4%, 71.7%] | 79.4% [78.3%, 80.5%] |
| White MWID | ≥2 Phys. | 27.9% [27.2%, 28.4%] | 38.9% [38.2%, 39.9%] | 42.0% [39.6%, 45.0%] |
| White MWID | ≥2 Any | 58.7% [58.0%, 59.4%] | 68.6% [67.8%, 69.5%] | 74.8% [73.0%, 77.3%] |
| White MWID | ≥1 Ment. & ≥2 Phys. | 17.6% [17.0%, 18.1%] | 29.2% [28.6%, 30.1%] | 34.6% [32.6%, 37.3%] |
| Black/AA MWID | Anxiety | 21.0% [20.5%, 21.6%] | 33.9% [33.2%, 34.5%] | 55.4% [54.6%, 56.2%] |
| Black/AA MWID | Depression | 32.9% [32.3%, 33.5%] | 42.6% [41.9%, 43.3%] | 48.4% [46.9%, 50.2%] |
| Black/AA MWID | CKD | 15.3% [14.8%, 15.9%] | 25.9% [25.1%, 26.7%] | 29.0% [26.0%, 33.0%] |
| Black/AA MWID | Dyslipidemia | 24.9% [24.3%, 25.4%] | 26.7% [26.1%, 27.5%] | 23.7% [21.7%, 26.2%] |
| Black/AA MWID | Diabetes | 21.0% [20.3%, 21.5%] | 26.3% [25.7%, 27.0%] | 30.4% [28.4%, 32.9%] |
| Black/AA MWID | Hypertension | 60.1% [59.4%, 60.7%] | 62.4% [61.7%, 63.1%] | 58.0% [56.4%, 59.5%] |
| Black/AA MWID | Cancer | 8.7% [8.4%, 9.2%] | 17.8% [17.2%, 18.3%] | 22.1% [19.7%, 25.4%] |
| Black/AA MWID | ESLD | 1.6% [1.4%, 1.7%] | 2.9% [2.7%, 3.1%] | 3.4% [3.0%, 4.0%] |
| Black/AA MWID | MI | 1.7% [1.5%, 1.8%] | 6.7% [6.3%, 7.0%] | 16.3% [13.7%, 19.5%] |
| Black/AA MWID | ≥1 Ment. | 46.8% [46.2%, 47.4%] | 59.6% [59.0%, 60.3%] | 72.8% [71.7%, 74.0%] |
| Black/AA MWID | ≥2 Phys. | 39.4% [38.7%, 40.1%] | 51.6% [50.5%, 52.5%] | 52.7% [48.2%, 58.8%] |
| Black/AA MWID | ≥2 Any | 60.3% [59.7%, 60.9%] | 73.3% [72.4%, 74.1%] | 79.9% [77.2%, 83.3%] |
| Black/AA MWID | ≥1 Ment. & ≥2 Phys. | 18.6% [18.1%, 19.0%] | 31.4% [30.6%, 32.2%] | 38.5% [35.0%, 42.8%] |
| Hispanic MWID | Anxiety | 34.7% [33.7%, 35.5%] | 55.3% [54.3%, 56.2%] | 83.5% [81.9%, 85.2%] |
| Hispanic MWID | Depression | 37.4% [36.5%, 38.2%] | 50.8% [50.0%, 51.6%] | 57.8% [56.8%, 59.2%] |
| Hispanic MWID | CKD | 8.6% [8.1%, 9.1%] | 17.4% [16.7%, 18.0%] | 25.1% [22.6%, 28.2%] |
| Hispanic MWID | Dyslipidemia | 24.9% [24.2%, 25.7%] | 29.4% [28.5%, 30.3%] | 27.3% [24.8%, 29.9%] |
| Hispanic MWID | Diabetes | 24.2% [23.4%, 24.9%] | 28.0% [27.0%, 29.1%] | 43.4% [42.0%, 45.2%] |
| Hispanic MWID | Hypertension | 46.4% [45.4%, 47.2%] | 48.0% [46.8%, 48.9%] | 50.1% [48.4%, 51.8%] |
| Hispanic MWID | Cancer | 8.6% [8.1%, 9.1%] | 15.9% [15.3%, 16.6%] | 30.5% [29.6%, 31.4%] |
| Hispanic MWID | ESLD | 1.7% [1.5%, 1.9%] | 3.3% [3.0%, 3.6%] | 3.4% [2.9%, 4.1%] |
| Hispanic MWID | MI | 1.6% [1.4%, 1.8%] | 6.2% [5.7%, 6.5%] | 15.4% [13.3%, 18.1%] |
| Hispanic MWID | ≥1 Ment. | 58.1% [57.2%, 59.0%] | 71.0% [70.1%, 71.8%] | 86.9% [85.8%, 88.1%] |
| Hispanic MWID | ≥2 Phys. | 32.7% [31.7%, 33.5%] | 43.9% [42.5%, 45.2%] | 56.6% [53.7%, 60.1%] |
| Hispanic MWID | ≥2 Any | 60.3% [59.3%, 61.2%] | 74.0% [72.7%, 75.1%] | 87.6% [85.8%, 89.9%] |
| Hispanic MWID | ≥1 Ment. & ≥2 Phys. | 19.3% [18.6%, 20.1%] | 32.3% [31.2%, 33.4%] | 50.1% [47.2%, 53.2%] |
| White WWID | Anxiety | 39.6% [38.6%, 40.6%] | 60.7% [59.9%, 61.7%] | 72.2% [71.3%, 73.2%] |
| White WWID | Depression | 51.8% [50.6%, 52.9%] | 54.4% [53.5%, 55.4%] | 49.5% [48.4%, 50.6%] |
| White WWID | CKD | 17.8% [16.9%, 18.6%] | 27.9% [27.1%, 28.7%] | 36.2% [34.9%, 37.7%] |
| White WWID | Dyslipidemia | 21.7% [20.9%, 22.8%] | 24.3% [23.4%, 25.3%] | 22.1% [21.2%, 23.0%] |
| White WWID | Diabetes | 14.4% [13.6%, 15.2%] | 23.2% [22.3%, 24.1%] | 38.1% [37.3%, 39.0%] |
| White WWID | Hypertension | 27.5% [26.4%, 28.3%] | 36.7% [35.7%, 37.8%] | 37.6% [36.8%, 38.5%] |
| White WWID | Cancer | 7.6% [7.0%, 8.1%] | 10.7% [10.2%, 11.3%] | 8.1% [7.6%, 8.5%] |
| White WWID | ESLD | 1.5% [1.2%, 1.8%] | 2.5% [2.1%, 2.8%] | 3.0% [2.7%, 3.4%] |
| White WWID | MI | 1.5% [1.3%, 1.8%] | 3.5% [3.1%, 3.8%] | 8.6% [8.0%, 9.3%] |
| White WWID | ≥1 Ment. | 70.2% [69.3%, 71.3%] | 78.7% [78.0%, 79.6%] | 82.6% [81.6%, 83.5%] |
| White WWID | ≥2 Phys. | 23.0% [22.2%, 23.8%] | 37.4% [36.4%, 38.4%] | 45.6% [44.4%, 46.9%] |
| White WWID | ≥2 Any | 59.1% [58.1%, 60.2%] | 74.3% [73.5%, 75.2%] | 79.2% [77.9%, 80.2%] |
| White WWID | ≥1 Ment. & ≥2 Phys. | 16.2% [15.6%, 17.1%] | 30.2% [29.3%, 31.1%] | 38.8% [37.6%, 40.0%] |
| Black/AA WWID | Anxiety | 23.7% [23.0%, 24.3%] | 46.0% [45.1%, 46.9%] | 59.4% [58.0%, 61.2%] |
| Black/AA WWID | Depression | 49.9% [49.0%, 50.7%] | 55.7% [54.6%, 56.6%] | 53.8% [51.6%, 56.1%] |
| Black/AA WWID | CKD | 21.8% [21.3%, 22.6%] | 68.6% [67.8%, 69.3%] | 90.6% [89.8%, 91.6%] |
| Black/AA WWID | Dyslipidemia | 27.5% [26.8%, 28.2%] | 31.4% [30.4%, 32.4%] | 30.5% [28.0%, 33.9%] |
| Black/AA WWID | Diabetes | 14.7% [14.1%, 15.2%] | 29.0% [28.2%, 29.7%] | 48.4% [46.9%, 50.1%] |
| Black/AA WWID | Hypertension | 58.5% [57.6%, 59.2%] | 59.2% [58.3%, 60.0%] | 55.5% [53.3%, 58.2%] |
| Black/AA WWID | Cancer | 7.5% [7.1%, 8.0%] | 10.7% [10.2%, 11.2%] | 7.6% [7.0%, 8.5%] |
| Black/AA WWID | ESLD | 1.4% [1.3%, 1.6%] | 2.1% [1.9%, 2.3%] | 2.2% [1.9%, 2.5%] |
| Black/AA WWID | MI | 1.6% [1.4%, 1.9%] | 5.8% [5.5%, 6.2%] | 18.4% [16.7%, 21.2%] |
| Black/AA WWID | ≥1 Ment. | 61.5% [60.7%, 62.3%] | 74.7% [74.0%, 75.5%] | 79.3% [77.8%, 81.3%] |
| Black/AA WWID | ≥2 Phys. | 39.7% [38.7%, 40.5%] | 67.2% [66.0%, 68.3%] | 78.2% [75.9%, 81.3%] |
| Black/AA WWID | ≥2 Any | 67.4% [66.5%, 68.1%] | 87.0% [86.2%, 87.8%] | 93.4% [92.0%, 95.2%] |
| Black/AA WWID | ≥1 Ment. & ≥2 Phys. | 24.4% [23.6%, 25.1%] | 50.6% [49.4%, 51.6%] | 62.8% [60.1%, 66.2%] |
| Hispanic WWID | Anxiety | 39.2% [37.8%, 41.4%] | 60.0% [58.6%, 61.9%] | 70.3% [68.3%, 73.6%] |
| Hispanic WWID | Depression | 51.6% [50.1%, 53.0%] | 55.5% [54.1%, 57.1%] | 51.3% [48.6%, 55.5%] |
| Hispanic WWID | CKD | 18.5% [17.4%, 19.6%] | 35.7% [33.7%, 37.6%] | 46.6% [41.5%, 54.7%] |
| Hispanic WWID | Dyslipidemia | 22.1% [20.7%, 23.1%] | 27.8% [26.0%, 29.3%] | 26.9% [24.1%, 31.0%] |
| Hispanic WWID | Diabetes | 14.3% [13.2%, 15.3%] | 23.5% [22.3%, 24.9%] | 37.6% [36.1%, 40.1%] |
| Hispanic WWID | Hypertension | 27.8% [26.6%, 29.2%] | 37.4% [36.0%, 38.8%] | 38.0% [36.6%, 39.2%] |
| Hispanic WWID | Cancer | 7.7% [6.9%, 8.4%] | 11.5% [10.3%, 12.6%] | 8.8% [7.7%, 10.2%] |
| Hispanic WWID | ESLD | 1.5% [1.1%, 1.9%] | 2.1% [1.8%, 2.7%] | 2.6% [2.2%, 3.1%] |
| Hispanic WWID | MI | 1.6% [1.1%, 2.0%] | 4.3% [3.7%, 5.0%] | 11.7% [9.9%, 14.4%] |
| Hispanic WWID | ≥1 Ment. | 70.0% [68.7%, 71.2%] | 79.1% [77.7%, 80.5%] | 82.2% [80.3%, 85.3%] |
| Hispanic WWID | ≥2 Phys. | 23.6% [22.2%, 24.9%] | 42.6% [40.7%, 44.4%] | 51.8% [47.8%, 57.8%] |
| Hispanic WWID | ≥2 Any | 59.3% [57.8%, 60.9%] | 77.3% [75.8%, 78.8%] | 81.7% [78.4%, 86.9%] |
| Hispanic WWID | ≥1 Ment. & ≥2 Phys. | 16.6% [15.5%, 17.8%] | 34.2% [32.5%, 36.1%] | 43.7% [39.8%, 49.9%] |
| White Heterosexual Men | Anxiety | 14.9% [13.8%, 16.1%] | 23.3% [22.3%, 24.3%] | 36.4% [35.3%, 37.6%] |
| White Heterosexual Men | Depression | 31.1% [29.7%, 32.3%] | 33.0% [32.1%, 34.1%] | 29.5% [28.2%, 31.1%] |
| White Heterosexual Men | CKD | 9.4% [8.3%, 10.4%] | 15.7% [14.9%, 16.4%] | 27.3% [25.7%, 29.2%] |
| White Heterosexual Men | Dyslipidemia | 30.5% [29.0%, 31.9%] | 24.6% [23.7%, 25.6%] | 26.0% [24.7%, 27.4%] |
| White Heterosexual Men | Diabetes | 10.2% [9.2%, 11.3%] | 14.5% [13.7%, 15.2%] | 25.4% [24.4%, 26.2%] |
| White Heterosexual Men | Hypertension | 31.6% [30.2%, 33.1%] | 34.8% [33.7%, 35.9%] | 35.3% [33.7%, 36.8%] |
| White Heterosexual Men | Cancer | 8.5% [7.6%, 9.5%] | 11.2% [10.5%, 12.1%] | 13.1% [12.2%, 14.0%] |
| White Heterosexual Men | ESLD | 1.1% [0.8%, 1.5%] | 0.9% [0.7%, 1.1%] | 0.7% [0.6%, 1.0%] |
| White Heterosexual Men | MI | 1.5% [1.1%, 1.8%] | 2.6% [2.2%, 3.0%] | 5.2% [4.6%, 6.0%] |
| White Heterosexual Men | ≥1 Ment. | 40.5% [38.9%, 41.8%] | 43.0% [41.9%, 44.2%] | 47.6% [46.5%, 49.0%] |
| White Heterosexual Men | ≥2 Phys. | 23.2% [22.0%, 24.4%] | 28.5% [27.3%, 29.7%] | 38.9% [36.7%, 41.0%] |
| White Heterosexual Men | ≥2 Any | 41.9% [40.5%, 43.6%] | 47.8% [46.6%, 48.9%] | 58.3% [56.2%, 60.9%] |
| White Heterosexual Men | ≥1 Ment. & ≥2 Phys. | 9.7% [8.8%, 10.6%] | 13.8% [13.0%, 14.7%] | 19.9% [18.6%, 21.4%] |
| Black/AA Heterosexual Men | Anxiety | 7.7% [7.4%, 8.1%] | 13.5% [13.2%, 14.0%] | 14.4% [14.0%, 15.0%] |
| Black/AA Heterosexual Men | Depression | 24.9% [24.4%, 25.5%] | 33.2% [32.6%, 33.8%] | 37.2% [35.8%, 39.1%] |
| Black/AA Heterosexual Men | CKD | 10.0% [9.6%, 10.5%] | 17.4% [17.0%, 17.8%] | 29.7% [27.6%, 32.3%] |
| Black/AA Heterosexual Men | Dyslipidemia | 27.5% [26.9%, 28.1%] | 26.3% [25.7%, 26.9%] | 27.2% [25.9%, 28.9%] |
| Black/AA Heterosexual Men | Diabetes | 13.8% [13.3%, 14.2%] | 18.7% [18.2%, 19.1%] | 29.0% [27.9%, 30.7%] |
| Black/AA Heterosexual Men | Hypertension | 45.8% [45.1%, 46.4%] | 48.7% [48.3%, 49.3%] | 45.4% [44.4%, 46.7%] |
| Black/AA Heterosexual Men | Cancer | 8.3% [7.9%, 8.6%] | 10.1% [9.8%, 10.5%] | 12.1% [11.4%, 12.9%] |
| Black/AA Heterosexual Men | ESLD | 1.2% [1.1%, 1.4%] | 1.2% [1.1%, 1.3%] | 1.0% [0.8%, 1.1%] |
| Black/AA Heterosexual Men | MI | 1.4% [1.3%, 1.6%] | 2.7% [2.5%, 2.8%] | 6.3% [5.7%, 7.2%] |
| Black/AA Heterosexual Men | ≥1 Ment. | 30.2% [29.6%, 30.8%] | 39.3% [38.7%, 39.8%] | 42.7% [41.5%, 44.4%] |
| Black/AA Heterosexual Men | ≥2 Phys. | 29.0% [28.4%, 29.6%] | 36.2% [35.6%, 36.8%] | 45.1% [42.6%, 48.2%] |
| Black/AA Heterosexual Men | ≥2 Any | 42.7% [42.0%, 43.3%] | 52.4% [51.7%, 53.0%] | 59.9% [57.6%, 63.3%] |
| Black/AA Heterosexual Men | ≥1 Ment. & ≥2 Phys. | 8.9% [8.4%, 9.3%] | 15.2% [14.7%, 15.6%] | 20.8% [19.3%, 22.7%] |
| Hispanic Heterosexual Men | Anxiety | 15.2% [14.5%, 16.1%] | 25.8% [24.9%, 26.7%] | 38.5% [37.0%, 39.9%] |
| Hispanic Heterosexual Men | Depression | 35.0% [34.1%, 35.8%] | 45.2% [44.0%, 46.2%] | 42.1% [39.7%, 45.4%] |
| Hispanic Heterosexual Men | CKD | 9.3% [8.8%, 10.0%] | 16.0% [15.3%, 16.7%] | 27.6% [25.8%, 30.4%] |
| Hispanic Heterosexual Men | Dyslipidemia | 29.4% [28.4%, 30.4%] | 26.9% [26.1%, 27.7%] | 27.4% [25.8%, 29.4%] |
| Hispanic Heterosexual Men | Diabetes | 10.3% [9.6%, 11.0%] | 15.7% [15.1%, 16.4%] | 25.7% [24.6%, 27.1%] |
| Hispanic Heterosexual Men | Hypertension | 25.3% [24.3%, 26.4%] | 33.8% [32.8%, 34.7%] | 35.9% [34.0%, 38.5%] |
| Hispanic Heterosexual Men | Cancer | 8.3% [7.7%, 9.0%] | 10.8% [10.3%, 11.5%] | 12.9% [12.1%, 14.0%] |
| Hispanic Heterosexual Men | ESLD | 1.2% [0.9%, 1.4%] | 1.0% [0.9%, 1.2%] | 0.8% [0.7%, 1.0%] |
| Hispanic Heterosexual Men | MI | 1.4% [1.2%, 1.7%] | 2.5% [2.2%, 2.8%] | 6.1% [5.2%, 7.0%] |
| Hispanic Heterosexual Men | ≥1 Ment. | 42.0% [41.0%, 43.2%] | 49.7% [48.7%, 50.9%] | 52.9% [50.8%, 55.4%] |
| Hispanic Heterosexual Men | ≥2 Phys. | 20.1% [19.1%, 21.0%] | 29.3% [28.2%, 30.3%] | 39.8% [37.2%, 43.6%] |
| Hispanic Heterosexual Men | ≥2 Any | 40.5% [39.3%, 41.6%] | 53.6% [52.2%, 54.7%] | 63.0% [60.2%, 67.2%] |
| Hispanic Heterosexual Men | ≥1 Ment. & ≥2 Phys. | 8.6% [8.0%, 9.3%] | 16.1% [15.3%, 16.9%] | 22.9% [21.0%, 25.6%] |
| White Heterosexual Women | Anxiety | 32.3% [31.5%, 33.1%] | 49.9% [49.4%, 50.6%] | 60.2% [59.1%, 61.3%] |
| White Heterosexual Women | Depression | 43.6% [42.9%, 44.5%] | 56.5% [55.8%, 57.3%] | 62.7% [61.1%, 64.3%] |
| White Heterosexual Women | CKD | 11.1% [10.5%, 11.7%] | 33.7% [33.0%, 34.4%] | 64.0% [61.1%, 67.1%] |
| White Heterosexual Women | Dyslipidemia | 40.2% [39.4%, 40.9%] | 36.7% [35.9%, 37.5%] | 33.4% [31.5%, 35.5%] |
| White Heterosexual Women | Diabetes | 10.8% [10.2%, 11.3%] | 20.9% [20.3%, 21.4%] | 36.1% [34.9%, 37.1%] |
| White Heterosexual Women | Hypertension | 29.0% [28.2%, 29.7%] | 31.7% [31.0%, 32.2%] | 29.1% [28.1%, 30.1%] |
| White Heterosexual Women | Cancer | 6.8% [6.4%, 7.2%] | 9.5% [9.1%, 9.8%] | 8.3% [7.9%, 8.8%] |
| White Heterosexual Women | ESLD | 1.2% [1.0%, 1.4%] | 1.1% [1.0%, 1.3%] | 1.3% [1.1%, 1.4%] |
| White Heterosexual Women | MI | 1.5% [1.3%, 1.7%] | 3.4% [3.1%, 3.6%] | 8.7% [7.7%, 9.4%] |
| White Heterosexual Women | ≥1 Ment. | 60.4% [59.6%, 61.2%] | 71.0% [70.4%, 71.7%] | 76.9% [75.7%, 77.8%] |
| White Heterosexual Women | ≥2 Phys. | 26.0% [25.3%, 26.7%] | 41.1% [40.3%, 41.9%] | 56.0% [53.2%, 58.6%] |
| White Heterosexual Women | ≥2 Any | 56.6% [55.8%, 57.3%] | 71.4% [70.6%, 72.2%] | 81.5% [79.3%, 83.4%] |
| White Heterosexual Women | ≥1 Ment. & ≥2 Phys. | 15.9% [15.3%, 16.5%] | 31.0% [30.3%, 31.8%] | 45.4% [42.9%, 47.7%] |
| Black/AA Heterosexual Women | Anxiety | 16.4% [16.0%, 16.7%] | 30.6% [30.3%, 31.0%] | 48.8% [47.9%, 49.5%] |
| Black/AA Heterosexual Women | Depression | 41.3% [40.9%, 41.7%] | 49.8% [49.5%, 50.2%] | 48.3% [47.4%, 49.2%] |
| Black/AA Heterosexual Women | CKD | 10.6% [10.3%, 10.9%] | 13.6% [13.3%, 13.8%] | 13.9% [13.4%, 14.4%] |
| Black/AA Heterosexual Women | Dyslipidemia | 30.8% [30.3%, 31.1%] | 31.5% [31.0%, 31.9%] | 30.0% [28.6%, 31.0%] |
| Black/AA Heterosexual Women | Diabetes | 14.4% [14.1%, 14.8%] | 24.1% [23.8%, 24.4%] | 38.6% [37.7%, 39.3%] |
| Black/AA Heterosexual Women | Hypertension | 44.3% [43.9%, 44.8%] | 47.5% [47.2%, 47.9%] | 46.1% [45.5%, 46.6%] |
| Black/AA Heterosexual Women | Cancer | 6.9% [6.7%, 7.1%] | 10.2% [10.0%, 10.4%] | 9.3% [9.0%, 9.7%] |
| Black/AA Heterosexual Women | ESLD | 1.2% [1.1%, 1.3%] | 1.1% [1.1%, 1.2%] | 1.2% [1.1%, 1.3%] |
| Black/AA Heterosexual Women | MI | 1.4% [1.3%, 1.5%] | 3.3% [3.2%, 3.4%] | 10.3% [9.6%, 10.8%] |
| Black/AA Heterosexual Women | ≥1 Ment. | 50.4% [50.0%, 50.8%] | 60.9% [60.6%, 61.3%] | 66.6% [65.7%, 67.3%] |
| Black/AA Heterosexual Women | ≥2 Phys. | 29.7% [29.4%, 30.1%] | 38.2% [37.7%, 38.6%] | 44.1% [42.7%, 45.4%] |
| Black/AA Heterosexual Women | ≥2 Any | 53.2% [52.9%, 53.7%] | 64.6% [64.1%, 65.0%] | 71.7% [70.2%, 72.9%] |
| Black/AA Heterosexual Women | ≥1 Ment. & ≥2 Phys. | 15.2% [14.9%, 15.6%] | 24.6% [24.2%, 25.0%] | 30.8% [29.5%, 31.9%] |
| Hispanic Heterosexual Women | Anxiety | 24.9% [24.1%, 25.6%] | 49.1% [48.5%, 49.7%] | 78.3% [77.9%, 78.9%] |
| Hispanic Heterosexual Women | Depression | 46.2% [45.3%, 47.0%] | 58.7% [58.2%, 59.4%] | 60.5% [59.4%, 62.1%] |
| Hispanic Heterosexual Women | CKD | 10.6% [10.2%, 11.1%] | 30.2% [29.5%, 31.0%] | 59.1% [55.9%, 63.2%] |
| Hispanic Heterosexual Women | Dyslipidemia | 32.1% [31.3%, 32.8%] | 33.9% [33.1%, 34.6%] | 31.8% [29.9%, 34.3%] |
| Hispanic Heterosexual Women | Diabetes | 10.8% [10.3%, 11.3%] | 21.6% [21.1%, 22.2%] | 39.1% [38.1%, 40.3%] |
| Hispanic Heterosexual Women | Hypertension | 29.1% [28.5%, 29.9%] | 32.6% [32.0%, 33.3%] | 30.2% [29.1%, 31.5%] |
| Hispanic Heterosexual Women | Cancer | 6.9% [6.4%, 7.2%] | 10.0% [9.7%, 10.4%] | 8.5% [8.0%, 9.2%] |
| Hispanic Heterosexual Women | ESLD | 1.2% [1.0%, 1.3%] | 1.1% [0.9%, 1.2%] | 1.1% [1.0%, 1.3%] |
| Hispanic Heterosexual Women | MI | 1.4% [1.2%, 1.6%] | 3.0% [2.7%, 3.2%] | 8.0% [7.3%, 9.0%] |
| Hispanic Heterosexual Women | ≥1 Ment. | 58.6% [57.8%, 59.2%] | 72.3% [71.7%, 72.8%] | 85.7% [85.3%, 86.1%] |
| Hispanic Heterosexual Women | ≥2 Phys. | 22.8% [22.2%, 23.5%] | 39.3% [38.5%, 40.2%] | 54.7% [52.0%, 58.4%] |
| Hispanic Heterosexual Women | ≥2 Any | 51.6% [50.8%, 52.4%] | 72.1% [71.4%, 72.9%] | 86.2% [84.7%, 88.2%] |
| Hispanic Heterosexual Women | ≥1 Ment. & ≥2 Phys. | 13.5% [13.0%, 14.1%] | 29.4% [28.7%, 30.2%] | 47.0% [44.8%, 50.0%] |

Footnotes:

AA = African American

PEARL = ProjEcting Age, multimoRbidity, and poLypharmacy in Adults with HIV

PWH = people with HIV

MSM = men who have sex with men

MWID = men who had injection drug use as their HIV acquisition risk factor

WWID = men who had injection drug use as their HIV acquisition risk factor

≥1 Ment. = Anxiety or depression or both (i.e., ≥1 of the mental comorbidities included)

≥2 Phys. = physical multimorbidity, defined as ≥2 physical comorbidities

≥2 Any = physical or mental multimorbidity, defined as ≥2 physical or mental comorbidities

≥1 Ment. & 2 Phys. = mental comorbidity and physical multimorbidity, defined as ≥1 mental comorbidity and ≥2 physical comorbidities

CDK = Stage ≥3 chronic kidney disease

ESLD = End-stage liver disease

MI = myocardial infarction

^a^Although these estimates are all PEARL projections, 2010 was during the calibration period (where observed NA-ACCORD data were available to inform the estimates) and 2020 and 2030 were projection periods (without observed NA-ACCORD data).

**Figure S3a-o:** Projected prevalence (and shaded 95% credibility intervals) of individual comorbidities, by subgroup

a) White men who have sex with men

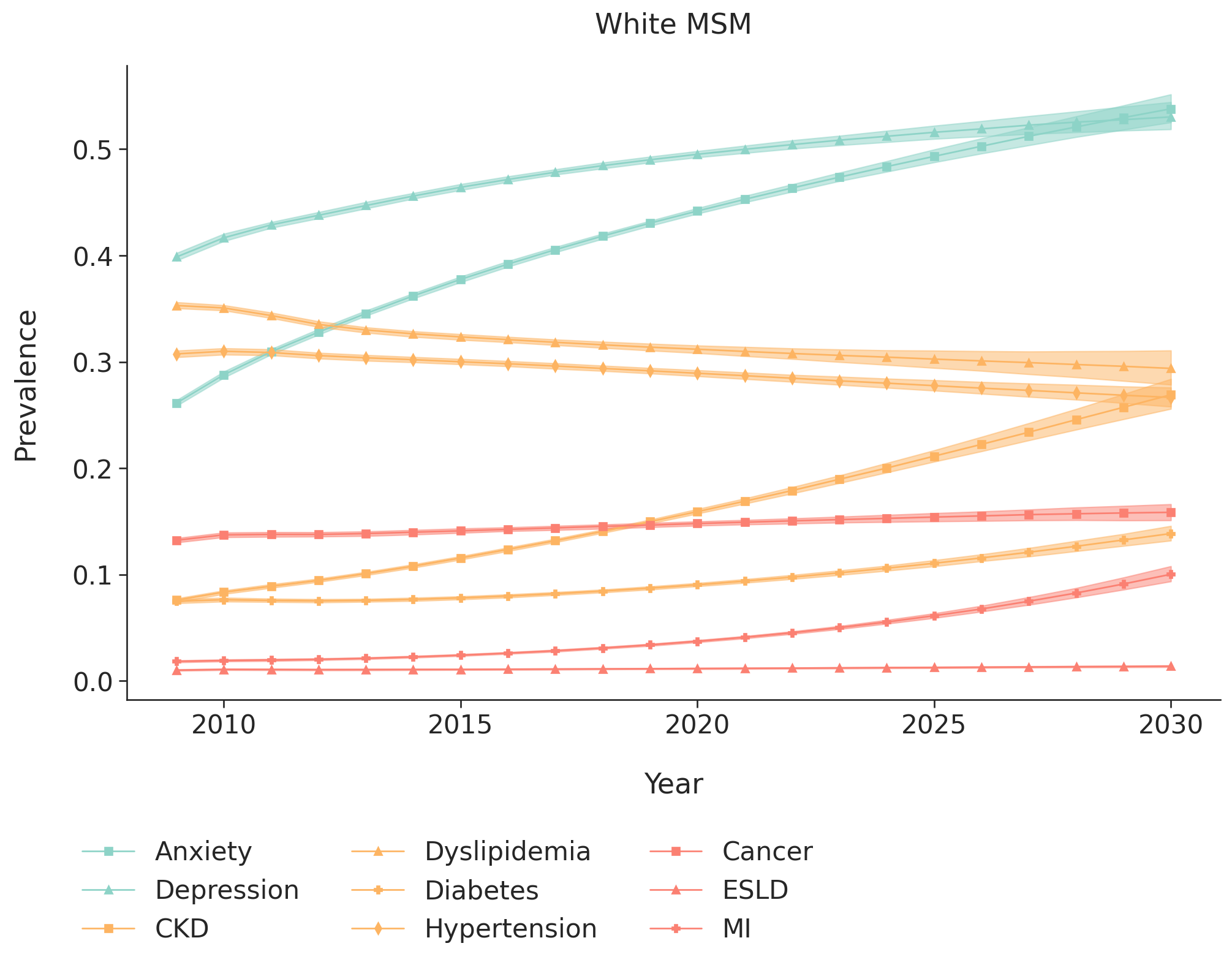

b) Black/African American men who have sex with men

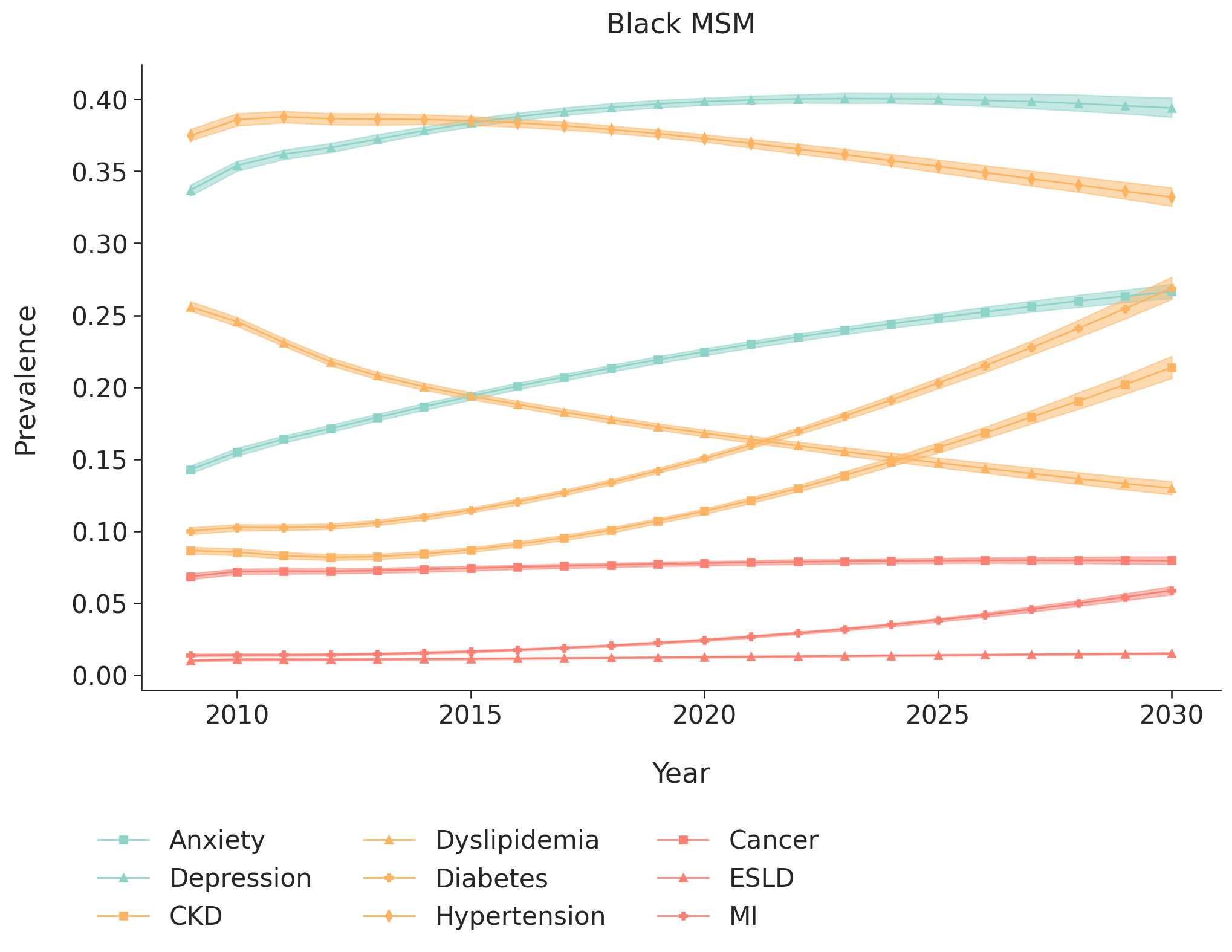

c) Hispanic men who have sex with men

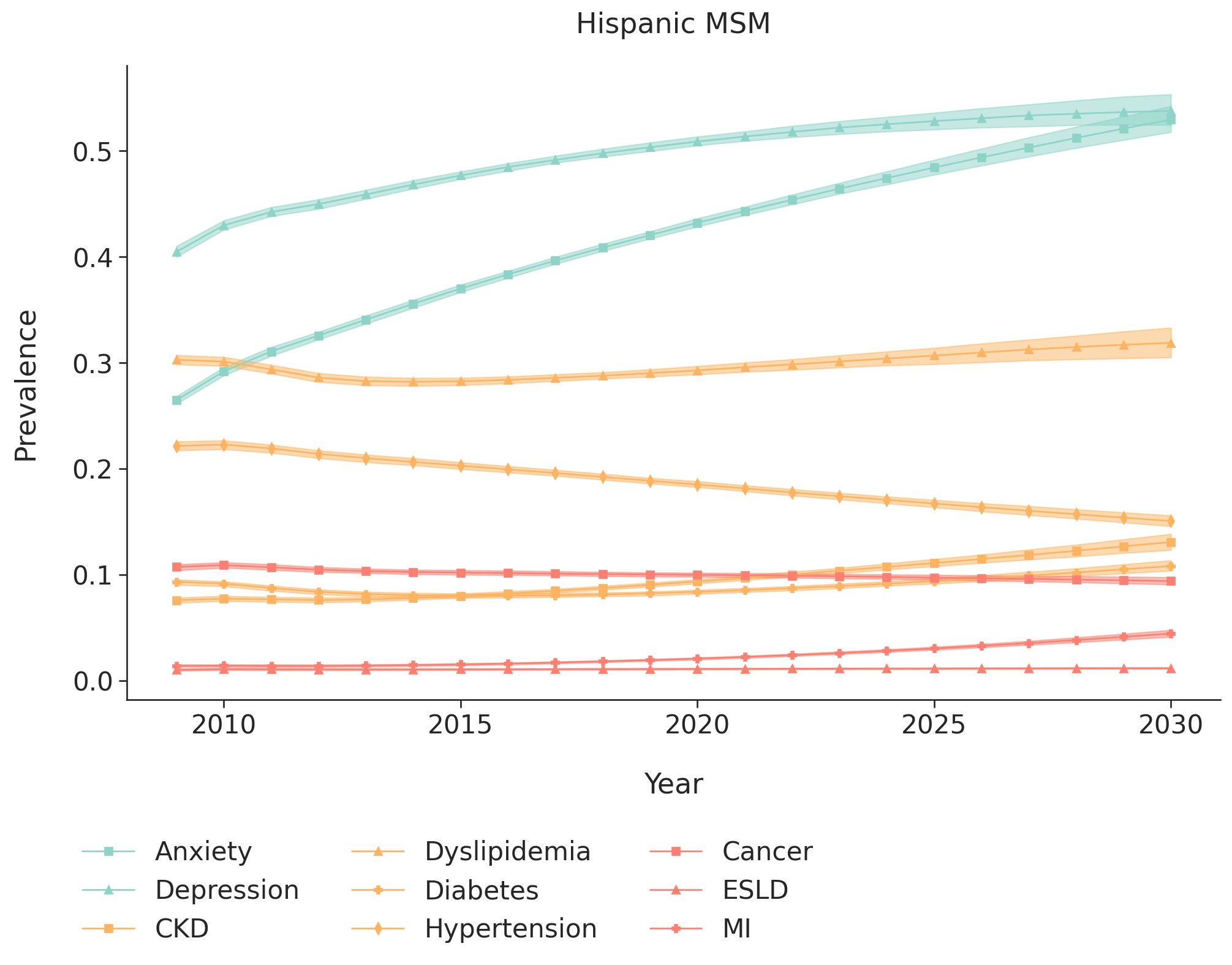

d) White men with injection drug use as their HIV acquisition risk factor

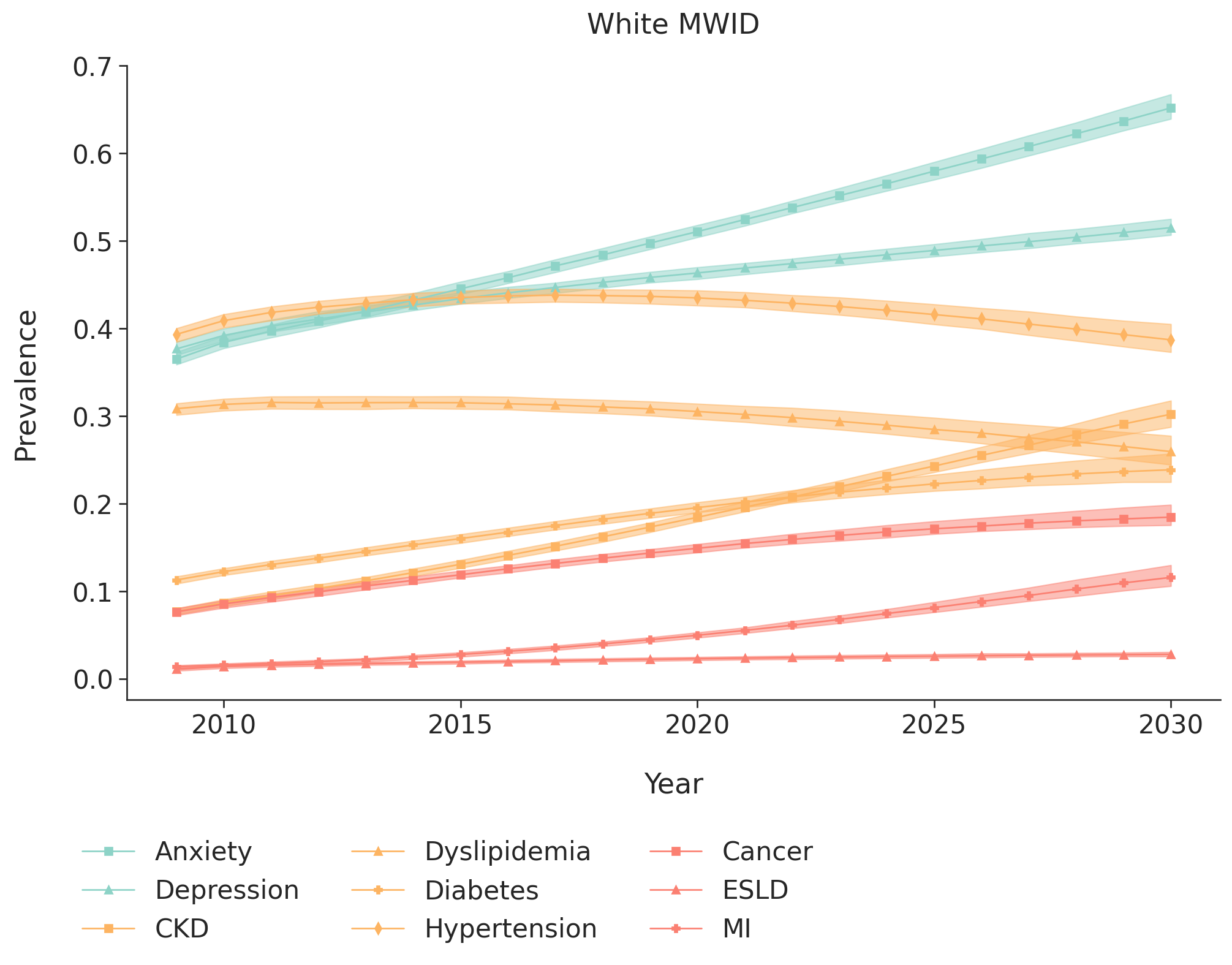

e) Black/African American men with injection drug use as their HIV acquisition risk factor

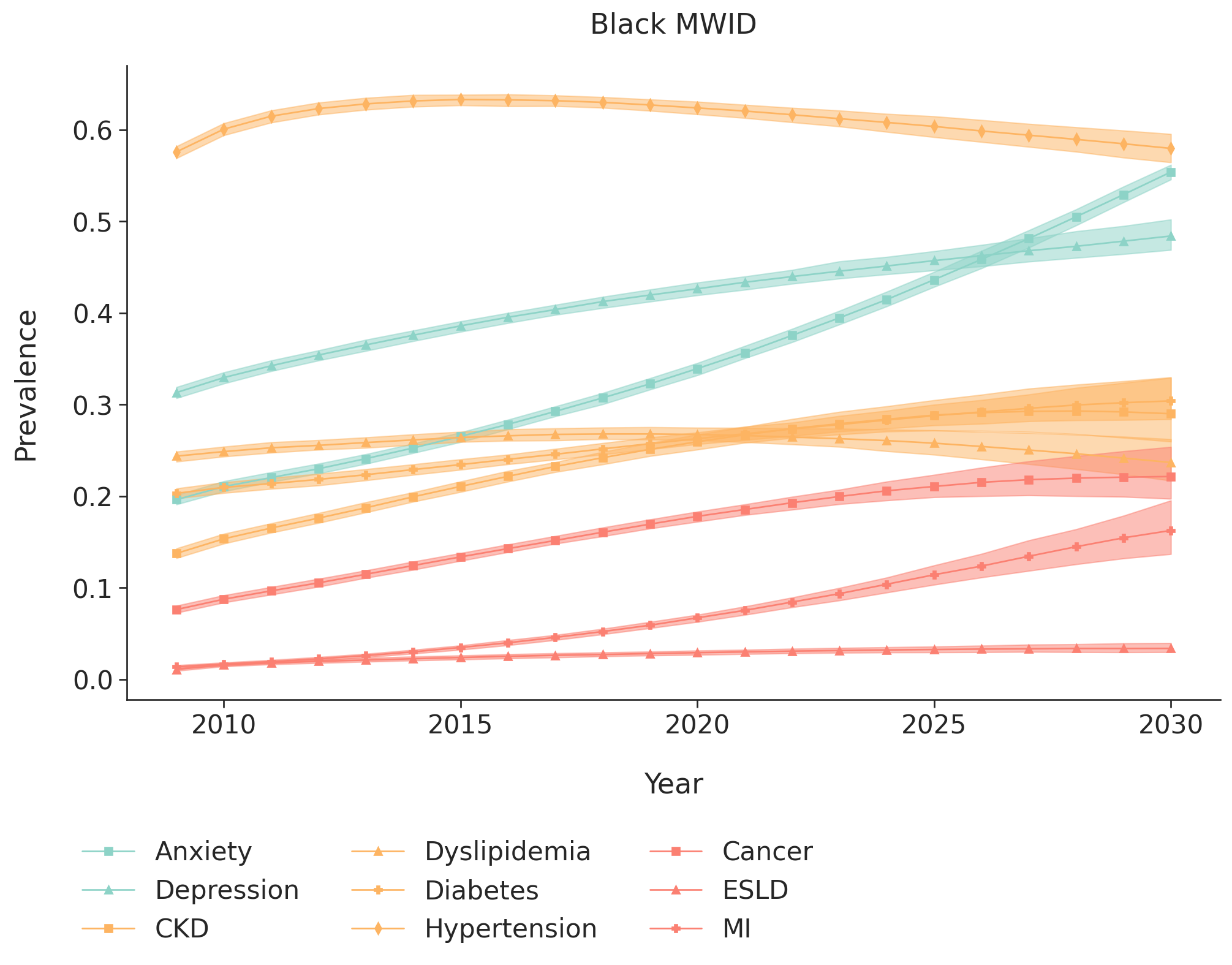

f) Hispanic men with injection drug use as their HIV acquisition risk factor

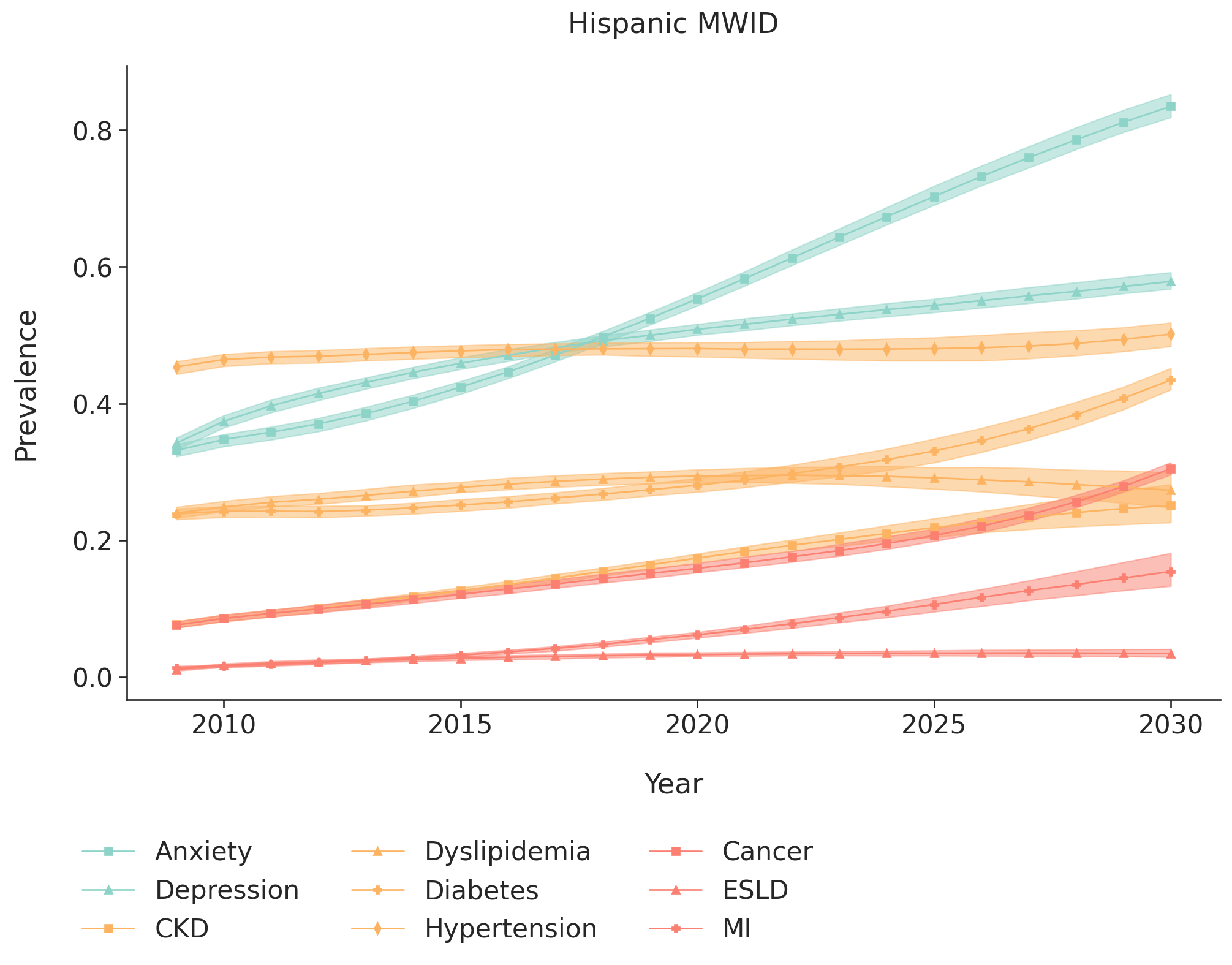

g) White women with injection drug use as their HIV acquisition risk factor

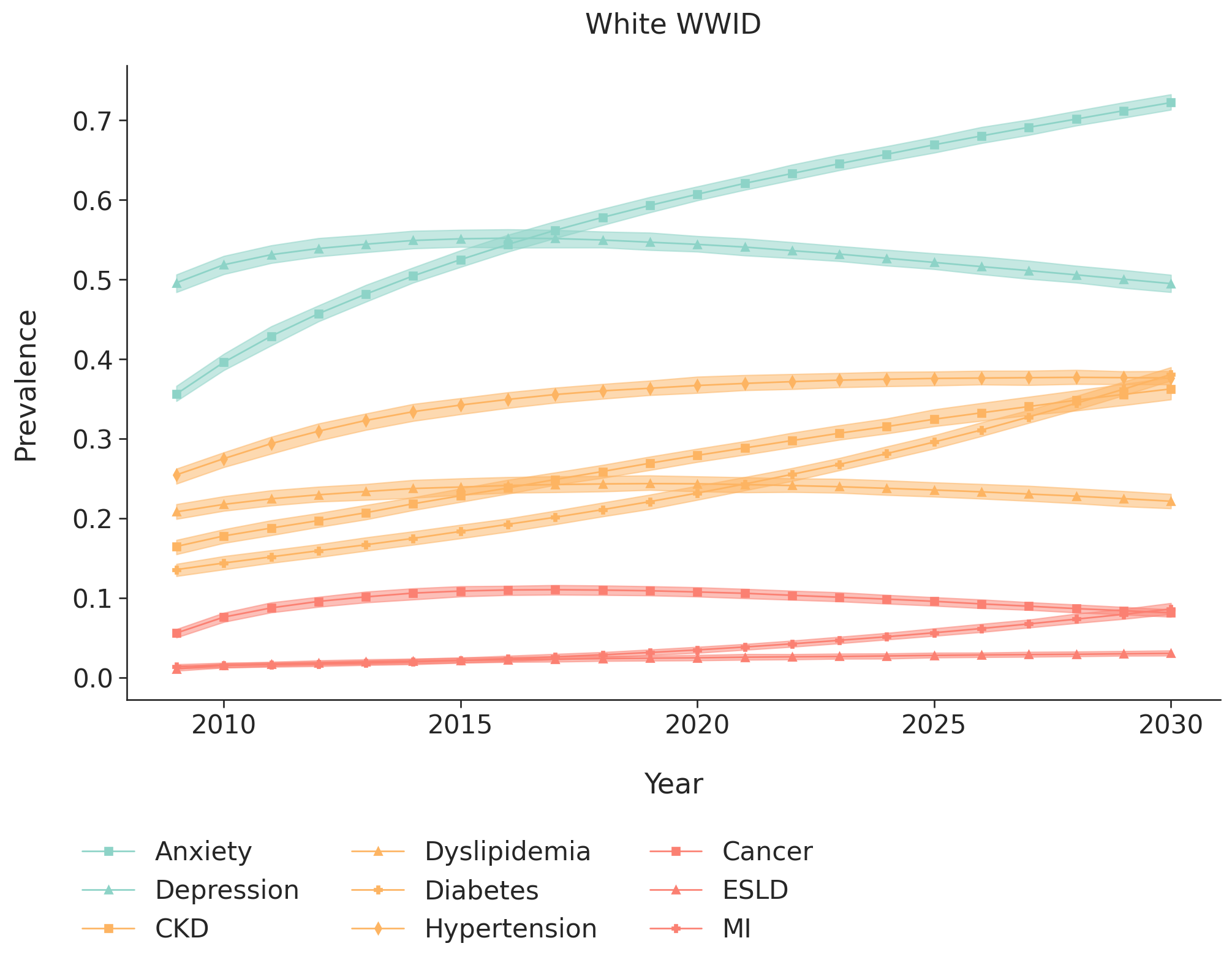

h) Black/African American women with injection drug use as their HIV acquisition risk factor

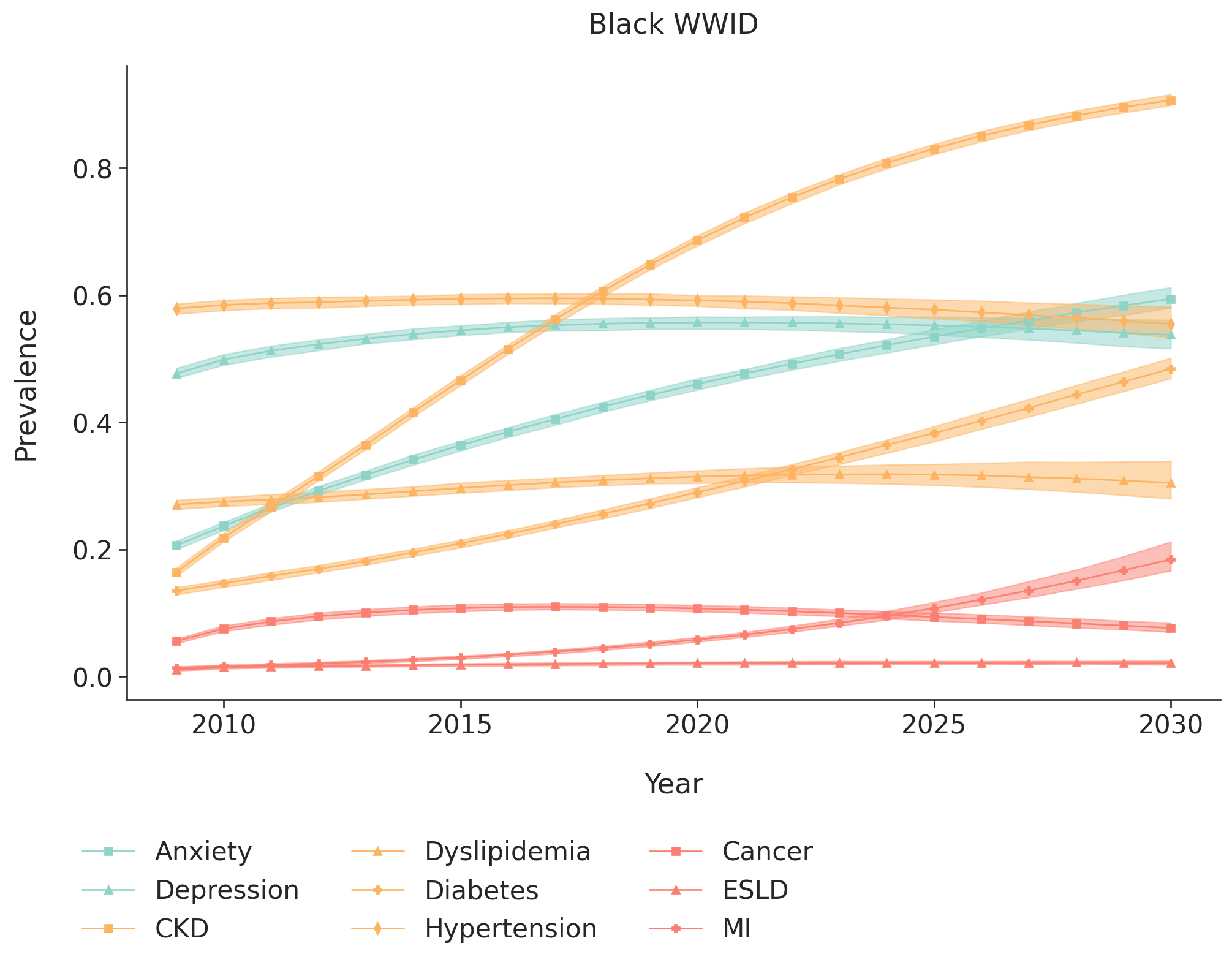

i) Hispanic women with injection drug use as their HIV acquisition risk factor

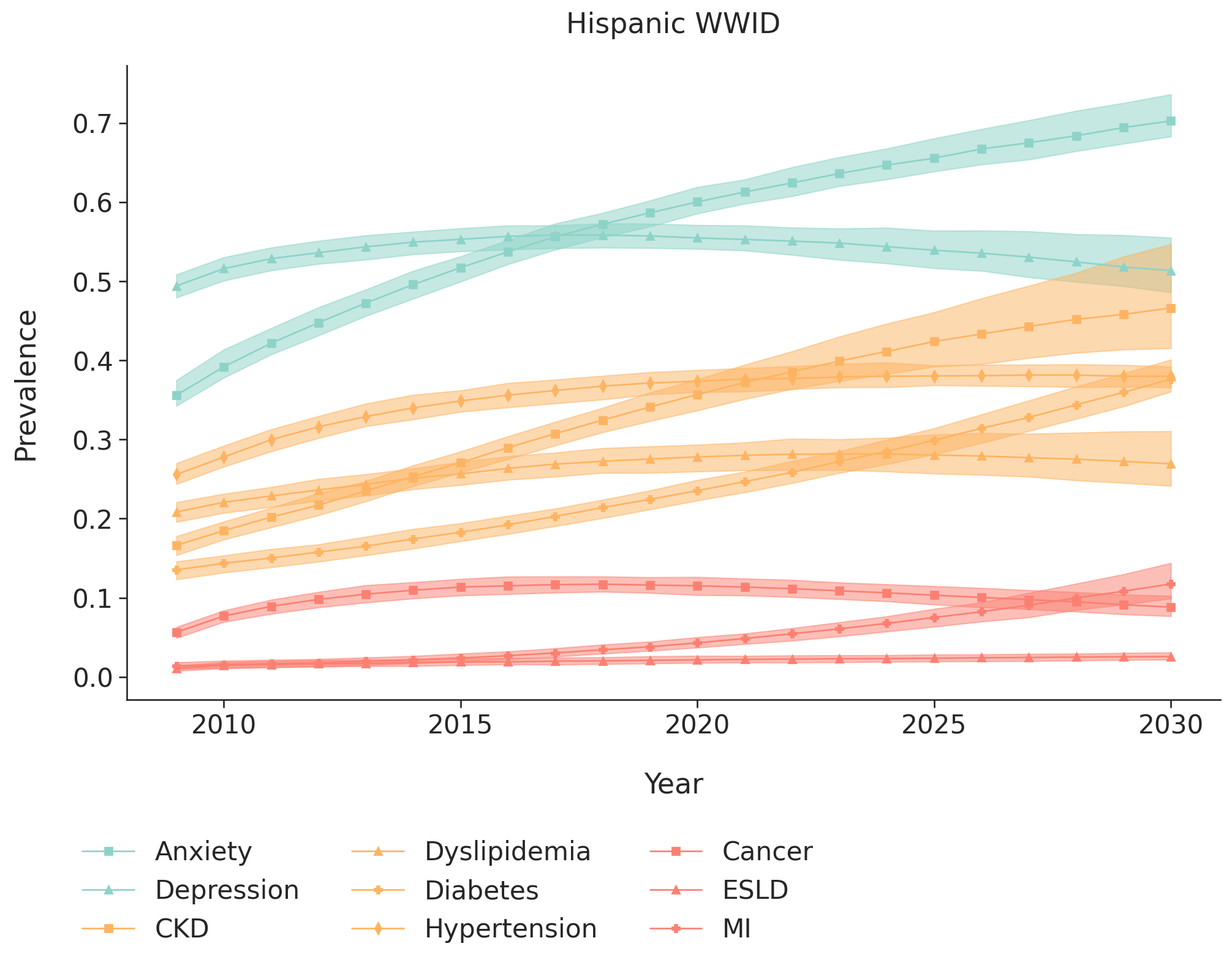

j) White heterosexual men

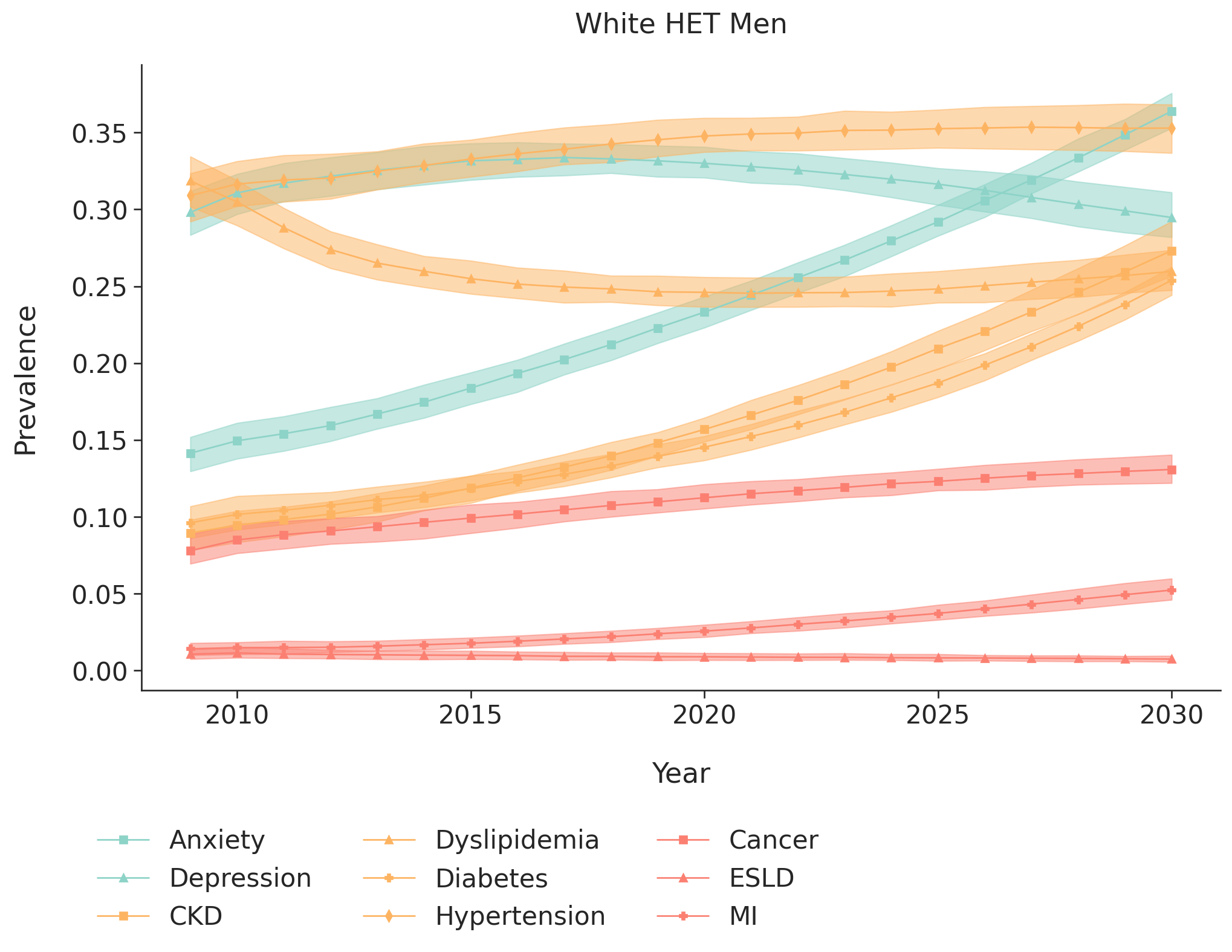

k) Black/African American heterosexual men

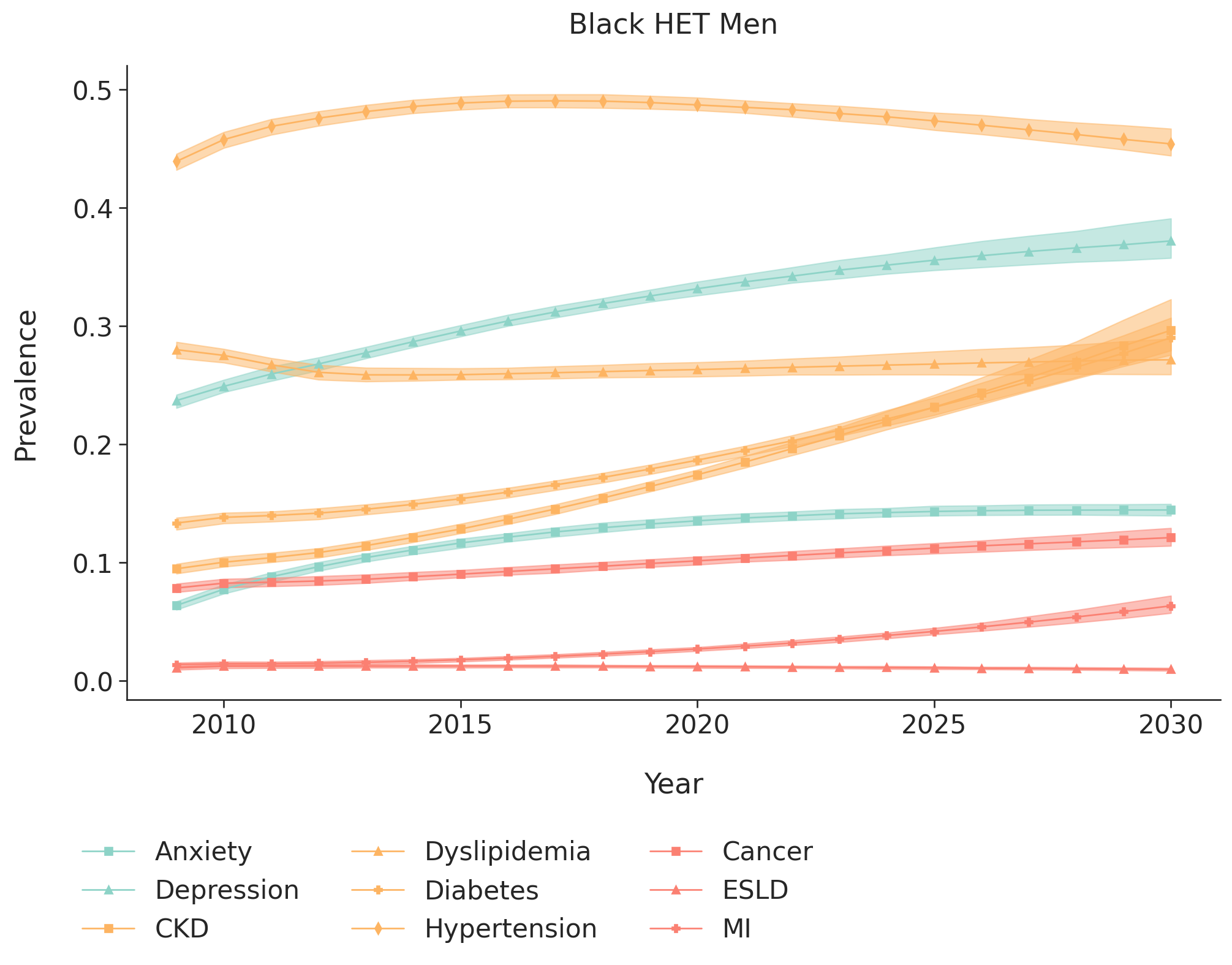

l) Hispanic heterosexual men

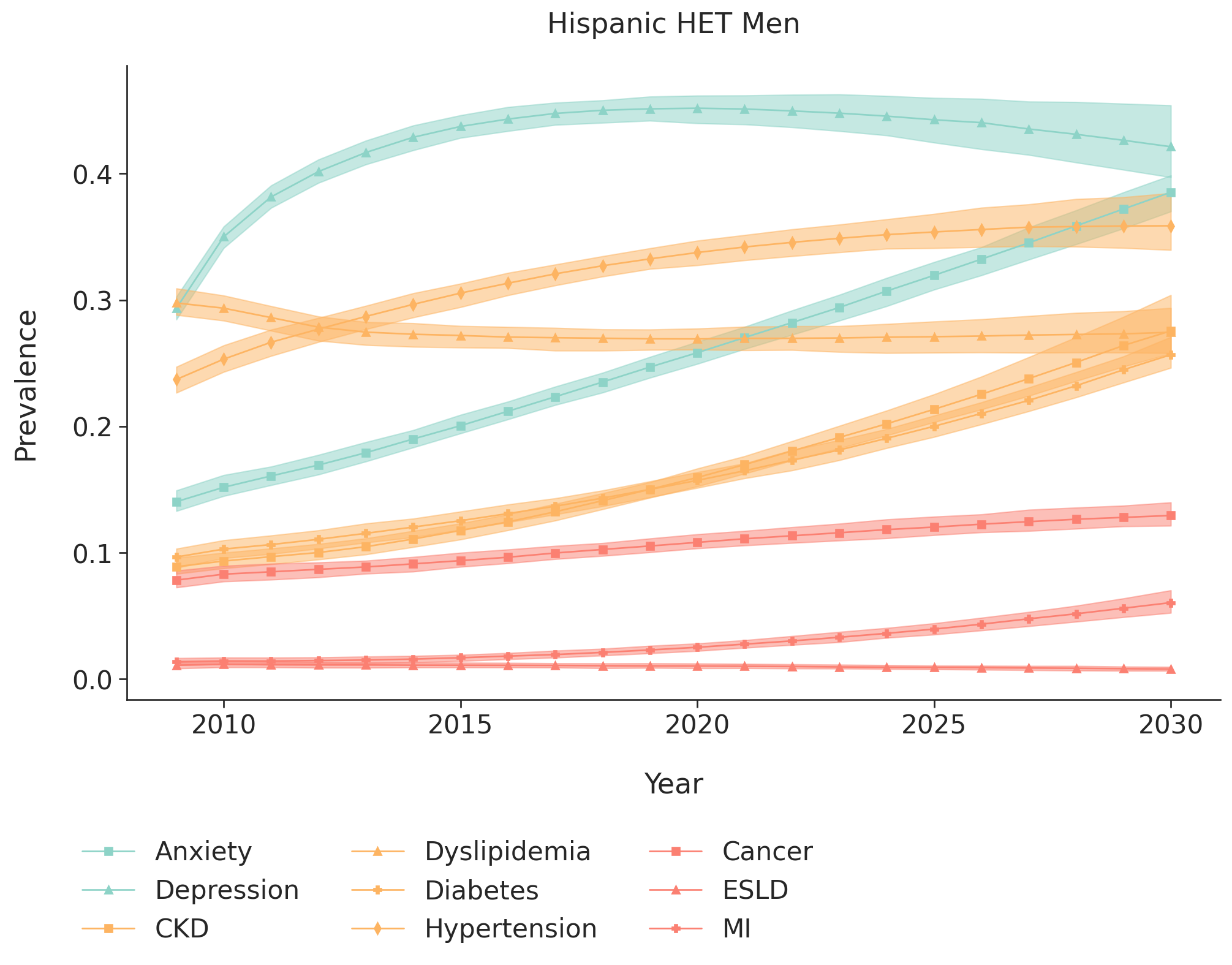

m) White heterosexual women

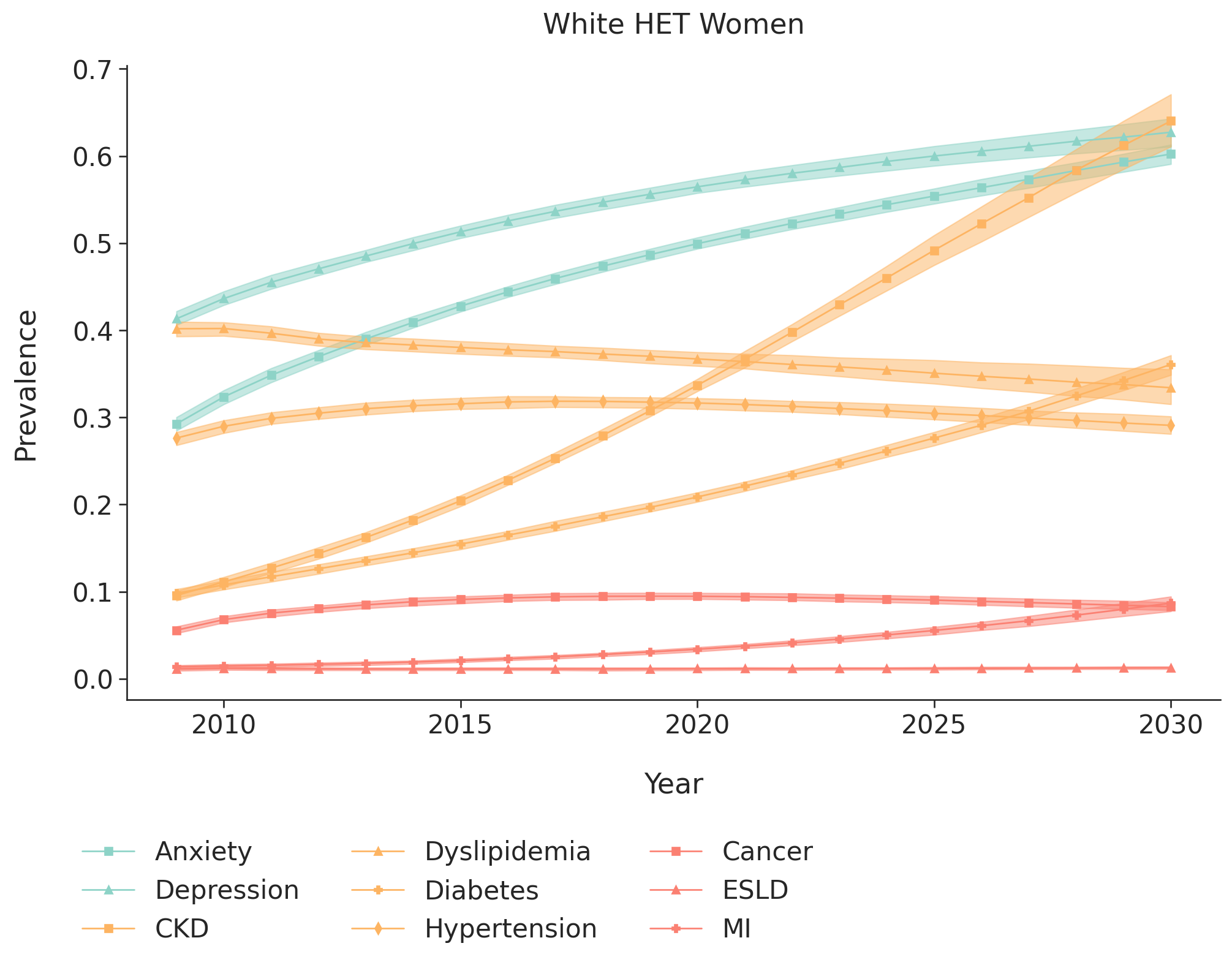

n) Black/African American heterosexual women

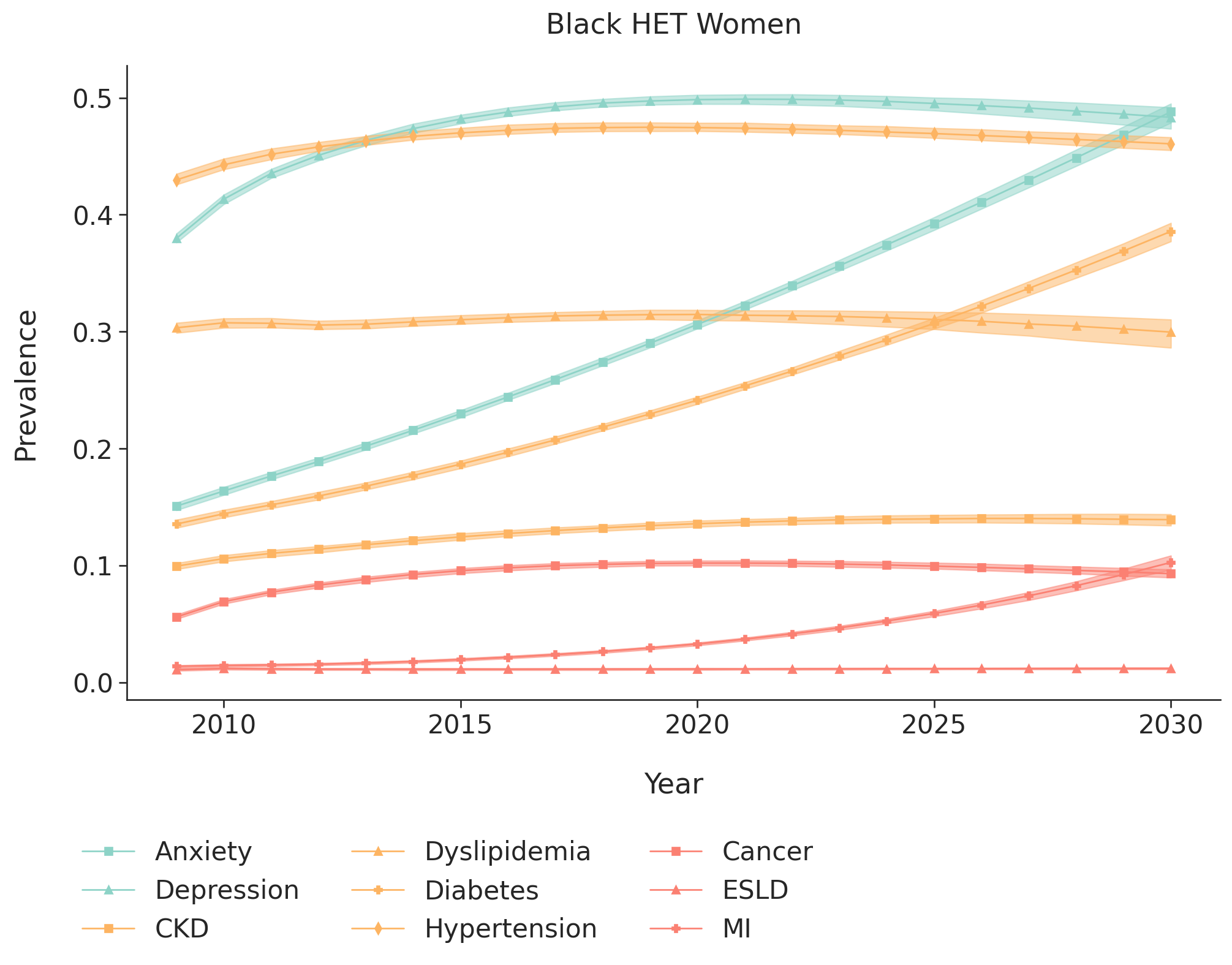

o) Hispanic heterosexual women

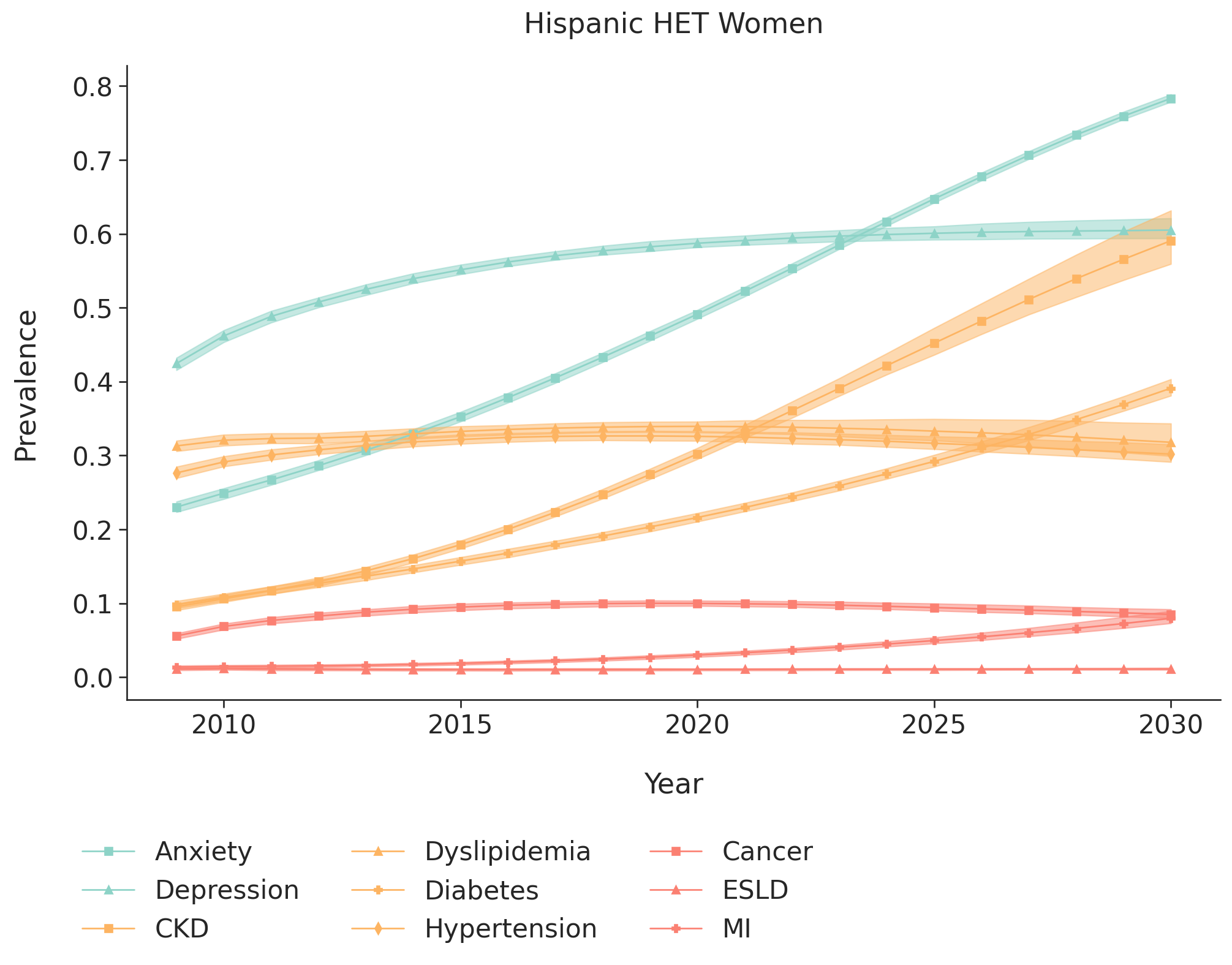

Footnotes:

CKD=stage ≥3 chronic kidney disease

ESLD=end-stage renal disease

MI=myocardial infarction

The 95% interquartile range is estimated as the 2.5% and 97.5% range of results from running the simulation 200 times.
